# Genome-wide association meta-analysis of spontaneous coronary artery dissection reveals common variants and genes related to artery integrity and tissue-mediated coagulation

**DOI:** 10.1101/2022.07.05.22277238

**Authors:** David Adlam, Takiy-Eddine Berrandou, Adrien Georges, Christopher P. Nelson, Eleni Giannoulatou, Joséphine Henry, Lijiang Ma, Montgomery Blencowe, Tamiel N. Turley, Min-Lee Yang, Peter S. Braund, Ines Sadeg-Sayoud, Siiri E. Iismaa, Matthew L. Kosel, Xiang Zhou, Stephen E. Hamby, Jenny Cheng, Lu Liu, Ingrid Tarr, David W.M. Muller, Valentina d’Escamard, Annette King, Liam R. Brunham, Ania A. Baranowska-Clarke, Stéphanie Debette, Philippe Amouyel, Jeffrey W. Olin, Snehal Patil, Stephanie E. Hesselson, Keerat Junday, Stavroula Kanoni, Krishna Aragam, Adam S. Butterworth, CARDIoGRAMPlusC4D, MEGASTROKE, International stroke genetics consortium (ISGC) intracranial aneurysm working group, Marysia S. Tweet, Rajiv Gulati, Nicolas Combaret, DISCO register investigators, Daniella Kadian-Dodov, Jon Kalman, Diane Fatkin, Jacqueline Saw, Tom R. Webb, Sharonne N. Hayes, Xia Yang, Santhi K. Ganesh, Timothy M. Olson, Jason C. Kovacic, Robert M. Graham, Nilesh J. Samani, Nabila Bouatia-Naji

## Abstract

Spontaneous coronary artery dissection (SCAD) is an understudied cause of acute myocardial infarction primarily affecting women. It is not known to what extent SCAD is genetically distinct from other cardiovascular diseases, including atherosclerotic coronary artery disease (CAD). Through a meta-analysis of genome-wide association studies including 1917 cases and 9292 controls of European ancestry, we identified 17 risk loci, including 12 new, with odds ratios ranging from 2.04 (95%CI 1.77-2.35) on chr21 to 1.25 (95%CI 1.16-1.35) on chr4. A locus on chr1 containing the tissue factor gene (*F3*), which is involved in blood coagulation cascade, appears to be specific for SCAD risk. Prioritized genes were mainly expressed in vascular smooth muscle cells and fibroblasts of arteries and are implicated predominantly in extracellular matrix biology (e.g. *COL4A1/A2, HTRA1* and *TIMP3*). We found that several variants associated with SCAD had diametrically opposite associations with CAD suggesting that shared biological processes contribute to both diseases but through different mechanisms. We also demonstrated an inferred causal role for high blood pressure, but not other CAD risk factors, in SCAD. Our findings provide novel pathophysiological insights involving arterial integrity and tissue-mediated coagulation in SCAD and set the stage for future specific therapeutics and prevention for this disease.

## Introduction

Cardiovascular disease is the leading cause of death in women but sex-specific aspects of the risk of heart disease and acute myocardial infarction (AMI) remain under-studied.^1^ Spontaneous coronary artery dissection (SCAD) and atherosclerotic coronary artery disease (CAD) are both causes of acute coronary syndromes leading to AMI.^2-6^ However, in contrast with CAD, SCAD affects a younger, predominantly female population,^7^ and arises from the development of a haematoma leading to dissection of the coronary tunica media with eventually formation of a false lumen, rather than atherosclerotic plaque erosion or rupture.^8^ SCAD has been clinically associated with migraine^9^ and extra-coronary arteriopathies including fibromuscular dysplasia (FMD).^10-13^ However, co-existent coronary atherosclerosis is uncommon.^8,14^ Whilst the genetic basis of CAD is increasingly well established,^15^ the pathophysiology of SCAD remains poorly understood.^4^ The search for highly penetrant mutations in candidate pathways or by sequencing has had a low yield, often pointing to genes involved in other clinically undiagnosed inherited syndromes manifesting as SCAD.^16^ Previous investigation of the impact of common genetic variation on the risk of SCAD has described 5 confirmed risk loci.^17-21^

Here we performed a meta-analysis genome-wide association studies (GWAS) comprising 1917 SCAD cases and 9292 controls of European ancestry. We identified 17 risk loci, including 12 new association signals demonstrating a substantial polygenic heritability for this disease. Importantly, we show that several common genetic risk loci for SCAD are shared with CAD but have a directionally opposite effect and a different genetic contribution of established cardiovascular risk factors. These findings implicate arterial integrity related to extracellular matrix biology, vascular tone, and tissue coagulation in the pathophysiology of SCAD.

## Results

### GWAS meta-analysis and SNP heritability

We conducted a GWAS meta-analysis from eight independent case control studies (**Supplementary Fig. 1, Supplementary Fig. 2, Supplementary Table 1**). Seventeen loci demonstrated genome-wide significant signals of association with SCAD, among which 12 were newly described for this disease, including one locus on chromosome X (**Table 1, Fig. 1A**).

**Table 1.**
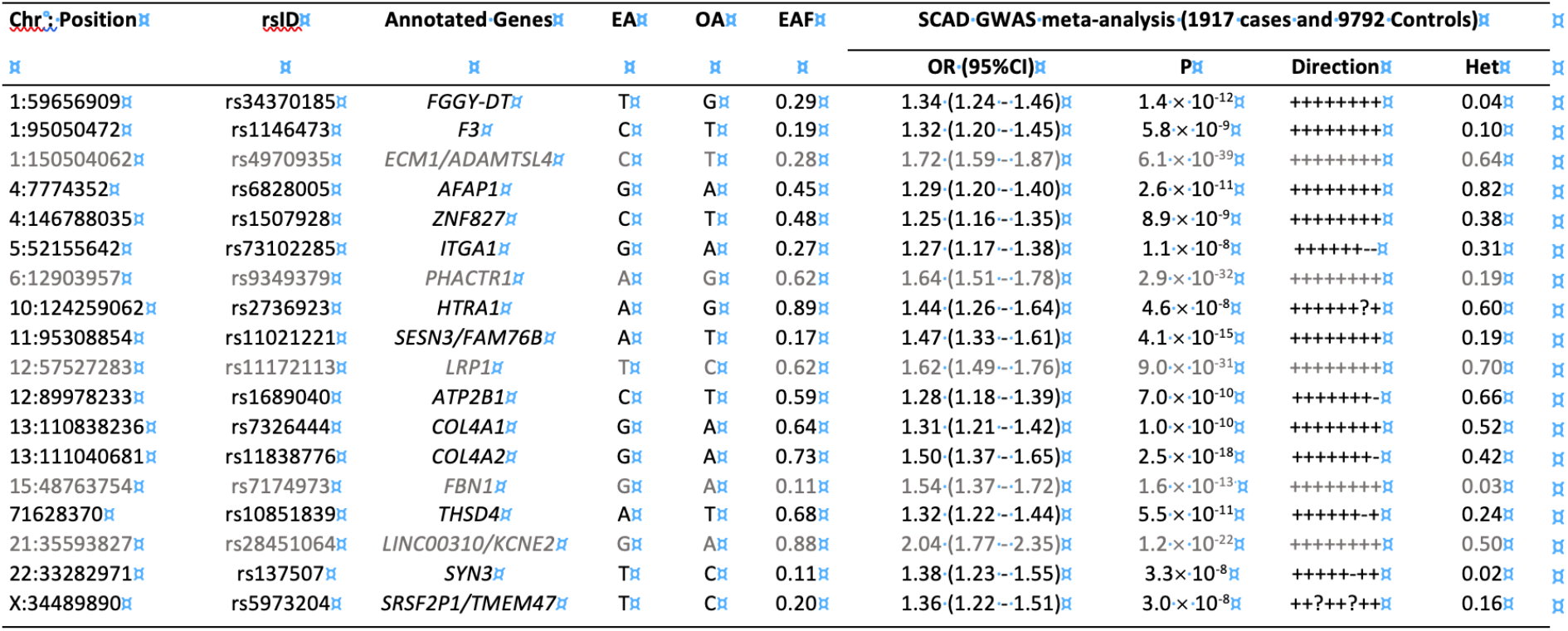
Lead associated variants at genome-wide significant SCAD loci. EA: effect alleles. OA: other alleles. EAF: effect allele frequencies. Het: P-value from the Cochran Q statistic heterogeneity test. Direction signs are provided for the individual association results in DISCO-3C, SCAD-UKI, Mayo Clinic, UBC/MGI, VCCRI, SCAD-UK II, VCCRIII and DEFINE-SCAD studies, respectively. Grey lines correspond to previously reported loci in SCAD.

**Figure 1.**
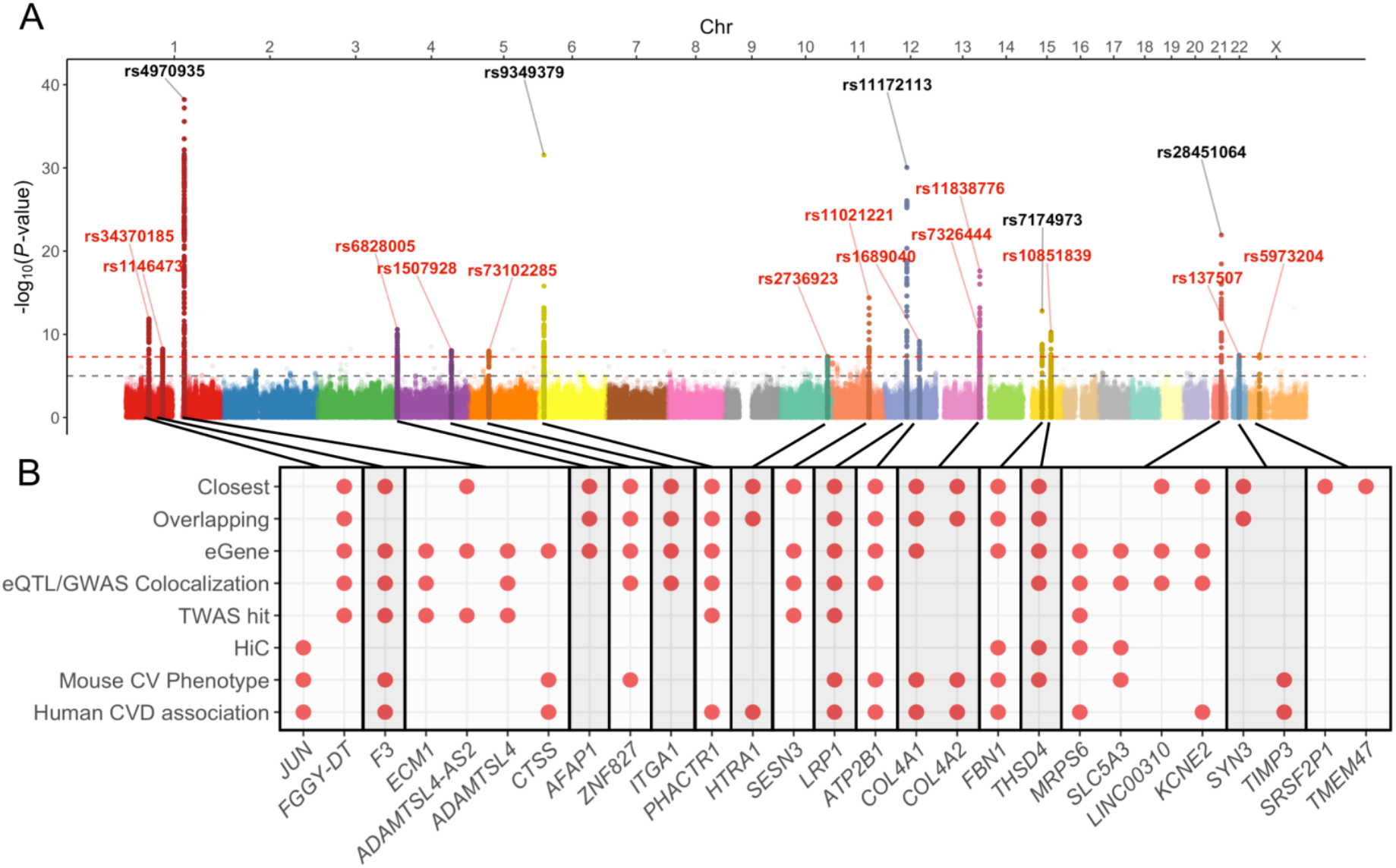
GWAS meta-analysis main association results, heritability estimates and gene prioritisation at risk loci. **A:** Manhattan plot representation of SNP-based association meta-analysis in SCAD. -log_10_ of association *P*-value (from a two-sided Wald test) is represented on the *y-*axis, genomic coordinates on the *x-*axis. SNPs located +/-500kb of genome-wide significant signals are highlighted. rsIDs of independent lead SNPs with *P*-value ≤ 5**×**10^−8^ are indicated. Label colours indicate newly identified loci (red) and previously known loci (grey). Dashed red line: *P*-value = 5×10^−8^. Dashed grey line: *P*-value = 1×10^−5^. **B:** Annotation of potential candidate genes at each SCAD locus. A red dot indicates that the corresponding gene fulfils one of the following eight criteria: 1) Closest gene upstream or downstream of SCAD lead SNP; 2) gene overlapping variants in the set of potential causal variants; 3) gene is an eGene of SCAD lead SNP in aorta, coronary artery, tibial artery, fibroblasts or whole blood samples from GTEx (v8 release); 4) colocalization of SCAD association signal and eQTL association in one of these 5 tissues; 5) the gene was a TWAS hit in any of the same 5 tissues; 6) promoter of the gene is in a chromatin loop with variants in the set of potential causal variants in human aorta tissue, as determined using HiC ^51^; 7) Gene knockout is known to be associated with a cardiovascular phenotype in mouse; or 8) an existing association of gene with a cardiovascular disease in humans. Prioritized genes included genes with most criteria at any loci and any genes fulfilling at least 3 criteria.

One locus on chromosome 4 (*AFAP1*) was recently reported for SCAD in the context of pregnancy^19^ is now confirmed as generally involved in SCAD (**Table 1)**. Estimated odds ratios of associated loci ranged from 1.25 (95%CI 1.16-1.35) in *ZNF827* on chromosome 4 to 2.04 (95%CI 1.77-2.35) on chromosome 21 near *KCNE2* (**Table 1**). We report evidence for substantial polygenicity for SCAD with an estimated SNP-based heritability above 0.70 (*h*^*2*^_*SNP*_ *=* 0.71 × 0.11 on the liability scale using LD score regression,^22^ and *h*^*2*^_*SNP*_ = 0.70 × 0.12 using SumHer^23^, **Supplementary Table 2)**. The *FALEC*/*ADAMTSL4-AS2* locus on Chr1 accounted for the largest proportion of heritability for SCAD in our dataset (*h*^*2*^ = 0.028), followed by the *COL4A1-COL4A2* locus, which contained 2 independent GWAS signals (*h*^*2*^ = 0.022, **Supplementary Table 3, Supplementary Fig. 3**). Overall, we estimate that the 16 autosomal loci explain ∼24% of the total SNP-based heritability of SCAD (**Supplementary Table 3)**.

### Functional annotation of variants in SCAD loci

We found SCAD-associated variants to be significantly enriched in enhancer marks specific to gene expression in arterial tissues from ENCODE ^24^ (e.g. aorta, tibial artery, thoracic aorta and coronary artery), as well as several smooth muscle cell rich tissues (e.g. colon, small intestine, uterus) (**Supplementary Fig. 4**). Based on recently published analyses of single-cell open chromatin in 30 adult tissues,^25^ we determined that vascular smooth muscle cells (VSMCs) and fibroblasts were the top enriched cell types for SCAD-associated loci among clusters represented in aorta and tibial artery datasets (**Fig. 2A, Supplementary Fig. 5**). Consistently, all but one SCAD locus included at least one variant that overlapped with enhancer marks or open chromatin peaks in coronary artery tissue, VSMCs or fibroblasts (**Supplementary Fig. 6, Supplementary Table 4**). Among SCAD top associated variants, 14 were expression quantitative trait loci (eQTLs) for nearby genes in aorta, coronary or tibial artery, whole blood or cultured fibroblasts (**Fig. 1B, Supplementary Table 5**).

**Figure 2.**
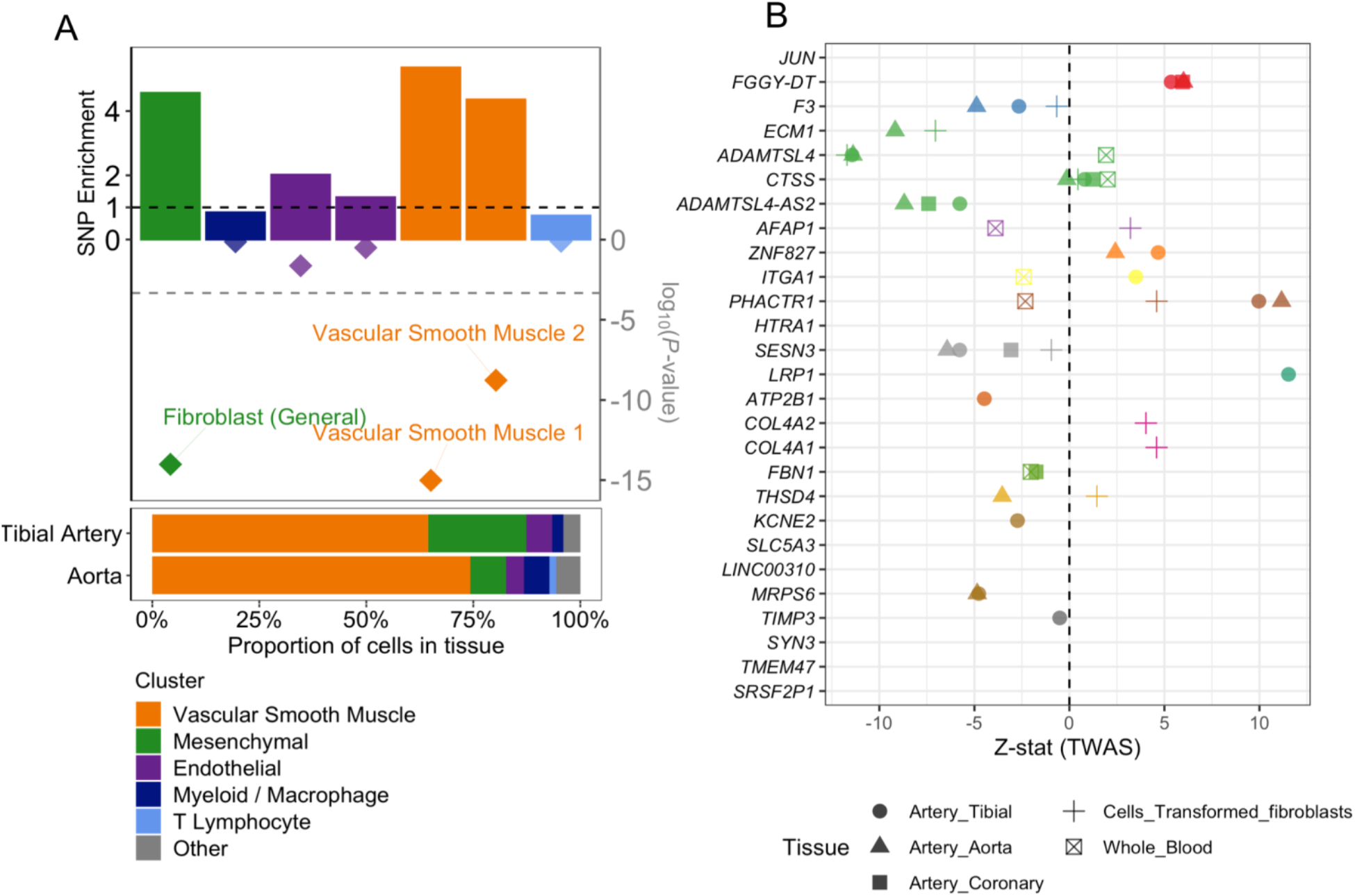
Enrichment of SCAD SNPs in open chromatin regions from arterial cells and genetically predicted expression changes of nearby genes. **A:** *Upper panel:* Representation of SCAD SNPs fold-enrichment (upper y-axis) and enrichment *P*-value (log scale, lower y-axis) among open chromatin regions of 7 single-cell sub clusters contributing to >1% of cells in artery tissue ^25^. A random background was created for each open chromatin dataset by randomly selecting 1000 sets of size and chromosome matched regions from the genome. Enrichment represents the ratio of the number of SCAD SNPs overlapping open chromatin regions over the average number of SCAD SNPs overlapping background regions. *P*-value was evaluated using a binomial test with greater enrichment as alternative hypothesis. Lower dashed line represents the threshold for significance (P<0.05) after adjusting for 105 sub clusters. *Lower panel*: Composition of artery tissues relative to 105 single-cell sub clusters determined from single-nuclei ATAC-Seq in 30 adult tissues ^25^. Only sub clusters representing more than 1% of cells from either aorta or tibial artery were represented. **B:** Representation of SCAD TWAS Z-score for SCAD candidate genes. Z-score is indicated along the *x*-axis. Point shape indicates the tissue for TWAS association. Point colour is used to distinguish between genes located in different SCAD loci.

### Tissue coagulation initiation as a novel biological mechanism and predominance of extracellular matrix biology in SCAD risk

We applied a multi-source strategy to identify candidate genes at risk loci. We prioritized genes that were *i)* the closest or the overlapping genes with the top associated variants, *ii)* targets of eQTLs colocalizing with GWAS signal (**Supplementary Fig. 7A, Supplementary Table 5)** or transcriptome-wide association study (TWAS) hits in at least one tissue relevant to arterial dissection (aorta, coronary or tibial artery, fibroblasts or whole blood from GTEx), (**Supplementary Fig. 7B, Supplementary Table 6**), *iii)* genes involved in significant long-range chromatin conformation interactions from Hi-C data with SCAD associated variants in aorta or *iv)* genes with biological function linked to the cardiovascular system in mice or humans. We identified 1 to 2 specific and strong candidate genes in 13 loci (**Fig. 1B**). For instance, tissue factor gene (*F3*) stood up as the most likely target gene near rs1146473 (OR=1.32, P=5.8 ×10^−9^), a locus on chromosome 1 that we describe as novel for SCAD or any cardiovascular disease or trait so far. *F3* is the closest coding gene to the association signal, and was a TWAS hit in artery tissues (**Supplementary Fig. 7B**). In addition, the rs1146473 risk allele for SCAD confidently (PP=94%) colocalised with an eQTL signal with *F3* in aorta supporting the genetic risk to be potentially through lower *F3* expression in arteries (**Supplementary Table 5, Fig. 2B**). Tissue factor, also known as coagulation factor III, forms a complex with factor VIIa, that is the primary initiator of blood coagulation. Hence, reduced factor III expression is potentially a key biological mechanism contributing to hematoma formation in coronary arteries of SCAD survivors.

To globally assess the biological mechanisms involving prioritized genes, we applied a network query based on Bayesian gene regulatory networks constructed from expression and genetics data from arterial tissues and fibroblasts.^26-28^ We found the extracellular matrix organisation to be the biological function where most prioritized genes and their respective immediate subnetworks clustered (**Supplementary Fig. 8**). Among genes we prioritized in novel loci, a number encode proteins involved in extracellular matrix formation, including integrin alpha 1 (*ITGA1*), basement membrane constituent collagen type IV alpha 1 chain (*COL4A1*) and alpha 2 chain (*COL4A2*), serine protease HtrA serine peptidase 1 (*HTRA1*), metallopeptidase thrombospondin type 1 domain containing 4 (*THSD4*), a partner of previously reported SCAD locus fibrillin 1 (*FBN1*), and TIM metallopeptidase inhibitor 3 (*TIMP3*). Of note, the *F3* sub-network also clustered in the extracellular matrix organisation and connected with *HTRA1* and *TIMP3* subnetworks through Bayesian network edges from aorta and coronary artery (**Supplementary Fig. 8**).

### Shared genetics between SCAD and arterial diseases

With the exception of the *F3* locus, SCAD risk loci located within 1 megabase of the top SCAD variants were at least suggestively (P<10^−5^) associated with other forms of cardiovascular and neurovascular disease (**Fig. 3A, Supplementary Table 7**). Using trait colocalization analyses, we found that the same variants were likely to be causal both for SCAD and the other diseases or traits at 15 loci (**Fig. 3A**). However, directions of effects were not systematically consistent across loci for all diseases.

**Figure 3:**
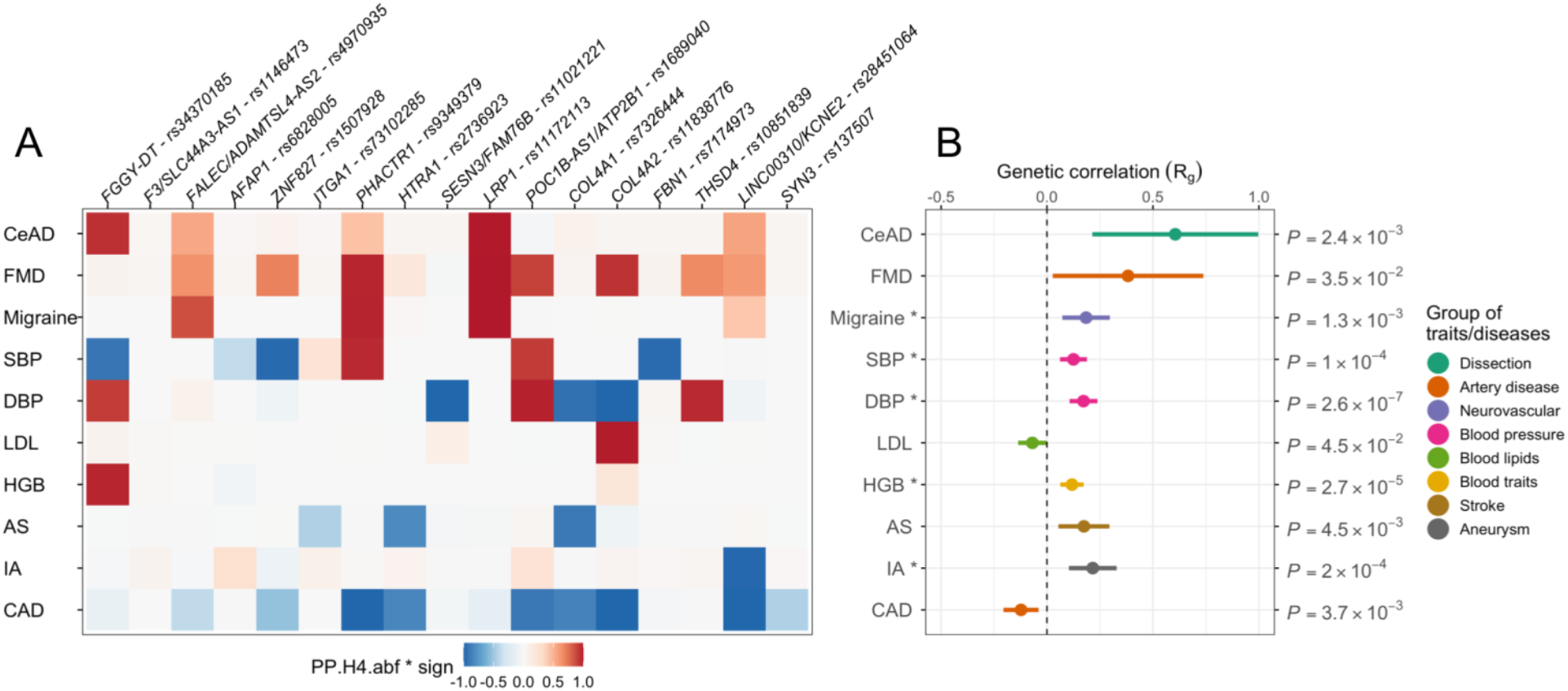
Colocalization and genetic correlation of SCAD genetic association with cardiovascular/neurovascular traits/diseases. **A:** Heatmap represents the colocalization of SCAD signals with GWAS analysis of 10 independent traits/diseases: cervical artery dissection (CeAD), multifocal fibromuscular dysplasia (FMD), migraine, blood pressure (SBP: systolic, DBP: diastolic), low-density lipoprotein cholesterol blood concentration (LDL), haemoglobin concentration (HGB), stroke (AS: any stroke), intracranial aneurysm (IA) and coronary artery disease (CAD). Tile colour represents the H4 coefficient of approximate Bayes factor colocalization, i.e. posterior probability for the two traits to share one causal variant at the locus (PP.H4.abf, 0 to 1), multiplied by the sign of colocalization (+1 is both traits have the same risk/higher value allele, −1 for opposed risk/higher value allele) on a blue/white/red color scale. **B:** Forest plot representing genetic correlation of SCAD and the aforementioned 10 traits/diseases. Rho coefficient of genetic correlation (Rg) is represented on the *x*-axis. Range indicates the 95% confidence interval. Unadjusted *P*-value of correlation is indicated. * indicates significant associations (*P*.*adj* < 0.05) after adjusting for 28 traits (**Supplementary Table 8**).

Globally, SCAD loci showed evidence for high posterior probability (PP) for the same risk alleles to also be likely causal for FMD and cervical artery dissection (CeAD) (**Fig. 3A, Supplementary Table 7**). LDSC-based genetic correlations indicated SCAD to positively correlate with FMD (rg=0.38 × 0.18; P=0.03) and CeAD (rg=0.61 × 0.20; P=2.4×10^−3^, **Fig. 3B, Supplementary Table 8**), which is consistent with the clinical observation of the frequent co-existence of these arteriopathies in SCAD patients. For instance, FMD is reported in ∼40 to 60% of SCAD patients.^11,29^ Stratified analyses in the largest four case-control studies where FMD arteriopathies were screened indicated globally similar associations with SCAD (**Supplementary Fig. 9, Supplementary Table 9**). Finally, genetic correlations indicated SCAD to positively correlate with several neurovascular diseases where predominantly arterial structure and/or function are altered, including stroke (rg=0.17 × 0.06; P=4.5×10^−3^), migraine (rg=0.18 × 0.06; P=1.3×10^−3^), intracranial aneurysm (rg=0.22 × 0.06; P=2.0×10^−4^), and subarachnoid haemorrhage (rg=0.27 × 0.07; P=6.4×10^−5^) (**Fig. 3B, Supplementary Table 8**).

### Opposite genetic link between SCAD and atherosclerotic coronary artery disease

While CAD patients are predominantly men (∼75%), who often have pre-existing cardiometabolic comorbidities, (mainly dyslipidaemia, hypertension and type 2 diabetes), SCAD patients are on average younger, present with fewer cardiovascular risk factors, and are overwhelmingly women (>90%).^2,4^ Using genetic association colocalization and genetic correlation we genetically compared SCAD with CAD. We found that among SCAD loci, several were known to associate with CAD. Disease association colocalization analyses showed that, for 6 loci, SCAD and CAD are likely to share the same causal variants with high posterior probabilities (PP H4: 84 to 100%) but all with opposite risk alleles (**Fig. 3A, Supplementary Table 7**). Genetic correlation confirmed a genome-wide negative correlation between SCAD and CAD (rg=-0.12 × 0.04; P=3.7×10^−3^) (**Supplementary Table 8**), including after conditioning SCAD GWAS results on SBP or DBP GWAS results using mtCOJO^30^ (rg_CAD/SBP_=-0.19 × 0.04, P= P=4.6×10^−6^; rg_CAD/DBP_=-0.19, × 0.04, P= P=1.3×10^−5^ (**Supplementary Table 10, Supplementary Fig. 10**).

### CVD risk factors and risk of SCAD and CAD

We found that SCAD shared several causal variants with systolic and diastolic blood pressure, involving both the same and opposite directional effects (**Fig. 3A, Supplementary Table 7**). We found several shared loci with haemoglobin levels and a significant genetic correlation with SCAD (rg=0.12 × 0.03; P=2.7×10^−5^, **Fig. 3B**). However, SCAD loci were not shared with BMI, lipid traits including LDL and HDL, type 2 diabetes or smoking and these traits did not correlate with SCAD at the genomic level (**Supplementary Tables 7 and 8**). Interestingly, we found significant positive genetic correlations both with systolic (rg=0.12 × 0.03; P=1.0 10^−4^) and diastolic (rg=0.17 × 0.03; P=2.6 10^−7^) blood pressure, indicating a shared genetic basis with SCAD (**Fig. 3B, Supplementary Table 8**). To assess the extent to which blood pressure and main cardiovascular risk factors may contribute to the risk of SCAD, we leveraged existing GWAS datasets to identify instrumental variables and conducted comparative Mendelian randomization associations with SCAD or CAD. We found robust significant associations estimated by inverse variance weighted (IVW), MR-Egger and weighted median methods between genetically-predicted blood pressure traits and increased risk of SCAD (beta_IVW-SBP_=0.05 × 0.01, P=7.6×10^−6^, beta_IVW-DBP_=0.10 × 0.02, P=1.9×10^−8^) and CAD (beta_IVW-SBP_=0.04 × 0.002, P=8.6×10^−49^; DBP: beta_IVW-DBP_=0.06 × 0.004, P=1.6×10^−44^) (**Fig. 4, Supplementary Table 11**). Similar associations were estimated when we analysed only women with SCAD, women with CAD and men with CAD; analyses only in men with SCAD were limited by the extremely small numbers of male cases (**Supplementary Table 12**). Genetically determined BMI, lipid traits, type 2 diabetes and smoking did not influence the risk for SCAD. However, we were able to overall confirm that these cardiometabolic traits are strong genetic risk factors for CAD (**Fig. 4)**. Our findings indicate that genetically elevated blood pressure is the only shared genetic risk factor between SCAD and CAD, albeit involving potentially different genetic loci.

**Figure 4.**
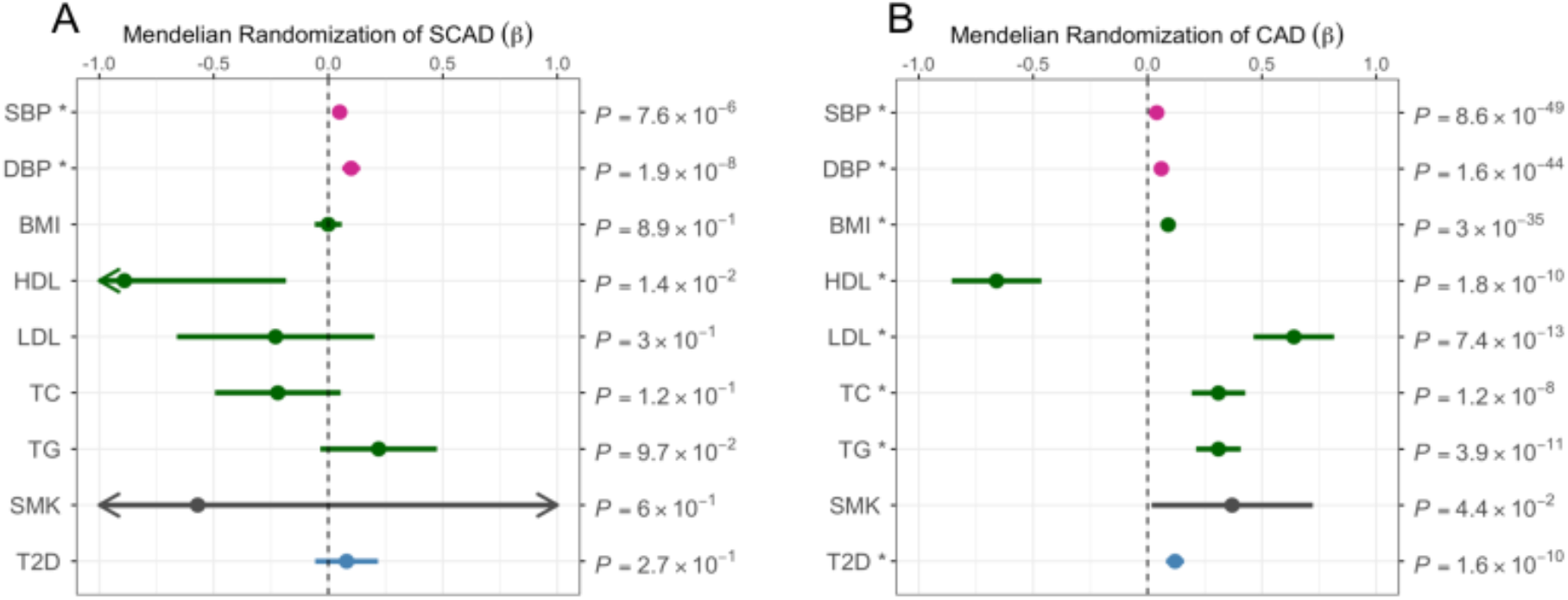
Mendelian randomization associations between main cardiovascular risk factors and SCAD or CAD. Forest plots representing Mendelian Randomization associations between main cardiovascular risk factors and SCAD (A) or CAD (B). Association estimates (β) obtained from Mendelian randomization analyses using inverse variance weighted method (IVW) are represented on the x-axis where range represents the 95% CI. Unadjusted *P*-values of the associations are indicated. Traits analysed were abbreviated as follows: Systolic blood pressure (SBP), Diastolic blood pressure (DBP), Body mass index (BMI), High-density lipoprotein (HDL), Low-density lipoprotein (LDL), Total cholesterol (TC), Triglycerides (TG), Smoking (SMK).

## Discussion

Here we provide the largest study to date aimed to understanding the genetic basis of SCAD, an understudied cause of AMI that primarily affects women. We report novel associations and demonstrate high polygenic heritability for SCAD. We leverage integrative functional annotations to prioritize genes, which are predominantly expressed by VSMCs and fibroblasts of arteries. Insights from biological functions of genes highlight the central role of extracellular matrix integrity and reveal impaired tissue coagulation as a novel potential mechanism for SCAD. Globally, we demonstrate the polygenic basis of SCAD to be shared with an important set of cardiovascular diseases. However, a striking directionally opposite genetic impact is found with atherosclerotic coronary artery disease (CAD), involving multiple risk loci and leading to a genome-wide negative genetic correlation. We provide evidence supporting that genetically predicted higher blood pressure as an important risk factor for SCAD, but not other well-established cardiovascular factors. Our results set the stage for future investigation of novel biological pathways relevant to both SCAD and CAD and potential therapeutic and preventive strategies specifically targeting SCAD.

As an understudied condition, previously thought to be uncommon, SCAD was initially suspected to involve rare and highly penetrant mutations. However, recent sequencing studies have suggested that only a small proportion (∼3.5%) of SCAD cases are due to rare variants.^16,31^ This is in keeping with increasing clinical recognition suggesting that this condition is not rare.^2,4^ Despite a modest sample size, we identified 17 risk loci accounting for about a quarter of the polygenic heritability that we estimate to be ∼71% indicating SCAD therefore to be predominantly a complex polygenic disease.

This study supports the presence of genetic overlap between risk of SCAD and other vascular diseases, involving generally younger individuals and more women such as cervical arterial dissection, migraine, subarachnoid haemorrhage and FMD. These conditions are reported to occur at increased frequency in SCAD patients,^10-13^ supporting shared causal biological mechanisms. Among the genes we prioritize as novel SCAD loci, we highlight the ATPase plasma membrane Ca2+ transporting 1 gene (*ATP2B1*) that we have recently reported to associate with FMD,^32^ a well-established locus for blood pressure risk^33^ via its role in VSMCs intracellular calcium homeostasis and blood pressure regulation.^34^ Most importantly, we provide evidence for a causal genetic effect of both systolic and diastolic blood pressure in SCAD risk. These findings provide an important genetic basis to support observational data suggesting control of blood pressure may be an important factor in reducing the risk of recurrence after SCAD.^35^ However, our findings also suggest that controlling other causal risk factors for CAD, such as LDL cholesterol with statins, may confer less benefit in SCAD than in CAD.

Knowledge of the molecular mechanisms leading to SCAD has been limited. Insights from sequencing studies of rare genetic variants has shown most are associated with genes known from hereditary connective tissue disorders such as vascular Ehlers-Danlos, Loeys-Dietz, Marfan syndromes, and adult polycystic kidney disease.^16,31^ In this study, we identify the regulation of the extracellular matrix of arteries as the predominant polygenetic biological mechanism for SCAD. Integrative prioritisation analyses revealed 13 potential causal genes with established key roles in maintaining arterial wall integrity and function. Among these, we highlight the serine protease *HTRA1* and metallopeptidase inhibitor *TIMP3* that are involved in matrix disassembly. Interestingly, we found a novel association signal with SCAD in the metallopeptidase thrombospondin type 1 domain containing 4 gene (*THSD4*) that promotes fibrillin-1 (FBN1) elastic fibre assembly, and confirm the previously reported associations with *ADAMTSL4* and *FBN1*.^18,20^ We showed that genetically decreased expressions of these genes in arteries were correlated with higher SCAD risk alleles in arteries or fibroblasts. This finding suggests that a genetic predisposition to a weaker extracellular matrix may increase the vulnerability of traversing intramural micro-vessels to disruption, increasing the risk of initiation and propagation of a false lumen within the coronary vessel wall leading to SCAD.

A striking finding from our study is the identification of tissue factor gene (*F3*), a critical component of tissue-mediated blood coagulation, as a strong candidate gene in a risk locus for SCAD. We found that genetically determined lower expression of *F3* in arterial tissue was associated with a higher risk for SCAD, involving variants located in putative functional regulatory elements in the coronary artery, VSMCs and fibroblasts. Tissue factor is synthesized at the subendothelial level of VSMCs and by fibroblasts in the adventitia surrounding the arteries.^36^ In SCAD, once an intramural haemorrhage has initiated, propagation and pressurisation of the false lumen may depend, in part, on coagulation and stabilisation of the haematoma. Tissue factor is widely studied in the context of pro-thrombotic conditions, including atherosclerosis, although notably the genetic variants we describe here do not associate with atherosclerotic disease. This feature is an exception to the highly pleiotropic nature of the variants we describe in the remaining SCAD loci, suggesting that impaired tissue-initiated coagulation as a putative specific mechanism in SCAD.

Many of the risk loci and their prioritized genes we report here for SCAD are already known from atherosclerotic disease GWAS. However, here we provide compelling and intriguing evidence for the opposite directionality of a substantial fraction of genetic basis for SCAD *versus* CAD, suggesting some key biological mechanisms involved in the two diseases are also likely to be opposite. For example, the association signals in the *COL4A1-COL4A2* locus are in an opposite direction to their contribution to CAD.^37^ This locus encodes α1 and α2 chains of type IV collagen, with transcripts generated through a common promoter. Type IV collagen is the main component of basement membrane of arterial cells and plays a key role in the structural integrity and biological functions of VSMCs in the *tunica muscularis*. Reduced collagen IV expression increases the risk of CAD.^15,38^ Proposed potential mechanisms for this include a disinhibition of VSMC-intimal migration during atherogenesis or an increase in the vulnerability of atherosclerotic plaque to rupture.^38^ In contrast to CAD, our data indicates that genetically mediated increased collagen IV expression also increased the risk of SCAD. Better understanding of how these directionally opposite changes modify risk of CAD and SCAD have considerable potential to enhance understanding of the molecular genetic mechanisms that confer risk in both diseases.

## Data Availability

All data in the present study will be available upon request to the authors

## Authors contributions

Writing of the manuscript: DA, T-EB, AG, NJS, NB-N. Study design / conception of analyses: DA, T-EB, AG, CPN, EG, MST, RG, DK-D, JS, SNH, XY, SKG, TMO, JCK, RMG, NJS, NB-N. Genotyping: DA, AG, CPN, LM, TNT, MLY, PSB, SEI, MLK, SHE, LL, Vd’E, AB-C, KJ, TRW, SKG, TMO, JK, NB-N. Sample /phenotype contribution: DA, IT, DWMM, Vd’E, AK, LB, SD, PA, JWO, SEH, MST, RG, NC, DK-D, JS, LB, SKG, JK, DF, SNH, JCK, RMG. Data analysis: T-EB, AG, CPN, EG, JH, LM, MB, M-LY, IS-S, MLK, XZ, JC, IT, SP, SGK, KA, AB, XY, JCK, NB-N. Scientific editing of the manuscript: DA, T-EB, AG, JH, TNT, SKG, MST, RMG, TRW, SNH, WY, TMO, JCK, NJS, NB-N.

## Methods

### Patients and control populations

The meta-analysis included participants of European ancestry from eight studies: DISCO-3C, SCAD-UK I, SCAD-UK II, Mayo Clinic, DEFINE-SCAD, CanSCAD/MGI, VCCRI I and VCCRI II. SCAD patients presented similar clinical characteristics (**Supplementary Table 1**) and homogeneous diagnosis, exclusion and inclusion criteria. All studies were approved by national and/or institutional ethical review boards. Further study-specific clinical details are provided in the **Supplementary Material**.

### Genome-wide association meta-analysis

Details on the pre-imputation quality control steps for each study are listed in **Supplementary Table 13**. In brief, genotyping was performed using commercially available arrays or genome sequencing (SCAD-UK Study II and VCCRI Study II). To increase the number of tested SNPs and the overlap of variants available for analysis between different arrays, all European ancestry cohorts, except for SCAD-UK Study II and VCCRI Study II, had their genotypes imputed to HRC v1.1 reference panel^39^ on the Michigan Imputation Server.^40^ GWAS was conducted in each study under an additive genetic model using PLINK v2.0.^41^ For chromosome X, males and females were both on a 0..2 scale under the chromosome X inactivation assumption model. Models were adjusted for population structure using residues from the first five principal components and sex, except in the women-only analyses. Prior to meta-analysis, we removed single nucleotide polymorphisms (SNPs) with low minor allele frequencies (MAF< 0.01), low imputation quality (r^2^ < 0.8), and deviations from Hardy-Weinberg equilibrium (*P* < 10^− 5^). A total of 6,697,670 variants met these criteria and were kept in the final results.

Individuals studies GWAS results were combined using an inverse variance weighted fixed-effects meta-analysis in METAL software,^42^ with correction for genomic control. Heterogeneity was assessed using the I^2^ metric from the complete study-level meta-analysis. Between-study heterogeneity was tested using the Cochran Q statistic and considered significant at *P* ≤ 10^−3^. Genome-wide significance threshold was set at the level of *P* = 5.0 × 10^− 8^. LocusZoom (http://locuszoom.org/) was used to provide regional visualization of results.

### Functional annotation

#### Identification of potential functional variants

To generate a list of potential functional variants, we first identified the 95% credible set of variants using ppfunc function of corrcoverage R package (v1.2.1). Posterior probability of causality was evaluated from marginal Z-scores for all variants within 500kb of the lead SNP at each locus. In the *COL4A1-COL4A2* locus, where we found two association signals, these were separated by placing an equidistant border from each lead SNP for the inclusion of SNPs in the analysis. Variants with cumulated posterior probability up to 95% were kept for further analyses. To consider potentially poorly imputed variants in one of the individual case-control studies, we also included variants in high LD (r^2^>0.7) with the lead SNP at each locus based on information from European populations (1000 Genome reference panel) queried using ldproxy function of LDlinkR package (v1.1.2).^43^

#### Enrichment of SCAD variants in regulatory regions

To calculate the enrichment of SCAD associated SNPs among functionally annotated genomic regions, we retrieved available H3K27ac ChIP-Seq datasets (narrowpeaks beds) in any tissue from ENCODE (https://www.encodeproject.org/,^44^ and single nuclei ATAC-Seq peak files (bed format) from Human enhancer atlas (http://catlas.org/humanenhancer.^25^ A complete list of datasets is available in **Supplementary Table 14**. For H3K27ac marks, bed files corresponding to the same tissue were concatenated and sorted before combining overlapping peaks using bedtools (v2.29.0) merge command. Variant enrichment was calculated using rtracklayer (v1.52.1) package as follows: for each peak file, 1000 datasets matched by peak number and sizes for each chromosome were generated. Positions of SCAD potential functional variants were converted to hg38 genomic coordinates using UCSC liftover tool (https://genome-store.ucsc.edu/). We counted overlaps of SCAD potential functional variants with peak file and background files using countOverlaps function. Variant enrichment was defined as the ratio of the number of overlaps between peak set and average of background sets. Enrichment *P*-value was evaluated using a binomial test with higher enrichment as alternative hypothesis. *P*-values were adjusted for multiple testing by the application of the Bonferroni correction.

#### Identification of variants with potential regulatory function

We used H3K27ac peaks in coronary arteries as described above, open chromatin regions in healthy coronary arteries obtained as previously described ^32,45^ and open chromatin regions from merged snATAC-Seq clusters, which were mapped fragments from snATAC-Seq in 25 adult tissues that we retrieved from GEO (GSE184462^25^) in bed format. Mapped fragments from all clusters representing more than 1% of cells in at least one arterial tissue (T lymphocyte 1, CD8^+^), endothelial general 2, endothelial general 1, macrophage general, fibro general, vasc sm muscle 2, vasc sm muscle 1) were extracted and grouped by annotated cell type as T-lymphocytes, macrophages, fibroblasts, endothelial cells, and vascular smooth muscle cells, respectively. Genome coverage was calculated using bedtools (v2.29.0) coverage function. We detected peaks from bedGraph output using MACS2 bdgpeakcall function (Galaxy Version 2.1.1.20160309.0) on Galaxy webserver.^46,47^ All peak files were extended 100bp upstream and downstream using bedtools (v2.29.0) slop function. We detected overlaps of SCAD potential functional variants with relevant genomic regions using findOverlap function from rtracklayer package (v1.52.1).^48^ We used Integrated Genome Browser (IGB, v9.1.8) to visualize read density profiles and peak positions in the context of human genome.^49^

#### Gene prioritization

Genes located within 500kb of lead variants were annotated to prioritize most likely causal genes. To find closest gene(s) from top SNP and genes overlapping variants in the list of potential functional SNPs, gene coordinates were retrieved from Gencode release 38, aligned to hg19 genomic coordinates (gencode.v38lift37.annotation.gff3.gz). Significant eQTL associations and all SNP-gene eQTL associations in v8 release of GTEx database were retrieved from GTEx website (www.gtexportal.org/home/datasets). Colocalization of association with SCAD and eQTLs was evaluated using R coloc package (v5.1.0) with default values as priors. We considered there was evidence for colocalization if H4 coefficients were over 75% or if eQTL association was significant for SCAD lead SNP and H4 was over 25%. TWAS was performed using FUSION R/python package.^50^ Gene expression models were pre-computed from GTEx data (v7 release) and were provided by the authors. Only genes with heritability P-value < 0.01 were used in the analysis. Both tools used LD information from the European panel of the 1000 Genomes phase 3. Bonferroni multiple testing correction was applied using the p.adjust function in R (v 4.1.0). Significant Capture HiC hits in aorta tissue were provided as supplementary data by Jung et al.^51^ Genes associated to mouse cardiovascular phenotypes (code MP:0005385) were retrieved from Mouse Genome informatics (www.informatics.jax.org) database.^52^ We also queried Disgenet database using disgenet2r package (v0.99.2) for genes with reported association to human cardiovascular disease (code C14) with a score over 0.2, including “ALL” databases.^53^ At each locus, we prioritized genes fulfilling at least 3 of the abovementioned criteria and those genes with the most criteria if these were less than three.

#### Bayesian network query of SCAD candidate genes

Gene expression data from aorta artery, coronary artery, tibial artery, and cultured fibroblast was curated from the Genotype Tissue Expression (GTEx) version 8.^27^ Gene expression data from the mouse aorta was curated from the Hybrid Mouse Diversity Panel (HMDP).^26^ Tissue-specific gene regulatory Bayesian networks (BNs) were constructed from the GTEx and HMDP gene expression data using RIMBANET.^28^ The BN from each dataset included only network edges that passed a probability of >30% across 1000 generated BNs starting from different random genes. BNs were combined for the top GWAS hits query, and mouse gene symbols were converted to their human orthologs. BNs were queried for the identified top GWAS hits to identify their first-degree network connections and to determine connections between their surrounding subnetwork nodes. The direction of edges was informed by prior knowledge such as eQTLs and previously known regulatory relationships between genes. Subnetworks were annotated by top biological pathways representative of the subnetwork genes by Enrichr with FDR <0.05.

### Colocalization with other traits and diseases

Summary statistics were retrieved from individual studies as indicated in **Supplementary Table 15**. At each locus, we selected variants found in both SCAD and the other studies with high quality of imputation (r^2^>0.9) and located within 500kb from the SCAD lead SNP. *COL4A1-COL4A2* loci were separated by placing an equidistant border from SCAD lead SNPs for the inclusion of SNPs in the analysis. Signal colocalization was evaluated using R coloc package (v5.1.0) with default values as priors. We reported H4 coefficients indicating the probability for two signals to share a common causal variant at each locus.

### Heritability estimates and genetic correlation attributable to common variants

We used LD score regression (LDSC)^22^ implemented in the ldsc package (v1.0.1, https://github.com/bulik/ldsc/) and SumHer^23^ implemented in LDAK software (www.ldak.org) to quantify the heritability explained by common variants or SNP-based heritability (h^2^_SNP_) for SCAD and the degree of genetic correlation between SCAD and other diseases and traits. We also used SumHer to estimate the SNP-based heritability attributable to loci associated with SCAD at genome-wide statistical significance. Loci were defined as 1 megabase region around lead SNPs in the GWAS meta-analysis. SNPs belonging to each locus were used as annotations to calculate the partitioned heritability. Two analyses were performed, one that considered separated loci and a second analysis that aggregated all SNPs as one annotation. Summary statistics were acquired from the respective consortia and are detailed in **Supplementary Table 15**. For each trait, we refined the summary statistics to the subset of HapMap 3 SNPs to reduce potential bias due to poor imputation quality. Correlation analyses were restricted to European ancestry meta-analyses summary statistics. We used the European LD-score files calculated from the 1000G reference panel and provided by the developers. A *P*<1.7×10^−3^, corresponding to adjustment for 30 independent phenotypes was considered significant. We conditioned SCAD association on cardiometabolic traits genetic association using multi-trait-based conditional and joint analysis (mtCOJO) tool from GCTA pipeline.^30^ The resulting summary statistics were then used to compute genetic correlations between SCAD, conditioned on cardiometabolic traits and trait of interest.

### Mendelian randomisation (MR) analyses

We used the multiplicative random-effects inverse variance weighted (IVW) method^54^ implemented in the TwoSampleMR R package to estimate the associations of genetically predicted cardiovascular risk factors that included blood pressure (SBP, DBP), lipids (HDL, LDL, TC, TG), body mass index (BMI), smoking liability and type 2 diabetes (T2D) with each of the outcomes of interest (SCAD or CAD). Estimates were scaled to a doubling in genetically predicted smoking risk, or to a one-unit increase in genetically predicted trait for the continuous traits. We performed sensitivity analyses using the weighted median and MR-Egger methods to assess the consistency of estimates under alternative assumptions about genetic pleiotropy as recommended.^54^ We also performed Cochran’s Q test to assess the heterogeneity between estimates obtained using different variants. As 11 risk factors were assessed, a Bonferroni-corrected significance level of 0.05/11= 4.5×10^−3^ was used as the threshold for statistical significance in this analysis. P-values between 4.5×10^−3^ and 0.05 are described as suggestively significant.

