## Supplementary for "Genome-wide association meta-analysis of spontaneous coronary artery dissection reveals common variants and genes related to artery integrity and tissue-mediated coagulation"

### Supplementary Material

SCAD Genetic Study

Adlam et al.,

### Supplementary Methods

#### Cohorts specific clinical and genetic analyses methods

##### DISCO-3C case control study

DISCO study was registered under Clinical Trials ID: NCT02799186, and approved by regional committee CPP (*comité de protection des personnes*) Sud-Est 6 2016 AU-1258. 3C study protocol was approved by «*comité consultatif de protection des personnes dans la recherche biomédicale*» Bicêtre Hôpital Bicêtre n°99-28 CCPPRB approved 10/06/99, 11/03/2003 and 17/03/2006.

Data from a national French registry of SCAD cases (51 cardiology centres) were analysed prospectively and retrospectively. All subjects 18 years old and above gave their written, informed consent to participate in the study. Exclusion criteria included in custody or guardianship, patients with iatrogenic or traumatic dissection or atherosclerotic dissection. SCAD was identified as an acute or chronic coronary syndrome defined according to universal recommendations with angiographic signs suggestive of SCAD[1] on the initial coronary angiography. In context of MINOCA, a pathological cardiac magnetic resonance (CMR) could lead to the diagnosis of SCAD retrospectively by rereading the angiograms. In case of ambiguity, intravascular ultrasound (IVUS), optical coherence tomography (OCT) or repeat coronary angiography was performed at the discretion of the operator to confirm the diagnosis of SCAD. All SCAD cases were subsequently confirmed by a core lab of 3 operators (NC, PM, GS) experienced in the field of SCAD and intra coronary imaging. Coronary segmentation was defined according to the AHA classification[2]. Screening for FMD was done at the discretion of the different centres either by CT scan, magnetic resonance, angiography or Doppler in cervico-cephalic, visceral and iliac arteries. The diagnosis of FMD was considered confirmed if at least one extra-cardiac location was verified by an experienced radiologist (LC) according to established criteria [3]. Complete screening was defined from brain to pelvis.

Blood samples were taken at the time of inclusion (during hospitalization for the prospective cohort or remotely in the retrospective cohort). DNAs was extracted from whole blood using a ChemagicTM 360 device (Perkin Elmer). Genotyping quality control was performed using PLINK v 2.022[4]. We checked for sex mismatches, first degree relatedness and ancestry estimation. Genotyping data were filtered using the criteria of Hardy-Weinberg equilibrium exact test p value smaller than 0.001 (--hwe 0.001, default value in PLINK 2.0) and missingness per marker >1% (--geno 0.01). The minimum allele frequency was set to 0.01 (--maf 0.01). Data was imputed using minimac4 incorporated in a cloud-based imputation server using HRC r1.1 2016 reference panel[5].

The Three-City Study (3C Study) is a population-based longitudinal study of the relation between vascular diseases and dementia in persons aged 65 years and older[6]. Participants were recruited from three French cities: Bordeaux (South-West), Dijon (North-East) and Montpellier (South-East). The 3C Study extended from 1999 to 2012. Participants underwent regular extensive examination. Examination included measurements of traditional vascular risk factors (blood pressure, glycaemia, lipids, etc.), cognitive functions, and subclinical vascular diseases using carotid ultrasound and cerebral magnetic-resonance imaging (MRI). DNA collection and genotyping were described previously[7].

**SCAD-UK case-control studies**

The UK SCAD study (ISRCTN42661582) was approved by the UK National Research Ethics Service (14/EM/0056) and the UK Health Research Authority.All patients gave written informed consent to participate in the study. Patients with SCAD were ascertained either by clinician referral to the UK national SCAD clinical service or by self-referral to an online portal. Only patients with SCAD confirmed on invasive angiography were included. Patients with atherosclerotic, iatrogenic or traumatic coronary dissections were excluded. Demographic data was obtained from the patients' medical record and from the patient through an online survey and direct contact as required. Hypertension was defined as being on treatment prior to the first clinical SCAD event. Gestational hypertension or diabetes were not included. All angiograms were jointly reported by at least 2 SCAD-experienced interventional cardiologists. Cross sectional imaging by MRA or CTA either undertaken on research or clinical grounds were used to determine the presence or absence of extra-coronary SCAD-associated arteriopathies. Those with FMD, aneurysms or dissections were considered abnormal. Those without were considered normal. Tortuosity or focal stenotic disease were not considered abnormal in the absence of these other features.

Sequencing of SCAD cases for arm I of UK SCAD study was described previously [8]. Genomic DNA from SCAD cases was extracted and underwent paired-end 150bp WGS at Human Longevity Inc using the NovaSeq6000 platform. For SCAD cases, >98% of consensus coding sequence release 22 (CCDS) has at least 10x coverage and average coverage of the CCDS achieved 42-fold read-depth. Variants were lifted over to genome assembly GRCh37 using NCBI remap and filtered to maf > 0.01 and hwe > 1e-6 using plink 1.9. Variant identifiers were updated to UK biobank formatting and individual sample files merged. SCAD-UK II case genomic DNA were genome-wide genotyped on the Illumina-Infinium™ Global Screening Array-24 v2.0 +MD array (665,608 markers) at the Spanish National Cancer Research Centre (CNIO), in the Human Genotyping lab, a member of National Genotyping Center (CeGen) [9], whereas genotyping of the SCAD-UK II control genomic DNA were genome-wide genotyped at Applied Biosystems™ Axiom™ Genotyping Services (Thermo Fisher Scientific) using the Applied Biosystems™ Axiom™ UK Biobank WCSG Genotyping Array (825,928 markers), as part of the UK Biobank genotyping project[10]. SCAD-UK II cases and controls were pre-imputation QC filtered using PLINK v1.90b6.14. Variants were excluded if the SNP call rate (CR) < 95% for minor allele frequency (maf) > 0.05 and CR < 98% for maf ≤ 0.05 for all chromosomes 1-22, X (except for cases in the X chromosome, where due to a poorer call rate we excluded variants only if the SNP CR was < 90% for maf > 0.05 and CR < 95% for maf ≤ 0.05), maf < 0.005, hwe deviation was P < 1 x 10-06. Samples were removed if there was a sex-mismatch between reported and genetically inferred sex or have an individual CR < 95% after SNP QC. Further samples were removed if relatedness using a PLINK PI-HAT was > 0.1875, or if samples had high heterozygosity, deviating from the mean sample heterozygosity rate (above ± 3 standard deviations). Will Rayner's HRC checking tool was used to assess and correct PLINK files for strand and REF/ALT orientation of alleles, using the Haplotype Reference Consortium release

1.1 (HRC r1.1) reference panel[11]. SCAD-UK II cases and controls were separately imputed on the Michigan Imputation Server with validated vcf.gz input files for each chromosome (1-22, X) with eagle (2.4) for pre-phasing and minimac4 (1.5.7) for imputation using either the Haplotype Reference Consortium release 1.1 (HRC r1.1) reference panel or the 1000g-phase-3-v5 (https://www.internationalgenome.org/) reference panel. Post imputation the two reference panels were combined using QCTOOLv2 (https://www.well.ox.ac.uk/~gav/qctool_v2/). To combine the two reference panels the 1000g-phase-3-v5 imputation variant dataset were merged onto the HRC r1.1 imputation variant dataset, so that only variants in addition to the HRC imputation were added. Subsequently, the cases and controls were combined again using using QCTOOLv2. Post-imputation QC excluded all SNPs with an imputation R-square < 0.3, hwe deviation of P < 1 x 10-06 in cases or P < 1 x 10-04 in controls, a minor allele count < 20 or maf < 0.01 in cases.

**VCCRI case control studies**

The study was approved by the St. Vincent's Hospital Human Research Ethics Committee (HREC/16/SVH/338, protocol number SVH 16/245) and conducted in accordance with the Australian National Health and Medical Research Council's National Statement on Ethical Conduct in Human Research and the CPMP/ICH Note for Guidance on Good Clinical Practice. Arm II control sample study was approved by the St. Vincent's Hospital Human Research Ethics Committee (HREC/17/SVH/315) and conducted in accordance with the Australian National Health and Medical Research Council's National Statement on Ethical Conduct in Human Research and the CPMP/ICH Note for Guidance on Good Clinical Practice. Ethics approval number 2015.028 for samples from Royal Melbourne Hospital, Melbourne, Victoria, Australia. SCAD cases were recruited via a social media platform or through direct referral from cardiologists. SCAD diagnosis was confirmed by review of coronary angiogram images by an expert interventional cardiologist (DM) blinded to the results of the genetic analysis. FMD diagnosis or negative scan was self-reported by subjects. Screening was partial in some cases. Subjects were considered to have hypertension if they were prescribed blood pressure lowering drugs before the SCAD occurred. All subjects were asked the clinical diagnostic criteria for migraines. Control samples for arm I of VCCRI study are from the MRGB cohort[12]. For the arm II of VCCRI study, control samples were unrelated atrial fibrillation cases and unrelated dilated cardiomyopathy patients and unrelated unaffected family members.

For VCCRI I, genomic DNA was extracted (PureLink GenomicDNA Mini Kit; Invitrogen) from buccal cells collected using a cheek swab. WGS was performed using paired-end KAPA PCR-Free v2.1 libraries on the Illumina HiSeq X Ten platform with 30x coverage (Kinghorn Centre for Clinical Genomics, Sydney). Bioinformatic processing was based on that used for the MGRB cohort[12]. Reads were aligned to the GRCh37 reference genome with Burrows-Wheeler Aligner[13] and SNVs and INDELs were called with the Genome Analysis Toolkit Best Practices pipeline[14]. All variants were annotated with Annovar[15] against RefSeq (version 01-06-2017). Principal component analysis using 17,453 SNVs and projection to the 1000 Genomes principal components were used to confirm ethnicity (akt v0.3.2). Non-relatedness of subjects in the cohort was confirmed with akt v0.3.2. Genotypes were considered missing if any of the following criteria were met: GQ < 30; DP < 10; DP > 150; heterozygous genotype with AB < 0.25 or AB > 0.75.

For VCCRI II, genomic DNA for cases was extracted from buccal cells collected using buccal swabs or mouthwash. Scope brand mouthwash was used for DNA collection. DNA was extracted using Purelink® Genomic Kit, Invitrogen, Australia for buccal swabs and Puregene

Core Kit A, Qiagen, USA for mouthwash. Control DNA extraction methods were described previously[16] (AF and DCM control samples). Samples were genotyped on the Axiom UK Biobank array (Kinghorn Centre for Clinical Genomics, Sydney). Genotyped case and control data were imputed using the MIS using the HRCr1.1 reference panel. Multiallelic SNPs and variants with gnomAD genome NFE <= 0.0001 were excluded. Non-relatedness of subjects in the cohort was confirmed with KING v2.1.4.

**Mayo Clinic case control study**

Study participants provided written informed consent under clinical and genetic research protocols approved by the Mayo Clinic Institutional Review Board (NCT01429727; NCT01427179).

Subjects were recruited from the Mayo Clinic patient population, including local residents, self- and physician-referred patients, and individuals who contacted investigators via the study website (www.mayo.edu/research/SCAD), and social media. Study subjects were consecutively enrolled in the Mayo Clinic SCAD registry after diagnostic confirmation of SCAD by review of coronary angiograms by an experienced interventional cardiologist. Demographic and clinical data were abstracted from questionnaires and medical records. A majority of individuals (95%) lived in the USA. FMD screening was performed in a subset of subjects by computed tomography angiography imaging of at least two arterial beds from brain to pelvis. Whole blood or saliva samples were obtained for genomic DNA extraction. Cases were genotyped for 713,599 SNPs by the Mayo Clinic Medical Genome Facility Genotyping Core using the Infinium OmniExpress-24 v1.2 BeadChip array (Illumina). Raw data were compiled using GenomeStudio software (Illumina) and exported to PLINK v1.9 for quality control (QC) and genome association analysis. Control genotypes were extracted from existing data files in the Mayo Genome Consortia, Center for Individualized Medicine, generated from Infinium HumanHap550, 610, 660 or OmniExpress BeadChip arrays (Illumina). QC filters excluded samples with a call rate <95%, sex-discordance, duplicates or cryptic evidence of relatedness using pairwise identical by descent estimates. To mitigate false-positive associations caused by population stratification, samples from individuals of non-Caucasian ancestry (<90% Caucasian) were detected using STRUCTURE software and excluded. Principal component analysis was used to examine the data for clustering due to platform and race. Exclusion criteria for genotyped SNPs were a call rate <95%, minor allele frequency <0.01, or deviation from Hardy-Weinberg equilibrium ($P < 1\times10^{-5}$). Following initial QC assessment, imputation was conducted separately for each platform, then combined to harmonize data. Imputation was performed on the University of Michigan Imputation Server using the Haplotype Reference Consortium reference panel. Imputed genotypes with a dosage $r^2$ of at least 0.7 and a minor allele frequency of at least 0.01 were used for analysis. Genome-wide association analysis of SNPs and SCAD risk were evaluated using logistic regression models assuming additive allele effects. To control for population stratification, the first 5 principal components were used as covariates in the models. Strength of association was estimated by calculating the odds ratio (OR) and corresponding 95% confidence interval (CI).

**CanSCAD/MGI (UBC/MGI) case control study**

Research ethics board approvals were obtained at each site of SCAD patient inclusion, and all patients provided informed consent for participation. GenSCAD study was approved by IRB approval: HUM00113268, SCAD Registry. Research ethics board approvals were obtained (IRB approval: HUM00112101), genetic analysis of arterial dysplasia and remodeling (MGI/AOS), and all patients provided informed consent for participation.

The CanSCAD genetic substudy (N=502) included SCAD patients from the prospective Canadian SCAD Cohort Study and the Non-Atherosclerotic Coronary Artery Disease (NACAD) Study. Patients presenting with acute SCAD were prospectively enrolled from 22 sites throughout North America (20 sites in Canada and 2 in the United States). SCAD diagnosis was confirmed on coronary angiography by the UBC core laboratory research team. Detailed baseline demographics, targeted history for predisposing conditions and precipitating stressors, and laboratory screening for predisposing conditions were performed. Screening for FMD was recommended for all SCAD patients, and multifocal FMD was defined according to consensus guidelines[3]. Patients were prospectively followed post-discharge at 1, 6, and 12 months, and annually thereafter for 3 years for cardiovascular (CV) events. Genetic studies were performed on CanSCAD patients who provided informed consent. Collection of DNA was obtained through blood or saliva self-collection kit (Oragene-500 kit, DNAGenotek). DNA was extracted according to the manufacturer's instruction (DNAGenotek) as previously described, and quantified using the Quant-iT PicoGreen assay (Life Technologies). DNA samples were normalized to a concentration of 50ng/µl for genotyping. The processed DNA samples were batched and transferred to the University of Michigan for GWAS analysis.

Due to the possibility of duplicate enrollment of individuals residing in the U.S. into multiple studies, particularly the Mayo clinic study which enrolled remote subjects from other institutions, this was accounted for in the current meta-analysis by removal of individuals from U.S. sites. This led to the removal of 49 individuals from the CanSCAD resource for the current analysis. By restricting to non-Finnish European individuals, 357 individuals were included in the final analysis.

The Michigan Genomics Initiative (MGI) is a program that recruited participants while awaiting diagnostic, interventional, and surgical procedures. Participants provided a blood sample for genetic analysis and agreed to link their sample to their electronic health record and other sources of health information. The current study's analyses involved 13,756 individuals from MGI genotyped with the same version (v1.1) of the Illumina BeadArray genotyping platform as the CanSCAD study SCAD cases at the University of Michigan DNA Sequencing Core Facility. Several ICD codes corresponding to diagnoses of as arterial diseases and connective tissue disorders were excluded. Controls were matched by age, sex, and ancestry through principal components of the GWAS data.

Genotyping of all CanSCAD, and 13,756 MGI samples, excluding samples with arterial diseases and connective tissue disorders based on ICD codes[17], were conducted by the University of Michigan DNA Sequencing Core using the Illumina Infinium HTS Assay Protocol, a semi-custom Infinium CoreExome-24v1.1 BeadArray with 607,778 SNP markers (UM_HUNT_Biobank_v1-1_20006200_A), and the Illumina GenomeStudio v2011.1. This GWAS+exome chip platform includes standard genome-wide tagging SNPs (N~240,000), exomic variants (n~280,000) and custom content from previously published GWASs, additional exonic variants selected from sequencing studies, ancestry informative variants and Neanderthal variants. Data Analysis Software package with Genotyping Module v1.9.4 and Illumina GenomeStudio (version 2.0) were used to cluster and call genotypes. Sample filtering was performed to exclude samples with call rate < 98%, estimated contamination > 2.5% (BAF regress), chromosomal missingness greater than 5 times other chromosomes, and sex mismatch between genotype-inferred sex and reported gender. Variant filtering was performed to exclude probes that could not be perfectly mapped to the human genome assembly (Genome Reference Consortium Human genome build 37 and revised Cambridge Reference Sequence of the human mitochondrial DNA; BLAT); Hardy-Weinberg equilibrium deviations in European ancestry samples (P<0.00001); variant call rate < 98%. Basic quality control (QC) filters including HWE P<0.000001, and variant missing call rate >2%, were implemented for each of our lab chip data and MGI chip data, before combining the two data sets.

We merged multiple batches of genotype data of SCAD cases and then applied pre-quality control using the HRC Imputation preparation and checking tool by the McCarthy Group before merging them with MGI genotyped data, which applied the same pre-HRC-imputation QC. It compares each of our individual genotyped data with HRC reference, and corrects the variants strand-flip as well as aligns the allele codes with HRC reference alleles. It also removes A/T & G/C SNPs if MAF > 0.4, SNPs with differing alleles, SNPs with > 0.15 allele frequency difference, and SNPs not in reference panel. After this process, 351,487 polymorphic variants remained (chr1-23). The total genotyping rate was 0.99. Based on the genotyping result, we excluded 1 sample with missing call rate >2%, 1 duplicate or close relatives based on whole genome genotyping data identity-by-descent (IBD) >0.35, 2 gender mismatched samples, 2 genetic syndrome cases, and 2 are further confirmed not SCAD cases. There were no samples failing QC due to checks of inbreeding coefficient. For the merged data, we further confirmed that none of the SCAD cases and MGI controls were duplicates or overlapped based on the IBD analysis.

We then imputed autosomal chromosome genotypes of the Haplotype Reference Consortium (HRC) using the Michigan Imputation Server on the 13,756 MGI and all the UBC SCAD samples. The parameters for imputation included: 1) Minimac4 method; 2) HRC r1.1 2016 reference panel; 3) Eagle v2.3 as phase output; 4) EUR as quality control population. We filtered poorly imputed variants (R2<0.8) and rare variants (MAF <1%). We also excluded SNPs with potential frequency mismatches comparing with reference panel (markers with Chi-squared greater than 300). 6,690,240 imputed variants (chromosomes 1-22) remained after filtering. The correlation $r^2$ between the ref allele frequency of our samples and the HRC reference panel was 0.999. For the matched control sample selected from 13756 MGI control pool for each case, we required the control to have the same gender, close birth years, and close ancestries as the case. We expected that every case was matched to at least one control. We took an approach that we searched from +/-5 years (5-year window) in age first, followed by +/-10 years instead, and so on to +/-30 years. We stopped searching once there was at least one control selected. From the possible controls in the applicable sex and age category, we chose the best ethnic match for each case that had the smallest principal components distance (via the top 3 PCs computed from the TRACE program). To guarantee every case could match to 6 controls, we repeated the entire procedure 6 times. Furthermore, in our final GWAS, we only focused on non-Finish European subjects, based on the criteria of within mean +/- 6 SD region of PC1 and PC2 from 1000 Genome reference. 2125 matched MGI controls were selected for 357 SCAD case samples. We finally tested the single genetic imputed variants using the PLINK program, with the first five principal components as covariates.

**DEFINE-SCAD case control study**

The DEFINE study (NCT01967511) protocol is approved by the Human Research Ethics Committee of the Icahn School of Medicine at Mount Sinai[18]. Patients with suspected arteriopathies were seen and assessed in the Mount Sinai Vascular Medicine Clinic. Inclusion criteria for entry into DEFINE included >=18 years of age, being freely willing to participate and fluency in English. FMD cases were required to have a clinical diagnosis of multifocal FMD

confirmed by imaging [computed tomographic angiography, magnetic resonance angiography, or catheter-based angiography]. SCAD patients were required to have a clinical diagnosis of SCAD confirmed by invasive coronary angiography. Initially only females were permitted to enter the study, but from early 2017 both males and females were enrolled. For healthy controls, inclusion criteria included no clinical features of FMD, cervical artery dissection, or SCAD (including no cervical or abdominal bruits, an absence of family history of sudden death or aneurysm) and absence of any major ongoing systemic disease including any condition requiring hospitalization, immune suppression, intravenous or injected medications or that result in functional impairment in the performance of activities of daily living. Healthy controls were recruited from the general population and were pre-screened by the same clinical team and matched to study cases according to age, sex, race/ethnicity, and body mass index (BMI). Exclusion criteria (for cases and controls) included: co-morbidities which reduce life expectancy to 1 year; any solid organ or haematological transplantation, or those in whom transplantation is considered; active autoimmune disease; illicit drug use; HIV positive; prior malignancy. In controls, an additional exclusion criterion was an early-onset family history of any form of vascular disease. Healthy controls also underwent screening clinical assessment, with specific attention paid to any history or physical examination findings suggestive of FMD or other vascular disease, by two clinical experts in FMD (J.W.O. and D.K.-D.).
If the entry criteria are met and following informed consent, blood draw and skin punch-biopsy (from the medial upper arm) were performed. At the blood draw, 10mL of blood were collected into ethylenediaminetetraacetic acid (EDTA) tubes and reserved for deoxyribonucleic acid (DNA) extraction. DNA was isolated from whole blood using the Puregene Blood Core kit B (cat# 158467, Qiagen, Germantown, MD, USA). DNA was aliquoted and frozen at -80°C. Of 384 total DNA samples processed, 336 samples were processed using Illumina-Human-Omni-Express-Exome genotyping array and 48 samples were processed using Infinium OmniExpressExome-8v1.6 array, which included additional CAUSE and DEFINE study subjects not included in this analysis. All samples showed a call rate > 99% and were thus retained. No genetically identical pairs were found, no outliers were identified in a Principal Components Analysis (run using eigenstrat), and the sex of all samples was correctly predicted for each subject. Further inspection of the samples' heterozygosity revealed that there were six subjects who did not pass the QC and were excluded. Genotyping array probes were filtered according to call rate (> 95%) and Hardy-Weinberg equilibrium test P value (> $1x10^{-6}$). Data from Illumina-Human-Omni-Express-Exome genotyping array (336 subjects) and Infinium OmniExpressExome-8v1.6 array (48 subjects) yielded 962,157 shared variants after QC. Genotyping data was imputed using the HRC (haplotype research consortium, http://www.haplotype-reference-consortium.org/) reference and the Michigan Imputation Server, using the MACH/minimac imputation pipeline. We successfully imputed 16,229,543 variants.

#### Funding and acknowledgements

##### DISCO-3C case control study

The DISCO study was supported by the European Research Council grant (ERC-Stg-ROSALIND-716628), the French Society of Cardiology foundations “Coeur et Recherche” and "La Fédération Française de Cardiologie", and the French Coronary Atheroma and Interventional Cardiology Group (GACI). The French study acknowledge the Spanish National Cancer Research Centre, in the Human Genotyping lab, a member of CeGen Biomolecular resources platform (PRB3), to be supported by grant PT17 /0019, of the PE I+D+i 2013-2016, funded by Instituto de Salud Carlos III and a European regional development fund (ERDF). The genotyping of the controls from the Three-City Study (3C) was supported by the non-profit organization Fondation Alzheimer (Paris, France) to PA. We acknowledge all clinicians and patients who contributed to the DISCO register, the French Society of Cardiology and the French Coronary Atheroma and Interventional Cardiology Group for their support, the Clinical Research Associates of the Clermont-Ferrand University Hospital: Elodie Chazot, Carole Bellanger, Laurie Cubizolles, Aurélie Thalamy, Ouarda Lamallem. We acknowledge the Spanish National Cancer Research Centre, in the Human Genotyping lab, a member of CeGen where genotyping was performed for DISCO patients and the European Global Screening Array Consortium.

**SCAD-UK case-control studies**
We thank AstraZeneca's Centre for Genomics Research, Discovery Sciences, BioPharmaceuticals R&D for funding the sequencing of and providing the bioinformatics support related to subjects in cohort SCAD-UK I. The study is supported by the British Heart Foundation (BHF) PG/13/96/30608, the National Institute for Health Research (NIHR) rare disease translational collaboration, the Leicester NIHR Biomedical Research Centre and BeatSCAD. Dr Webb is funded by the British Heart Foundation (SP/16/4/32697). We are grateful for the support of SCAD-survivors and our clinical colleagues throughout the UK and the leadership of the ESC-ACCA SCAD Study Group. We specifically acknowledge the support of Jenny Middleton, Jane Plume, Donna Alexander, Daniel Lawday and Andrea Marshall for all their support for SCAD research. We thank the AstraZeneca Centre for Genomics Research Analytics and Informatics team for processing and analysis of sequencing data.

**VCCRI case control studies**
The VCCRI study was supported by Cardiac Society of Australia and New Zealand (CSANZ) Cardiovascular Research Innovation Grant (RMG, DWMM, DF, JCK), National Health and Medical Research Council (NHMRC), Australia (APP1161200), NSW Health Early Mid-Career Cardiovascular Grant (EG), NSW Health Senior Clinician Cardiovascular Grant (RMG). The authors would like to thank the SCAD patients for their enthusiastic participation in this study, Sarah Ford and Pamela McKenzie and SCAD Research Inc, our colleagues who referred SCAD cases, and the Medical Genome Reference Bank including the 45 and Up and ASPREE study patients who were controls for this study. The authors would like to thank Claire MY Wong, Ketan Mishra, and Renee Johnson for their contributions to data collection and sample processing.

**Mayo Clinic case control study**
The Mayo Clinic study was supported by funding from SCAD Research Inc, National Institutes of Health (NIH: T32 GM72474), and resources from the Genome Consortia, Mayo Clinic Center for Individualized Medicine. The authors thank our clinical colleagues who referred SCAD cases and the patients who supported this research.

**CanSCAD/MGI (UBC/MGI) case control study**
The U.S. site of the CanSCAD/MGI study funding support included grants from National Institutes of Health (R01HL139672, R35HL161016), Heart and Stroke Foundation of Canada (G-17-0016340), Canadian Institutes of Health Research (grant #136799), and the University

R35HL161016, R01HL086694, Department of Defense, and the A. Alfred Taubman Institute. J.S. and L.R.B are supported by a Michael Smith Foundation for Health Research Scholar award. L.R.B is a Canada Research Chair in Precision Cardiovascular Disease Prevention. The University of Michigan Advanced Genomics Core performed genotyping of all CanSCAD and MGI samples. The authors acknowledge the University of Michigan Precision Health Initiative and Medical School Central Biorepository for providing biospecimen storage, management, processing and distribution services and the Center for Statistical Genetics in the Department of Biostatistics at the School of Public Health for genotype data management in support of this research. We thank all the participants of the studies included in the analyses. We acknowledge the FMD Society of America and Vancouver SCAD Conference organizers for enabling study enrolments at patient meetings.

**DEFINE-SCAD case control study**
DEFINE is supported by the US National Institutes of Health (R01HL148167) and we acknowledge the many philanthropic supporters of DEFINE who from the outset have enabled and facilitated this study. JCK acknowledges research funding from, New South Wales health grant RG194194, the Bourne Foundation and Agilent.
We acknowledge and thank all the subjects, both cases and controls, in the DEFINE study. We also thank all our clinical colleagues who referred SCAD, FMD and CeAD patients to this study.

**Xia Yang lab**
We acknowledge funding from the NIH/NHLBI R01HL147883 for XY, and American Heart Association Predoctoral Fellowship 829009 and UCLA Integrative Biology and Physiology Edith Hyde Fellowship for MB.

Supplementary Figure 1

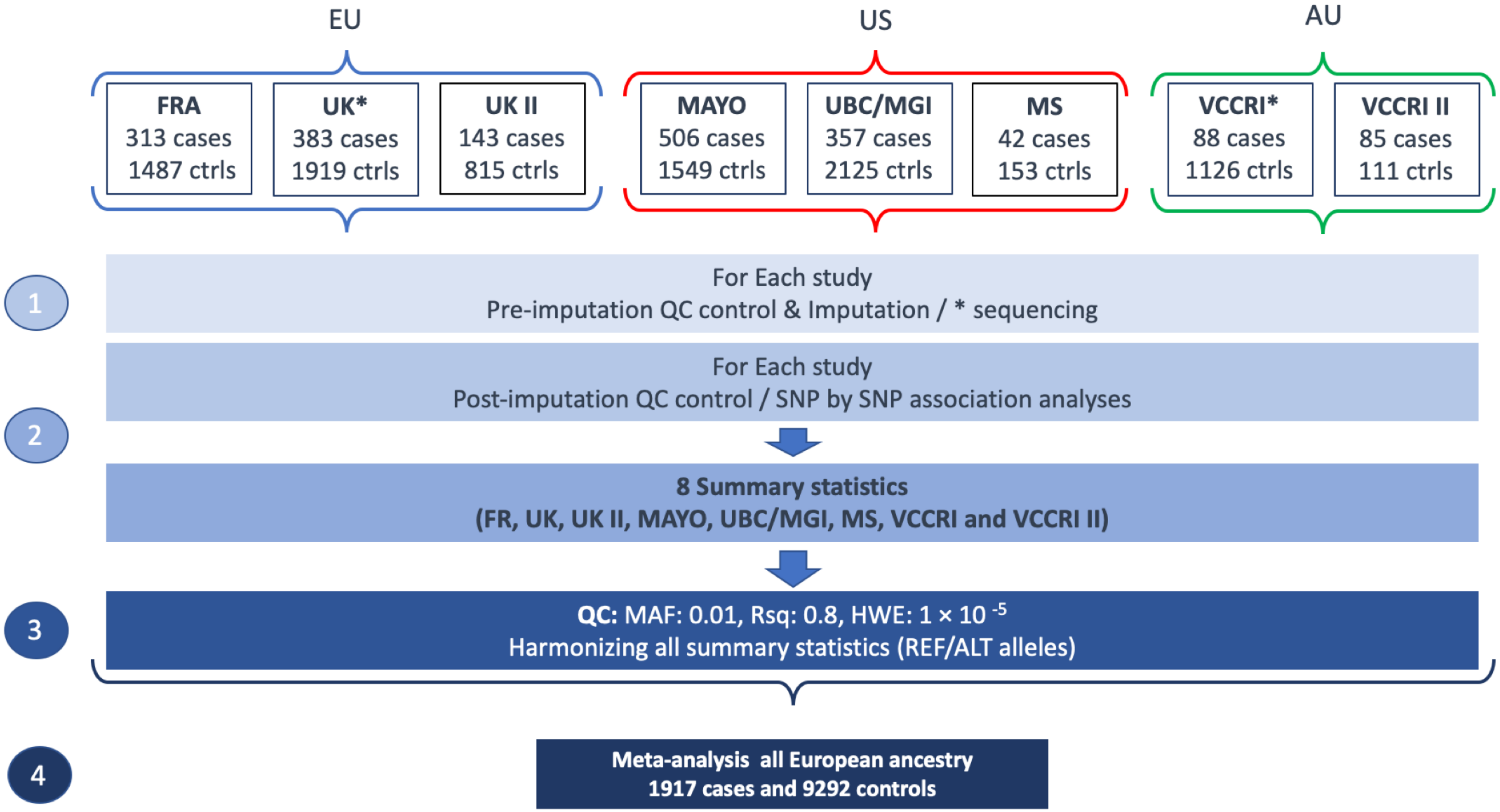

**Supplementary Figure 1: GWAS meta-analysis design**

#### Supplementary Figure 2

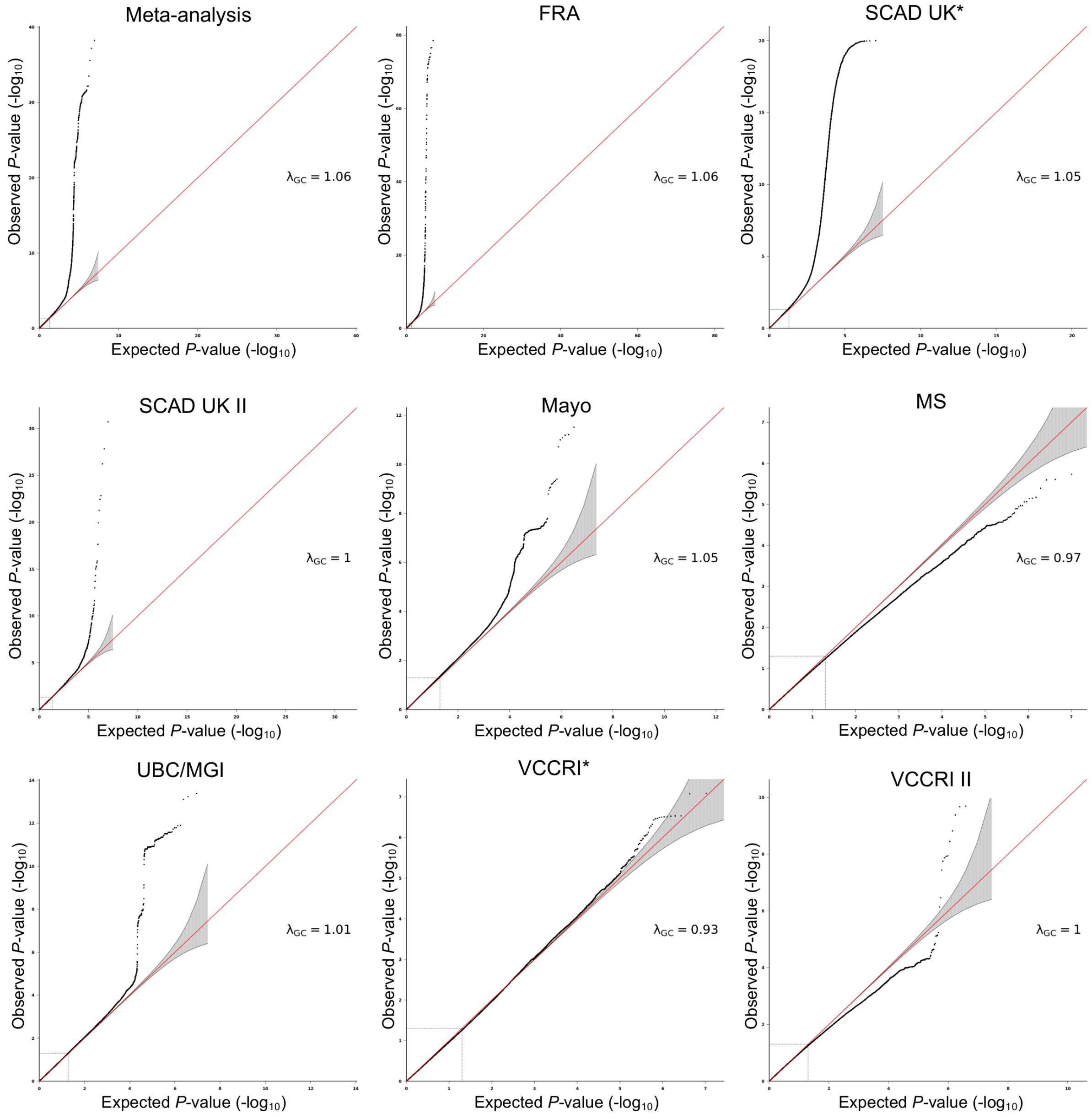

**Supplementary Figure 2. Quantile-quantile plot representation of SNP-based association analysis in individual studies.**

$-\log_{10}$ of observed association *P*-value value is represented on the y-axis, expected *P*-value on the x-axis. Genomic control value ($\lambda$GC) is indicated for each study.

Supplementary Figure 3

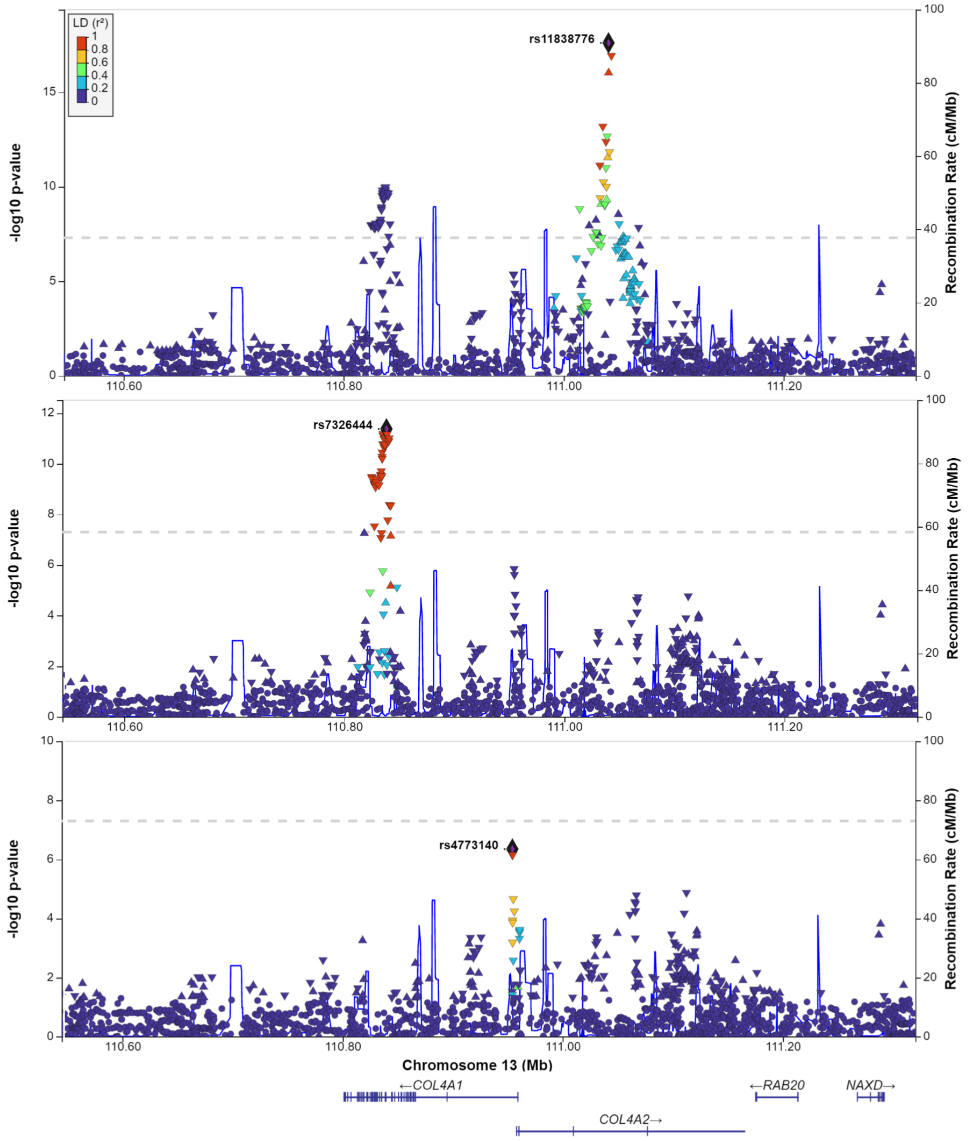

**Supplementary Figure 3. Conditional analysis showing independence of COL4A1/COL4A2 genetic signals.** LocusZoom plots represent SCAD association at COL4A1/COL4A2 locus (chromosome 13) before conditioning (A), after conditioning on rs11838776 (B) and after conditioning on rs11838776 and rs7326444 (C). No variant with genome-wide significant association remains after conditioning on these two SNPs, although other suggestive signals are present at the locus. Dot shape indicate the lead SNP (diamond shape), and Dot shape indicate the lead SNP (diamond shape) and effect size (β) of nominally significant variants (upper triangle: β>0, lower triangle, β<-0, round shape: p-value>0.05). Dot-color indicates linkage disequilibrium ($r^2$) with the lead SNP at each locus in the European subset of 1000G reference panel.

Supplementary Figure 4

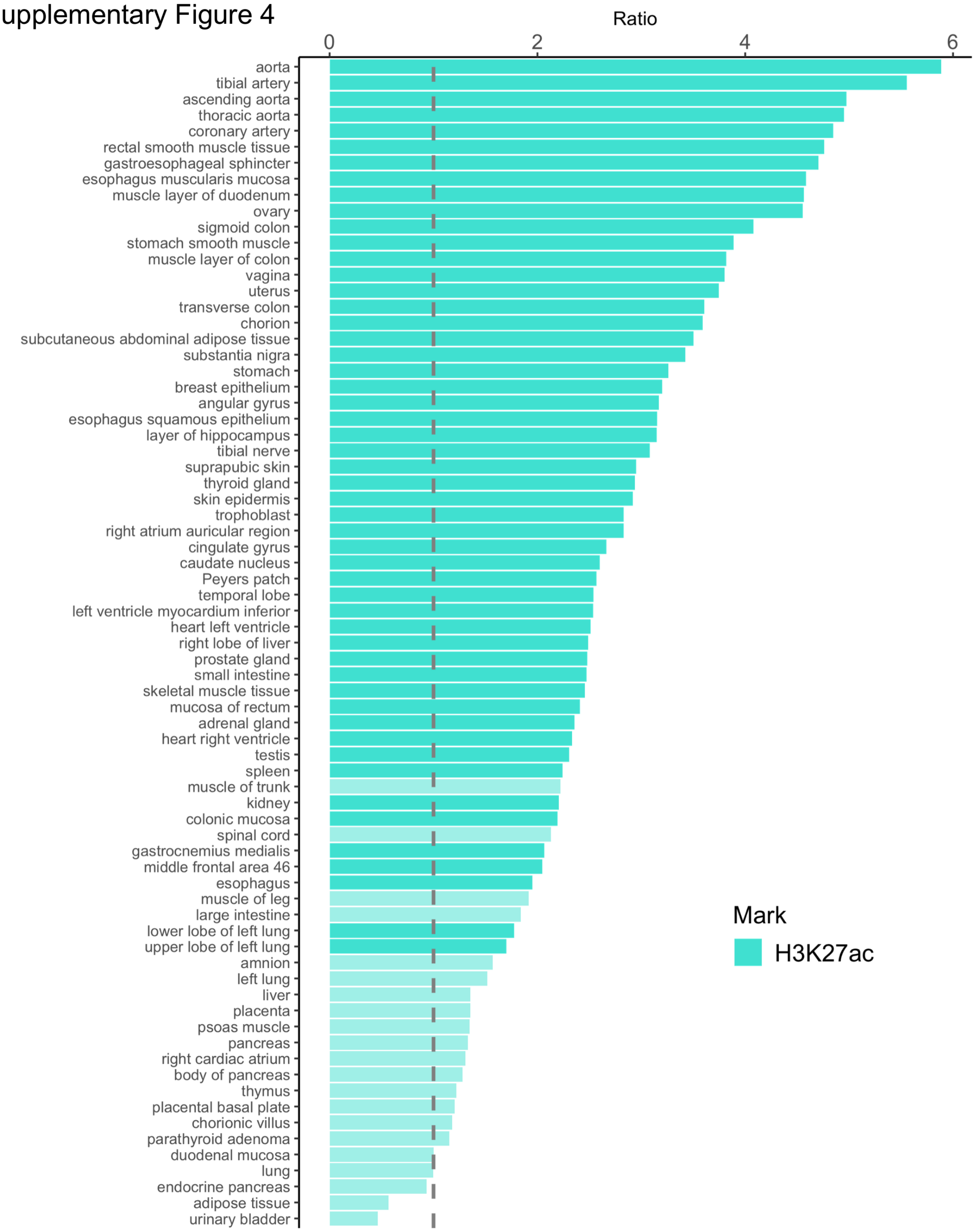

**Supplementary Figure 4. Representation of SCAD SNPs enrichment open chromatin regions of 73 tissue with available H3K27ac ChIP-Seq experiments in ENCODE database.** For each tissue, H3K27ac narrowpeak files of all experiments were merged to generate one dataset. A random background was created for each H3K27ac peak file by randomly selecting 1000 sets of size and chromosome matched regions from the genome. Enrichment represents the ratio of the number of SCAD SNPs overlapping H3K27ac peaks over the average number of SCAD SNPs overlapping background regions. *P*-value was evaluated using a binomial test with greater enrichment as alternative hypothesis, and is indicated by a brighter color for significant enrichment (Bonferroni adjusted *P*-value <0.05)

### Supplementary Figure 5

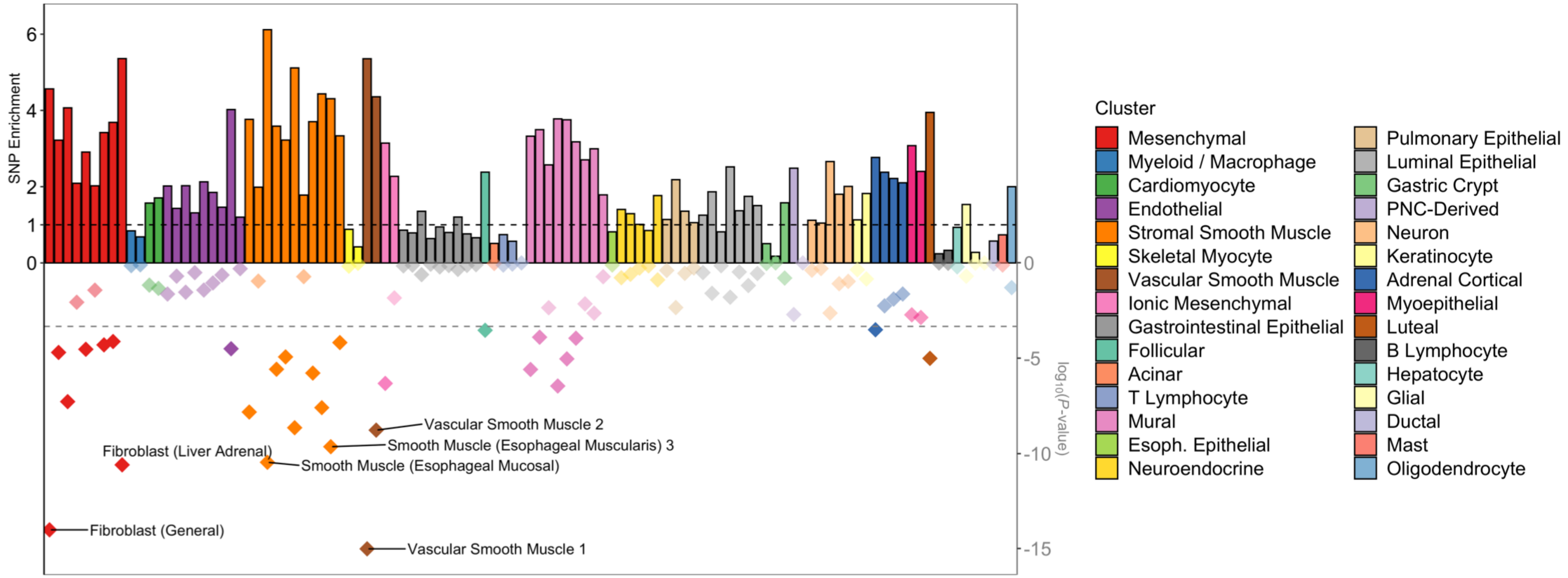

**Supplementary Figure 5. Enrichment of SCAD SNPs in open chromatin regions from human adult cells.** Representation of SCAD SNPs fold-enrichment (upper y-axis) and enrichment *P*-value (log scale, lower y-axis) among open chromatin regions of 105 sub clusters determined from single-nuclei ATAC-Seq in 30 adult tissues[19]. A random background was created for each open chromatin dataset by randomly selecting 1000 sets of size and chromosome matched regions from the genome. Enrichment represents the ratio of the number of SCAD SNPs overlapping open chromatin regions over the average number of SCAD SNPs overlapping background regions. *P*-value was evaluated using a binomial test with greater enrichment as alternative hypothesis. Bar colour represents the main cell clusters which are defined in the legend. The names of the 5% sub clusters with lower P-value for SCAD SNPs enrichment are indicated over the graph.

### Supplementary Figure 6

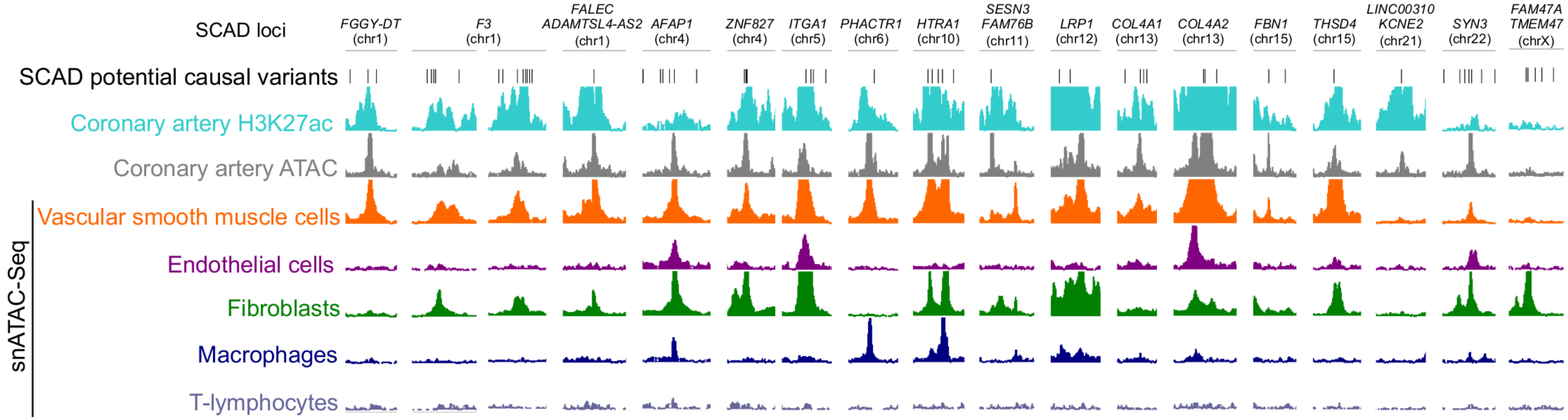

**Supplementary Figure 6. Overlap of SCAD associated variants with chromatin marks for regulatory regions.** Genome browser visualization of H3K27ac ChIP/ATAC-Seq/snATAC-Seq read densities (in reads/million, r.p.m.) in the regions surrounding putative SCAD causal variants. snATAC-Seq density profiles correspond to sub clusters representing more than 1% of cells in artery tissue, and grouped by main cell type cluster[19]. A complete list of SCAD variants overlapping functional regions in artery tissue is available in supplementary file.

Supplementary Figure 7

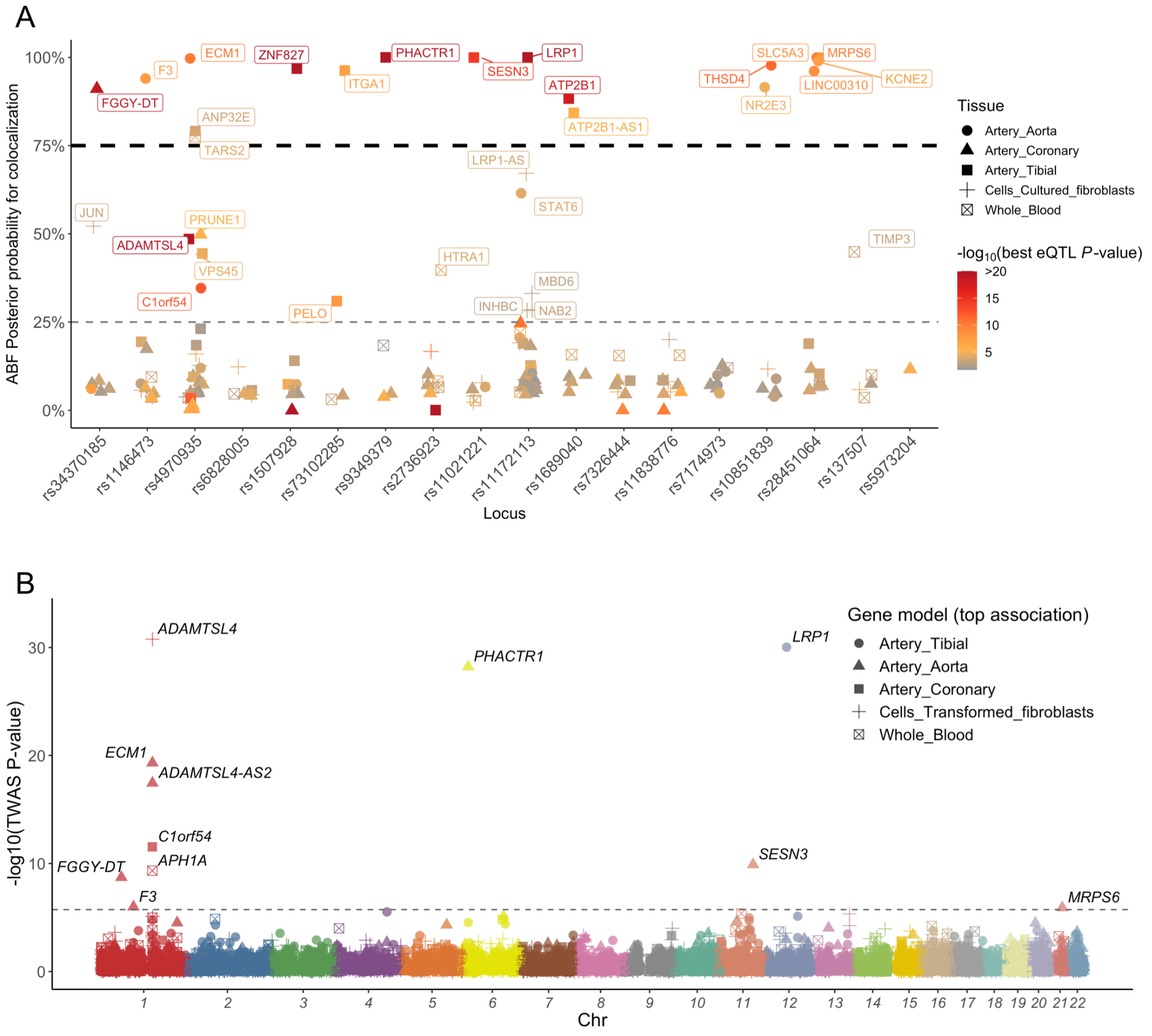

**Supplementary Figure 7. Integration of SCAD genetic association with expression of potential target genes in relevant tissues.**

**A:** Colocalization of eQTL and GWAS association at SCAD loci. Colocalization was assessed for all genes located within 500kb of SCAD top SNPs in five relevant tissues from GTEx (v8 release). *y*-axis represents the approximate Bayes factor posterior probability for eQTL and GWAs association to share one common variant at the locus. Dot color represents the *P*-value of top eQTL association involving variants at the locus. Only the tissue with highest probability for colocalization is represented for each gene, and indicated by the dot shape. Genes are grouped per SCAD lead SNP ranked by genomic position along *x*-axis. Names of genes with colocalization probability > 25% are indicated **B:** Manhattan plot representation of Transcriptome-wide association analysis (TWAS) in SCAD with gene expression models computed in 5 tissues based on GTEx (v7 release). −log10 of association *P*-value is represented on the *y*-axis, genomic coordinates on the x-axis. Name of genes with Bonferroni corrected *P*-value < 0.05 are indicated.

Supplementary Figure 8

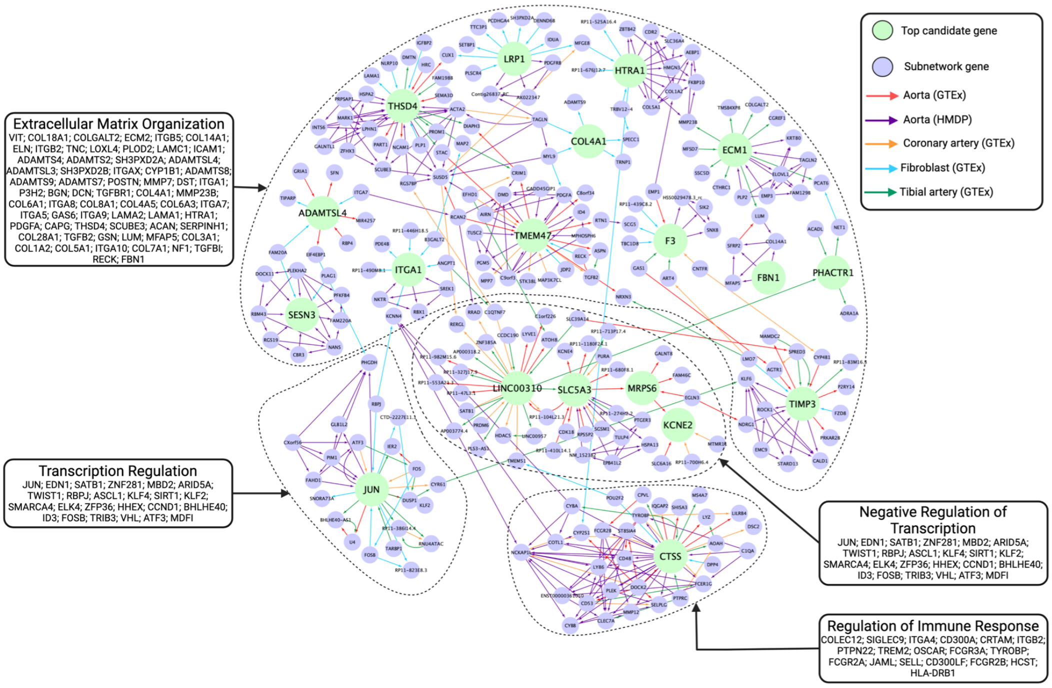

**Supplementary Figure 8. Bayesian networks constructed using gene expression of top candidate genes in SCAD risk loci.**

Tissue-specific Bayesian networks (BN) were constructed from the Genotype-Tissue Expression project Version 8 and Hybrid Mouse Diversity Panel gene expression data via RIMBANET. The BNs representing tissues and cell types relevant to SCAD onset were queried for the top GWAS hits. The top GWAS hits are emphasized by their larger size and color (green), whereas their surrounding subnetwork gene are shown in a smaller size and contrasting color (purple). The network edges inform on the source of the connection and the direction of regulation. Suggested biological pathways in which denoted subnetworks function as well as the genes involved in the pathway are indicated

Supplementary Figure 9

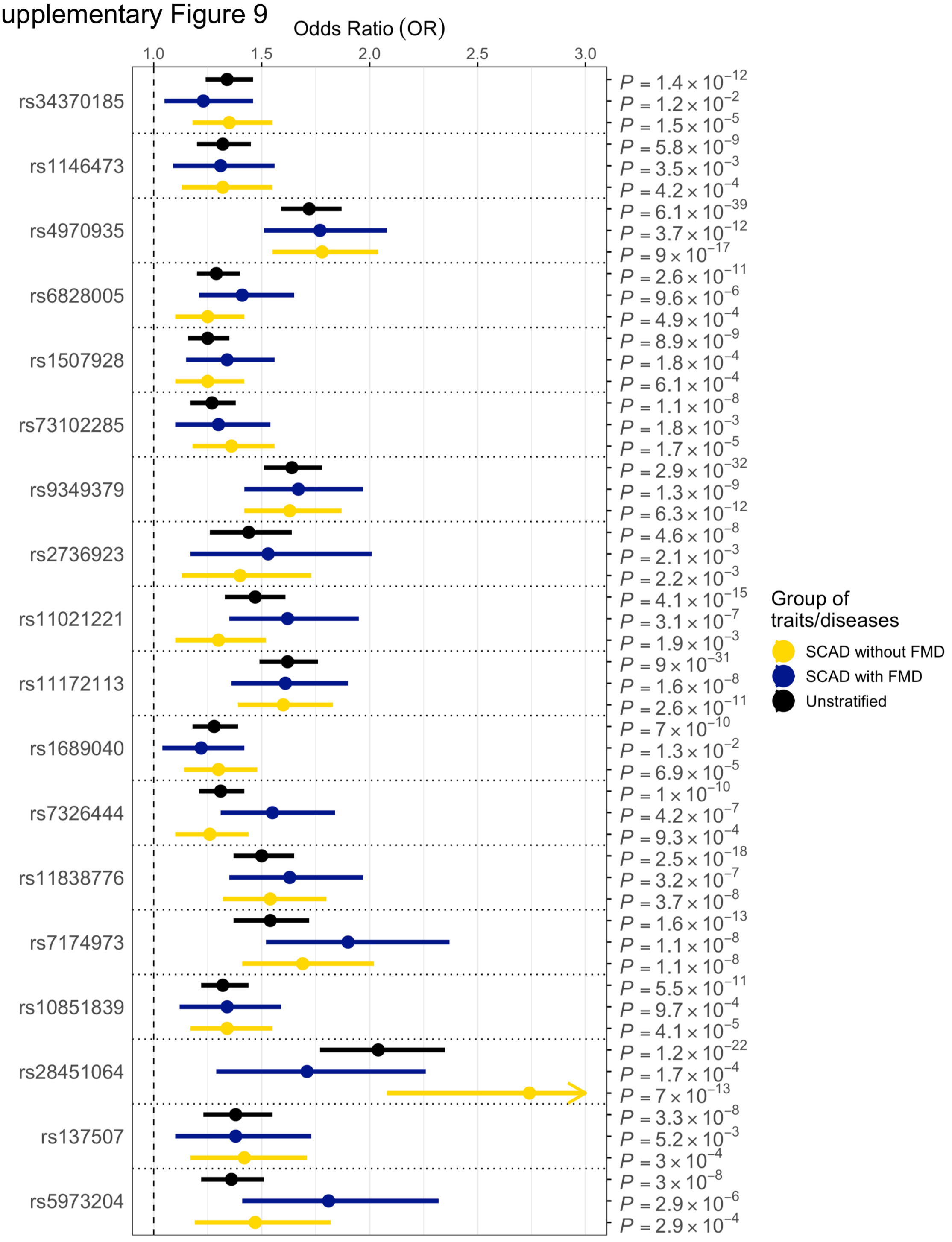

**Supplementary Figure 9. Association of lead SNPs with SCAD according to FMD status.** Forest plot representing the association of each SCAD top SNP in the general meta-analysis with 8 studies (black) as well as in the FMD stratified meta-analyses, FMD+ (blue) and FMD- (yellow) with 4 studies each. Odds Ratio of the association is represented on the x-axis and range represents the 95 % confidence interval. Each association's P-value is indicated with the trait on the y-axis.

Supplementary Figure 10

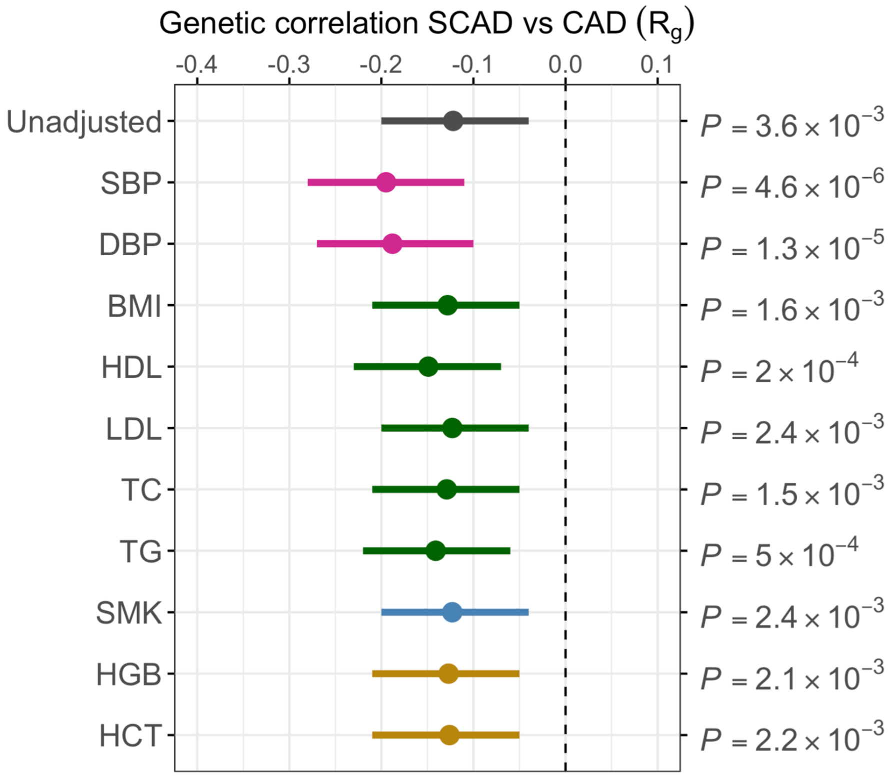

**Supplementary Figure 10**. **Forest plot representing the genetic correlation between SCAD and CAD unadjusted and after mtCOJO conditioning on cardiometabolic traits**. Before re-estimation, SCAD genetic association statistics were conditioned on systolic blood pressure (SBP), Diastolic blood pressure (DBP), Body mass index (BMI), High-density lipoprotein (HDL), Low-density lipoprotein (LDL), Total cholesterol (TC), Triglycerides (TG), Smoking (SMK), Haemoglobin (HGB) and Haematocrit (HCT). Rho coefficient of genetic correlation (Rg) is represented on the x-axis and range represents the 95 % CI. Undajusted *P*-value of correlation is indicated.

### Supplementary Tables

**Supplementary Table 1: Clinical characteristics of the study populations**
BMI: Body-Mass Index, HTN: Hypertension, n: count, Q1: 25% quantile, Q3: 75% quantile

| Study | French Study | | SCAD-UK Study I | | SCAD-UK Study II | | Mayo Clinic Study | |
|---|---|---|---|---|---|---|---|---|
| | Cases (DISCO) | Controls (3C-Study) | Cases | Controls | Cases | Controls | Cases | Controls |
| Type | Clinical based | Population based | Clinical based | Population based | Clinical based | Population based | Clinical based | Healthy volunteers |
| Inclusion criteria | age> 18,<br>1) retrospective with a diagnostic of SCAD made from 2010, or 2) prospective at the time of hospitalisation during which the diagnosis of SCAD was made. | Geographic sampling | SCAD confirmed on invasive angiography | | SCAD confirmed on invasive angiography | | SCAD confirmed by angiogram | No reported SCAD |
| Exclusion criteria | Age <18; atherosclerotic ischemic disease; iatrogenic hematoma | Age < 65y | Atherosclerotic dissection, iatrogenic dissection | | Atherosclerotic dissection, iatrogenic dissection | | Diagnosis of connective tissue disorder or aortopathy; iatrogenic | Diagnosis of atherosclerotic coronary artery disease, acute myocardial infarction, FMD, arterial aneurysm or dissection, cerebral infarction, Marfan syndrome, Ehlers-Danlos syndrome |
| Total (n) | 313 | 1487 | 383 | 1925 | 139 | 815 | 506 | 1549 |
| Women (n,%) | 285 (91) | 876 (58.9) | 361 (94.2) | 1815 (94.3) | 115 (82.7) | 665 (81.6) | 484 | 1477 |
| Age at study inclusion (Median, Q1,Q3) | 51, 44, 59 | 74.36 ±5.5<br>[65 - 94] | | 56,49,62 | | 56, 48, 61 | 46.6 ±9.2 | 64 ±14.5 |
| Age at SCAD (Median, Q1/Q3) | 52.2, 44.55, 60 | NR | 47, 41, 52 | NR | 49.0, 43, 54 | NR | 46.6, 39, 53 | NR |
| SCAD Type (1, 2, 3) | 49, 237, 32 | NR | | NR | | NR | Single vessel-397;<br>Multi-vessel-87 | NR |
| FMD (Yes, No, NA) | 140, 152, 21 | NR | 104,108,171 | | 20,71,48 | | 175, 140, 169 | unknown |
| BMI (kg/m2) | **58** | NA | median 25 (23,29)<br>6 missing values | | 27 (23,31)<br>1 missing value) | NA | 26.0 ±5.9 | unknown |
| HTN (n,%) | 96 (30.7) | 1171 (78.7) | 94 (24.6; 1 missing value) | | 25 (18.0) | NA | 157 (32.4) | unknown |
| T2D (n,%) | 12 (3.2) | | 7 (1.8) | | 2 (1.4%) | NA | 14 (2.9) | unknown |
| Migraine (n,%) | 76 (28.4) | | 176 (48,9; 23 missing values) | | 67 (48.2; 2 missing values) | NA | 175 (36.2) | unknown |
| Smoking (Non/Current/Ex) | Ever<br>220/93 | | 266/14/103 | | 94/9/36<br>(67, 6, 26) | NA | 343/ 12 /129<br>(71 /2.5/26.7) | unknown |

| Study | CanSCAD/MGI Study | | DEFINE-SCAD Study | | VCCRI Study I | | VCCRI Study II | |
|---|---|---|---|---|---|---|---|---|
| | Cases | Controls | Cases | Controls | Cases | Controls | Cases | Controls |
| **Type** | Clinical based | Population based | Clinical based | Clinical based | Clinical based | Population based | Clinical based | Clinical Based |
| **Inclusion criteria** | SCAD diagnosis was confirmed on coronary angiography by the UBC core laboratory research team, and categorized according to previously established Saw classification | Age, Sex, PC (PC1-PC3) matched controls | SCAD confirmed on invasive angio | Vascular disease excluded on history and physical exam. Also matched to SCAD cases by age, BMI, sex. | SCAD confirmed by angiogram | No reported SCAD | SCAD confirmed by angiogram | No Reported SCAD |
| **Exclusion criteria** | Angiogram unavailable or did not appear to be SCAD; from N=502, only Canadian samples consistent with 1000G non-Finish European ancestry (+/- 6 SD of PC1 and PC2) were retained for analysis. | Of 13,756 MGI samples eligible for the study after exclusion of vascular or connective tissue diagnoses, and matching for age, sex and ancestry (based upon genetic PC's) 2,125 matched MGI controls were retained for analysis. | Age < 18, diagnosis of connective tissue disorder or aortopathy; iatrogenic. Plus several other exclusions (HIV+, active malignancy) | Multiple - mostly relate to exclusion of vascular disease and other things like HIV+, active malignancy) | Angiogram unavailable or did not appear to be SCAD | No reported history of cancer, cardiovascular disease or neurodegenerative diseases before 70 years old | Angiogram unavailable or did not appear to be SCAD | Related to other sample |
| **Total (n)** | 357 | 2125 | 42 | 153 | 88 | 1127 | 85 | 111 |
| **Women (n,%)** | 315 (88.2%) | 1873 (88.1%) | 41 (97.6%) | 153 (100%) | 80, 90.9% | 672, 59.6% | 83, 97.6% | 46, 41.4% |
| **Age at study inclusion (Median, Q1,Q3)** | 53, 46, 60 | 53, 46, 61 | 49, 41.5, 53.75 | 50 (43-58) | 50, 44, 59 | all >70 years old | 52, 48, 60 | 61, 52, 67 |
| **Age at SCAD (Median, Q1/Q3)** | | NR | 45.5, 36, 50.25<br>6 missing values | NA | 44, 39, 52 | NA | 49, 43, 56 | NA |
| **SCAD Type (1, 2, 3)** | 117,193,36 | NR | 4; 32; 1; NA: 5 | NR | 32, 50, 4, (2 NR) | NA | 29, 48, 6 | NA |
| **FMD (Yes, No, NA)** | 149,123,85 | NR | 31, 10, 1 | NR | 14, 32, 42 | NR | 10, 22, 53 | NR |
| **BMI (kg/m2)** | median 25.5 (22,30;9 missing values) | NA | 24.55 (22, 29.15) | 23.85 (20.98, 26.6) | 26.32 | 27.5 | 27.04 | NR |
| **HTN (n,%)** | 108 ( 30%); 33 missing values | NA | 18 (42.86%) | 4 (2.61%) | 17, 19% | NR | 19, 22.3% | NR |
| **T2D (n,%)** | 9 (2.5%); 33 missing values | NA | 0 (0%) | 1 (0.65%) | 3, 3.4% | NR | 6, 7% | NR |
| **Migraine (n,%)** | 100 (28%); 33 missing values | NA | 9 (21.4%) | NA | 41, 46.6% | NR | 48, 56.5% | NR |
| **Smoking (Non/Current/Ex)** | 211/25/88 | NA | 34/1/7 | NA | 54/3/28 | NR | 51/3/31 | NR |

**Supplementary Table 2: SNP heritability estimates**

Values are given on the observed scale (h2obs) and liability scale (h2liab).
Prevalence used for conversion to the liability scale is shown.
Effective number samples were used for the conversion, Neff = 4/(1/Ncases+1/NControls).
For SumHer, two analyses were done: one assuming LDAK-THIN model, using a pre-calculated tagging file obtained by using LD reference data from UKBB, and one to mimic LDSC, with the same settings and reference panel (HapMap3 and 404 nonFin european from 1000G project).
Neff, effective sample size.

| Trait | Method | *h* 2obs (s.e.) | *intercept (s.e.)* | Prevalence | *h* 2liab (s.e.) | Cases | Controls | Neff |
|---|---|---|---|---|---|---|---|---|
| SCAD | LDSC | 0.73 (0.11) | 1.016 (0.0072) | 0.01 | 0.71 (0.11) | 1917 | 9292 | 6357 |
| SCAD | SumHer (LDSC) | 0.77 (0.10) | 1.020 (0.0079) | 0.01 | 0.75 (0.10) | 1917 | 9292 | 6357 |
| SCAD | SumHer (LDAK-THIN) | 0.71 (0.12) | 1.027 (0.0088) | 0.01 | 0.70 (0.12) | 1917 | 9292 | 6357 |

**Suplementary Table 3: SNP heritability of the top loci region (1 Mb)**

The table shows local heritability estimation, p-values obtained from partitionned heritability using SumHer LOCUS: region of 1 Mb named on lead SNP (GWAS hit), CHROM: chromosome,START: position of lead SNP - 500k bp, STOP: position of lead SNP + 500k bp,EA: effect allele, h2obs: observed heritability estimation, SD: standard deviation of h2obs

| **GENE** | **LOCUS** | **CHROM** | **START** | **STOP** | **h2obs** | **SD** |
|---|---|---|---|---|---|---|
| *FGGY-DT* | rs34370185 | 1 | 59156909 | 60156909 | 0.013 | 0.005 |
| *F3* | rs1146473 | 1 | 94550472 | 95550472 | 0.009 | 0.004 |
| *FALEC/ADAMTSL4-AS2* | rs4970935 | 1 | 150004062 | 151004062 | 0.028 | 0.012 |
| *AFAP1* | rs6828005 | 4 | 7274352 | 8274352 | 0.012 | 0.004 |
| *ZNF827* | rs1507928 | 4 | 146288035 | 147288035 | 0.005 | 0.003 |
| *ITGA1* | rs73102285 | 5 | 51655642 | 52655642 | 0.011 | 0.005 |
| *PHACTR1* | rs9349379 | 6 | 12403957 | 13403957 | 0.012 | 0.004 |
| *HTRA1* | rs2736923 | 10 | 123759062 | 124759062 | 0.012 | 0.004 |
| *SESN3/FAM76B* | rs11021221 | 11 | 94808854 | 95808854 | 0.012 | 0.005 |
| *LRP1* | rs11172113 | 12 | 57027283 | 58027283 | 0.019 | 0.007 |
| *ATP2B1* | rs1689040 | 12 | 89478233 | 90478233 | 0.008 | 0.004 |
| *COL4A1* | rs7326444 | 13 | 110338236 | 111540681 | 0.022 | 0.006 |
| *COL4A2* | rs11838776 | 13 | | | | |
| *FBN1* | rs7174973 | 15 | 48263754 | 49263754 | 0.003 | 0.002 |
| *THSD4* | rs10851839 | 15 | 71128370 | 72128370 | 0.007 | 0.003 |
| *LINC00310/KCNE2* | rs28451064 | 21 | 35093827 | 36093827 | 0.013 | 0.005 |
| *SYN3* | rs137507 | 22 | 32782971 | 33782971 | 0.007 | 0.003 |
| **all 16 loci (all SNPs)** | - | - | - | - | 0.188 | 0.018 |

**Supplementary Table 4: Annotation of candidate functional SNPs**

We display SNPs previously identified as potential functional SNPs (LD r > 0.7 with the lead SNPs or 95% credible set) and overlapping with either H3K27ac enriched regions or open chromatin regions in coronary artery or any of 5 single-cell clusters represented in artery tissue (SMCs, endothelial cells, fibroblasts, macrophages and T-lymphocytes). Lead SNP from each locus is indicated in bold and comes first before other SNPs in the locus. Other SNPs are ranked by genomic coordinates (hg38 genome). Manually curated best candidates SNPs are indicated by a *. Overlap with H3K27ac signal or open chromatin region is indicated by a +.

Chr: chromosome, r2: r2 of linkage desequilibrium with lead SNP of the locus (European population in 1000 Genome reference panel), PP: posterior probability ,EA: Effect allele, EAF: effect allele frequency, OR: Odds ratio, CI: confidence interval.

| Locus | Candidate SNP | SNP | Chr | Position (hg38) | Alleles | r2 | Correlated Alleles | PP | EA | EAF | Direction | P | OR (95% CI) | Coronary artery H3K27ac | Coronary artery ATAC-Seq | snATAC-Seq vascular SMCs | snATAC-Seq ECs | snATAC-Seq Fibroblasts | snATAC-Seq Macrophages | snATAC-Seq T-lymphocytes |
|---|---|---|---|---|---|---|---|---|---|---|---|---|---|---|---|---|---|---|---|---|
| *FGGY-DT* |  | **rs34370185** | **chr1** | **59656909** | **(G/T)** | **-** | **-** | **10.9%** | **G** | **0.71** | **--------** | **1.42E-12** | **0.74 (0.69-0.81)** |  |  |  |  |  |  |  |
| | | rs3737157 | chr1 | 59617813 | (A/G) | 0.708 | G=A,T=G | 3.8% | G | 0.30 | ++++++++ | 4.40E-12 | 1.33 (1.23-1.45) | + | | | | | | |
| | | rs12730605 | chr1 | 59627712 | (T/C) | 0.708 | G=T,T=C | 5.2% | C | 0.30 | ++++++++ | 2.94E-12 | 1.33 (1.23-1.45) | + | | | | | | |
|  |  | rs11207414 | chr1 | 59635514 | (G/T) | 0.708 | G=G,T=T | 3.9% | G | 0.70 | -------- | 4.08E-12 | 0.75 (0.69-0.81) |  | + |  |  |  |  |  |
| | | rs12745935 | chr1 | 59644082 | (A/G) | 0.708 | G=A,T=G | na | na | na | na | na | na | + | + | | | + | | |
| | * | rs12746365 | chr1 | 59644084 | (G/A) | 0.708 | G=G,T=A | na | na | na | na | na | na | + | + | | | + | | |
|  | * | rs17535443 | chr1 | 59646056 | (G/A) | 0.7744 | G=G,T=A | 0.5% | G | 0.72 | -------- | 3.40E-11 | 0.76 (0.7-0.82) | + | + |  |  |  |  |  |
|  | * | rs11207420 | chr1 | 59646524 | (G/A) | 0.7697 | G=G,T=A | 0.4% | G | 0.72 | -------- | 5.02E-11 | 0.76 (0.7-0.82) | + | + |  |  |  |  |  |
|  | * | rs12733512 | chr1 | 59646978 | (C/T) | 0.7744 | G=C,T=T | 0.1% | C | 0.72 | -------- | 1.53E-10 | 0.76 (0.7-0.83) | + | + |  |  |  |  |  |
|  |  | rs12130314 | chr1 | 59651063 | (G/T) | 0.7605 | G=G,T=T | 0.1% | G | 0.72 | -------- | 2.81E-10 | 0.77 (0.71-0.83) | + |  |  |  |  |  |  |
| | | rs12753566 | chr1 | 59654019 | (T/C) | 1 | G=T,T=C | 0.0% | C | 0.29 | ++?+++++ | 2.40E-08 | 1.31 (1.19-1.44) | + | | | | | | |
|  |  | rs12752853 | chr1 | 59668705 | (C/T) | 0.9567 | G=C,T=T | 0.0% | C | 0.71 | --?----- | 4.52E-07 | 0.78 (0.71-0.86) | + |  |  |  |  |  |  |
| | | rs12739904 | chr1 | 59669488 | (T/C) | 0.9567 | G=T,T=C | 1.2% | C | 0.29 | ++++++++ | 1.46E-11 | 1.32 (1.22-1.44) | + | + | | | | | |
|  | * | rs12758643 | chr1 | 59669918 | (C/T) | 0.9567 | G=C,T=T | 0.0% | C | 0.71 | --?----- | 4.05E-07 | 0.78 (0.71-0.86) | + | + | + | + | + | + |  |
| | * | rs7543389 | chr1 | 59680536 | (A/G) | 0.9567 | G=A,T=G | 0.8% | G | 0.29 | ++++++++ | 2.16E-11 | 1.32 (1.22-1.43) | + | + | + | | | | |
|  |  | rs10493256 | chr1 | 59688948 | (C/T) | 0.8222 | G=C,T=T | 0.0% | C | 0.70 | --?----- | 1.07E-06 | 0.79 (0.72-0.87) | + | + |  |  | + |  |  |
| *F3* | | **rs1146473** | **chr1** | **95050472** | **(T/C)** | **-** | **-** | **6.5%** | **C** | **0.19** | **++++++++** | **5.82E-09** | **1.32 (1.2-1.45)** | | | | | | | |
|  |  | rs1146481 | chr1 | 95045247 | (G/C) | 0.9593 | T=G,C=C | 1.4% | G | 0.82 | -------- | 3.09E-08 | 0.77 (0.7-0.84) | + |  |  |  |  |  |  |
|  |  | rs1146480 | chr1 | 95045362 | (G/A) | 0.9593 | T=G,C=A | 1.3% | G | 0.82 | -------- | 3.20E-08 | 0.77 (0.7-0.84) | + |  |  |  |  |  |  |
| | * | rs12354005 | chr1 | 95045461 | (A/G) | 0.9459 | T=A,C=G | 0.7% | G | 0.18 | ++++++++ | 6.04E-08 | 1.3 (1.18-1.43) | + | + | + | | + | | |
| | | rs34379452 | chr1 | 95046652 | (-/TT) | 0.9593 | T=-,C=TT | na | na | na | na | na | na | | + | | | | | |
|  |  | rs1772904 | chr1 | 95055132 | (C/T) | 0.9932 | T=C,C=T | 2.6% | C | 0.81 | -------- | 1.63E-08 | 0.77 (0.7-0.84) | + |  |  |  |  |  |  |
|  |  | rs968662 | chr1 | 95055317 | (T/A) | 0.9864 | T=T,C=A | 2.4% | T | 0.81 | -------- | 1.78E-08 | 0.76 (0.7-0.84) | + |  |  |  |  |  |  |
| | | rs968661 | chr1 | 95055331 | (A/G) | 0.9932 | T=A,C=G | 2.2% | G | 0.19 | ++++++++ | 1.84E-08 | 1.31 (1.19-1.43) | + | | | | | | |
| | * | rs1778208 | chr1 | 95056122 | (A/G) | 1 | T=A,C=G | 5.4% | G | 0.19 | ++++++++ | 7.19E-09 | 1.32 (1.2-1.44) | + | + | + | | + | | |
| | * | rs927636 | chr1 | 95056425 | (A/G) | 1 | T=A,C=G | 4.4% | G | 0.19 | ++++++++ | 8.67E-09 | 1.31 (1.2-1.44) | + | + | | | + | | |
| | * | rs927635 | chr1 | 95056533 | (A/G) | 0.9932 | T=A,C=G | 1.2% | G | 0.19 | ++++++++ | 3.44E-08 | 1.3 (1.18-1.43) | + | + | | | | | |
| | * | rs927634 | chr1 | 95056635 | (A/T) | 0.9865 | T=A,C=T | 4.5% | T | 0.19 | ++++++++ | 8.35E-09 | 1.31 (1.2-1.44) | + | + | | | | | |
| *FALEC/ADAMTSL4* | | **rs4970935** | **chr1** | **150504062** | **(C/T)** | **-** | **-** | **90.8%** | **C** | **0.28** | **++++++++** | **6.14E-39** | **1.72 (1.59-1.87)** | | | | | | | |
| | | rs4970996 | chr1 | 150506589 | (G/C) | 0.8742 | C=G,T=C | 8.9% | G | 0.27 | ++++++++ | 6.42E-38 | 1.72 (1.58-1.87) | + | | | | | | |
| | * | rs6693567 | chr1 | 150510660 | (C/T) | 0.9041 | C=C,T=T | 0.0% | C | 0.29 | ++?+++++ | 9.79E-29 | 1.7 (1.55-1.87) | + | + | + | | + | | |
| | | rs7364686 | chr1 | 150513711 | (A/G) | 0.7289 | C=A,T=G | na | na | na | na | na | na | | + | | | | | |
| *AFAP1* | | **rs6828005** | **chr4** | **7774352** | **(G/A)** | **-** | **-** | **31.6%** | **G** | **0.45** | **++++++++** | **2.60E-11** | **1.29 (1.2-1.4)** | | | | | | | |
|  |  | rs2285764 | chr4 | 7763161 | (T/C) | 0.3094 | G=T,A=C | 4.5% | C | 0.77 | -------- | 1.91E-10 | 0.75 (0.69-0.82) |  | + |  |  |  |  |  |
|  |  | rs2285762 | chr4 | 7763405 | (T/C) | 0.2975 | G=T,A=C | 9.0% | C | 0.77 | -------- | 9.31E-11 | 0.75 (0.69-0.82) |  | + | + |  | + |  |  |
|  | * | rs2269851 | chr4 | 7763605 | (T/C) | 0.2975 | G=T,A=C | 9.6% | C | 0.77 | -------- | 8.82E-11 | 0.75 (0.69-0.82) |  | + | + |  | + | + |  |
| | | **rs1507928** | **chr4** | **146788035** | **(T/C)** | **-** | **-** | **13.1%** | **C** | **0.48** | **++++++++** | **8.94E-09** | **1.25 (1.16-1.35)** | | | | | | | |
| | * | rs1979974 | chr4 | 146800815 | (A/G) | 0.9683 | T=A,C=G | 0.0% | G | 0.48 | ++?+++++ | 2.44E-05 | 1.21 (1.11-1.32) | + | + | + | | + | | |
|  | * | rs17020769 | chr4 | 146800922 | (C/T) | 0.9683 | T=C,C=T | 0.0% | C | 0.52 | --?----- | 6.04E-06 | 0.81 (0.75-0.89) | + | + | + |  | + |  |  |

| | | | | | | | | | | | | | | | | | | | | |
|---|---|---|---|---|---|---|---|---|---|---|---|---|---|---|---|---|---|---|---|---|
| ***ZNF827*** | * | rs13128814 | chr4 | 146801002 | (G/A) | 0.7085 | T=G,C=A | 4.1% | G | 0.49 | -------- | 3.15E-08 | 0.81 (0.75-0.87) | + | + | + |  | + |  |  |
| | | rs28590383 | chr4 | 146803248 | (T/C) | 0.9761 | T=T,C=C | 9.0% | C | 0.48 | ++++++++ | 1.34E-08 | 1.25 (1.16-1.35) | + | | | | | | |
|  | * | rs7679068 | chr4 | 146808682 | (C/T) | 0.7482 | T=C,C=T | 0.0% | C | 0.49 | --?----- | 5.59E-05 | 0.83 (0.76-0.91) | + | + | + | + | + |  |  |
| | * | rs7662069 | chr4 | 146809016 | (T/G) | 0.8921 | T=T,C=G | 12.3% | G | 0.49 | ++++++++ | 9.53E-09 | 1.25 (1.16-1.35) | + | + | | | + | | |
| | * | rs7662070 | chr4 | 146809017 | (T/C) | 0.8958 | T=T,C=C | 10.7% | C | 0.49 | ++++++++ | 1.11E-08 | 1.25 (1.16-1.35) | + | + | | | + | | |
| | | rs10000888 | chr4 | 146809168 | (A/G) | 0.7521 | T=A,C=G | 3.1% | G | 0.51 | ++++++++ | 4.27E-08 | 1.24 (1.15-1.34) | + | + | | | + | | |
| ***ITGA1*** | * | **rs73102285** | **chr5** | **52155642** | **(A/G)** | **-** | **-** | **7.3%** | **G** | **0.27** | **++++++--** | **1.05E-08** | **1.27 (1.17-1.38)** | | + | + | | | | |
| | | rs6867399 | chr5 | 52135543 | (C/A) | 0.701 | A=C,G=A | 0.0% | C | 0.71 | --?---+- | 9.82E-06 | 0.81 (0.73-0.89) | + | | | | | | |
| | | rs73756268 | chr5 | 52141109 | (T/G) | 0.9684 | A=T,G=G | 5.1% | G | 0.27 | ++++++-+ | 1.61E-08 | 1.27 (1.17-1.38) | + | | | | | | |
|  |  | rs73756269 | chr5 | 52141120 | (C/T) | 0.9684 | A=C,G=T | 4.9% | C | 0.73 | ------+- | 1.68E-08 | 0.79 (0.73-0.86) | + |  |  |  |  |  |  |
|  |  | rs12110170 | chr5 | 52142544 | (T/A) | 0.9737 | A=T,G=A | 6.6% | T | 0.73 | ------+- | 1.18E-08 | 0.79 (0.72-0.85) | + |  |  |  |  |  | + |
| | | rs60422098 | chr5 | 52143928 | (A/G) | 0.9737 | A=A,G=G | 4.0% | G | 0.27 | ++++++-+ | 2.12E-08 | 1.27 (1.17-1.37) | + | | | | | | |
| | | rs59192299 | chr5 | 52144090 | (T/G) | 0.9789 | A=T,G=G | 5.3% | G | 0.27 | ++++++-+ | 1.53E-08 | 1.27 (1.17-1.38) | + | | | | | | |
| | | rs35302472 | chr5 | 52145479 | (-/A) | 0.8617 | A=-,G=A | na | na | na | na | na | na | | | | + | | | |
| | | rs10513000 | chr5 | 52149451 | (A/G) | 0.9895 | A=A,G=G | 2.0% | G | 0.27 | ++++++-- | 4.56E-08 | 1.26 (1.16-1.37) | + | | | | | | |
| | | rs10513001 | chr5 | 52149529 | (T/G) | 0.9895 | A=T,G=G | 0.1% | G | 0.27 | ++?+++-- | 9.06E-07 | 1.27 (1.15-1.4) | + | | | | | | |
| | | rs10046016 | chr5 | 52150178 | (A/T) | 0.9842 | A=A,G=T | 2.1% | T | 0.27 | ++++++-- | 4.24E-08 | 1.26 (1.16-1.37) | + | | | | | | |
| | | rs7704305 | chr5 | 52150852 | (A/G) | 0.9737 | A=A,G=G | 2.3% | G | 0.27 | ++++++-- | 3.92E-08 | 1.26 (1.16-1.37) | + | | | + | | | |
|  |  | rs73754055 | chr5 | 52154837 | (G/C) | 0.9894 | A=G,G=C | 3.2% | G | 0.73 | ------++ | 2.59E-08 | 0.79 (0.73-0.86) | + |  |  |  |  |  |  |
| | | rs6886404 | chr5 | 52154995 | (A/C) | 0.9894 | A=A,G=C | 2.7% | C | 0.27 | ++++++-- | 3.05E-08 | 1.26 (1.16-1.37) | + | | | | | | |
|  |  | rs73754057 | chr5 | 52156781 | (T/A) | 0.9528 | A=T,G=A | 0.9% | T | 0.73 | ------++ | 9.93E-08 | 0.8 (0.73-0.87) | + |  |  |  |  |  |  |
|  |  | rs1478445 | chr5 | 52163105 | (G/A) | 0.9166 | A=G,G=A | 1.1% | G | 0.73 | ------++ | 8.12E-08 | 0.8 (0.73-0.86) | + |  |  |  |  |  |  |
|  |  | rs6884370 | chr5 | 52163352 | (G/C) | 0.9115 | A=G,G=C | 0.5% | G | 0.73 | ------++ | 2.05E-07 | 0.8 (0.74-0.87) | + |  |  |  |  |  |  |
| ***PHACTR1*** | * | **rs9349379** | **chr6** | **12903957** | **(A/G)** | **-** | **-** | **100.0%** | **G** | **0.38** | **--------** | **2.88E-32** | **0.61 (0.56-0.66)** | + | + |  |  |  |  |  |
| ***HTRA1*** | * | **rs2736923** | **chr10** | **124259062** | **(G/A)** | **-** | **-** | **4.9%** | **G** | **0.11** | **------?-** | **4.58E-08** | **0.69 (0.61-0.79)** | + | + | + |  | + | + |  |
|  |  | rs11200638 | chr10 | 124220544 | (G/A) | 0.0212 | NA | 1.2% | G | 0.78 | ----+-+- | 2.54E-07 | 0.79 (0.72-0.86) | + | + | + |  |  | + |  |
| | | rs2239588 | chr10 | 124250093 | (G/C) | 0.8201 | G=C,A=G | 0.5% | G | 0.91 | ++++?+++ | 5.31E-07 | 1.47 (1.26-1.7) | + | | | | | | |
| | | rs2268349 | chr10 | 124250384 | (G/T) | 0.9725 | G=T,A=G | 3.4% | G | 0.89 | ++++++++ | 6.54E-08 | 1.44 (1.26-1.64) | + | | | | | | |
| | * | rs2268350 | chr10 | 124251060 | (C/T) | 0.9725 | G=T,A=C | 4.5% | C | 0.89 | ++++++++ | 4.89E-08 | 1.44 (1.26-1.64) | + | + | | | | | + |
| | | rs2268351 | chr10 | 124251098 | (C/T) | 0.8201 | G=T,A=C | 0.0% | C | 0.91 | ??++?+?? | 5.94E-05 | 1.49 (1.22-1.8) | + | + | | | | | |
|  |  | rs2300434 | chr10 | 124251697 | (T/C) | 0.9725 | G=C,A=T | 3.6% | C | 0.11 | -------- | 6.23E-08 | 0.7 (0.61-0.79) | + |  |  |  |  |  |  |
| | | rs2300435 | chr10 | 124252279 | (G/A) | 0.8201 | G=A,A=G | 1.4% | G | 0.91 | ++++++++ | 1.64E-07 | 1.48 (1.28-1.71) | + | | | | | | |
| | | rs2300436 | chr10 | 124252285 | (C/T) | 0.8201 | G=T,A=C | 1.4% | C | 0.91 | ++++++++ | 1.62E-07 | 1.48 (1.28-1.71) | + | | | | | | |
| | | rs2268353 | chr10 | 124254061 | (C/T) | 0.9816 | G=T,A=C | 0.0% | C | 0.89 | ++?+++++ | 1.24E-05 | 1.4 (1.2-1.63) | + | | | | | | |
|  |  | rs2246731 | chr10 | 124254180 | (G/A) | 0.9908 | G=G,A=A | 1.7% | G | 0.11 | ------?- | 1.49E-07 | 0.7 (0.62-0.8) | + |  |  |  |  |  |  |
| | | rs11200654 | chr10 | 124254343 | (C/T) | 0.9816 | G=T,A=C | 4.2% | C | 0.89 | ++++++++ | 5.11E-08 | 1.44 (1.26-1.64) | + | | | | | | |
|  |  | rs3106650 | chr10 | 124254360 | (G/T) | 0.9908 | G=G,A=T | 1.0% | G | 0.11 | ------?- | 2.57E-07 | 0.71 (0.62-0.81) | + |  |  |  |  | + |  |
|  |  | rs10887154 | chr10 | 124255340 | (T/C) | 0.8201 | G=C,A=T | 1.4% | C | 0.09 | -------- | 1.61E-07 | 0.68 (0.59-0.78) | + | + |  |  |  |  |  |
| | | rs10887155 | chr10 | 124255686 | (G/T) | 0.8201 | G=T,A=G | 1.2% | G | 0.91 | ++++++++ | 1.82E-07 | 1.47 (1.27-1.7) | + | | | | | | |
|  | * | rs736960 | chr10 | 124258136 | (C/T) | 0.7226 | G=C,A=T | 0.9% | C | 0.10 | ------?- | 2.84E-07 | 0.69 (0.6-0.8) | + | + | + |  | + |  |  |
|  | * | rs2736922 | chr10 | 124258377 | (C/T) | 0.9908 | G=C,A=T | 1.0% | C | 0.11 | ------?- | 2.51E-07 | 0.71 (0.62-0.81) | + | + |  |  |  |  |  |
| | | rs2672607 | chr10 | 124258758 | (A/T) | 0.9908 | G=A,A=T | 1.0% | T | 0.89 | ++++++?+ | 2.56E-07 | 1.41 (1.24-1.61) | + | + | | | | | |
|  |  | rs2247430 | chr10 | 124259750 | (C/T) | 0.9908 | G=C,A=T | 1.4% | C | 0.11 | ------?- | 1.79E-07 | 0.7 (0.62-0.8) | + |  |  |  |  |  |  |
|  |  | rs11200655 | chr10 | 124260564 | (C/G) | 0.8201 | G=G,A=C | 1.3% | G | 0.09 | -------- | 1.75E-07 | 0.68 (0.59-0.78) | + |  |  |  |  |  |  |
|  |  | rs2247541 | chr10 | 124260739 | (C/T) | 0.9908 | G=C,A=T | 2.1% | C | 0.12 | ------?- | 1.17E-07 | 0.7 (0.61-0.8) | + |  |  |  |  |  |  |
| | | rs2250511 | chr10 | 124262419 | (A/G) | 0.9908 | G=A,A=G | 1.4% | G | 0.89 | ++++++?+ | 1.79E-07 | 1.42 (1.25-1.62) | + | | | | | | |
| | | rs2142309 | chr10 | 124263692 | (T/C) | 0.9817 | G=T,A=C | 2.1% | C | 0.88 | ++++++?+ | 1.14E-07 | 1.43 (1.25-1.63) | + | | | | | | |
| | | rs1009326 | chr10 | 124263873 | (C/T) | 0.811 | G=T,A=C | 1.6% | C | 0.91 | ++++++++ | 1.38E-07 | 1.48 (1.28-1.72) | + | | | | | | |
| | | rs1009325 | chr10 | 124263877 | (A/G) | 0.9817 | G=A,A=G | 2.8% | G | 0.88 | ++++++?+ | 8.53E-08 | 1.43 (1.26-1.64) | + | | | | | | |

| | | | | | | | | | | | | | | | | | | | |
|---|---|---|---|---|---|---|---|---|---|---|---|---|---|---|---|---|---|---|---|
| | | rs909290 | chr10 | 124264032 | (A/G) | 0.9727 | G=A,A=G | 3.7% | G | 0.88 | ++++++?+ | 6.16E-08 | 1.44 (1.26-1.65) | + | + | + | | | |
|  |  | rs17103659 | chr10 | 124274603 | (T/C) | 0.7933 | G=C,A=T | 1.4% | C | 0.09 | -------- | 1.55E-07 | 0.67 (0.58-0.78) |  |  |  |  | + |  |
| | | rs142773245 | chr10 | 124274615 | (-/T) | 0.7933 | G=T,A=- | na | na | na | na | na | na | | | | | + | |
|  |  | rs714989 | chr10 | 124280725 | (A/G) | 0.5295 | G=G,A=A | 4.2% | G | 0.18 | ------+- | 6.15E-08 | 0.74 (0.67-0.83) |  |  | + |  |  |  |
| ***SESN3/FAM76B*** | * | **rs11021221** | **chr11** | **95308854** | **(T/A)** | **-** | **-** | **91.2%** | **T** | **0.83** | **--------** | **4.11E-15** | **0.68 (0.62-0.75)** | + | + |  |  |  |  |
| | | rs1255528 | chr11 | 95299962 | (G/A) | 0.7814 | T=A,A=G | 0.1% | G | 0.20 | ++++++++ | 4.89E-12 | 1.38 (1.26-1.52) | + | | | | | |
| ***LRP1*** | * | **rs11172113** | **chr12** | **57527283** | **(T/C)** | **-** | **-** | **99.9%** | **C** | **0.38** | **--------** | **9.03E-31** | **0.62 (0.57-0.67)** | + | + |  |  |  |  |
| | | rs4759275 | chr12 | 57525756 | (G/A) | 0.7586 | T=G,C=A | 0.0% | G | 0.60 | ++++++++ | 4.56E-21 | 1.47 (1.36-1.59) | + | | | | | |
| | | rs4759276 | chr12 | 57526646 | (G/A) | 0.8512 | T=G,C=A | 0.0% | G | 0.64 | ++++++++ | 6.16E-26 | 1.57 (1.44-1.7) | + | | | | | |
| ***ATP2B1*** | | **rs1689040** | **chr12** | **89978233** | **(C/T)** | **-** | **-** | **23.5%** | **C** | **0.59** | **+++++++-** | **7.04E-10** | **1.28 (1.18-1.39)** | | | | | | |
| ***COL4A1*** | | **rs7326444** | **chr13** | **110838236** | **(G/A)** | **-** | **-** | **11.7%** | **G** | **0.64** | **++++++++** | **1.02E-10** | **1.31 (1.21-1.42)** | | | | | | |
| | | rs3783111 | chr13 | 110835049 | (C/T) | 0.9784 | G=C,A=T | 6.3% | C | 0.65 | ++++++++ | 2.12E-10 | 1.31 (1.2-1.42) | + | | | | | |
|  |  | rs2305080 | chr13 | 110838703 | (T/C) | 0.9957 | G=T,A=C | 6.5% | C | 0.36 | -------- | 2.00E-10 | 0.77 (0.71-0.83) | + |  |  |  |  |  |
|  |  | rs9521638 | chr13 | 110839428 | (A/G) | 0.987 | G=A,A=G | 0.0% | G | 0.36 | --?----- | 1.93E-07 | 0.78 (0.71-0.85) | + |  |  |  |  |  |
|  | * | rs2391817 | chr13 | 110840412 | (A/T) | 0.9699 | G=A,A=T | 4.5% | T | 0.36 | -------- | 3.01E-10 | 0.77 (0.71-0.83) | + | + |  |  |  |  |
| | | rs1977892 | chr13 | 110840624 | (G/T) | 0.9656 | G=G,A=T | 6.1% | G | 0.65 | ++++++++ | 2.16E-10 | 1.31 (1.2-1.42) | + | + | | | | |
|  |  | rs1977893 | chr13 | 110840860 | (T/C) | 0.9656 | G=T,A=C | 0.0% | C | 0.36 | --?----- | 4.22E-08 | 0.77 (0.7-0.84) | + | + |  |  |  |  |
| | | rs3783107 | chr13 | 110842102 | (G/A) | 0.8918 | G=G,A=A | 0.2% | G | 0.63 | ++++++++ | 9.42E-09 | 1.27 (1.17-1.38) | + | | | | | |
| | | rs3783106 | chr13 | 110842153 | (T/G) | 0.8841 | G=G,A=T | 0.0% | G | 0.63 | ++?+++++ | 1.02E-05 | 1.24 (1.12-1.36) | + | | | | | |
|  |  | rs3783105 | chr13 | 110842158 | (T/A) | 0.8841 | G=A,A=T | 0.0% | T | 0.37 | -------- | 1.22E-07 | 0.8 (0.74-0.87) | + |  |  |  |  |  |
| ***COL4A2*** | * | **rs11838776** | **chr13** | **111040681** | **(G/A)** | **-** | **-** | **78.2%** | **G** | **0.73** | **+++++++-** | **2.46E-18** | **1.5 (1.37-1.65)** | + | + | + | | | |
| | | rs9583489 | chr13 | 111032833 | (C/T) | 0.8178 | G=C,A=T | 0.0% | C | 0.73 | ++?++++- | 7.43E-12 | 1.44 (1.3-1.6) | + | | | | | |
| | | rs2391825 | chr13 | 111035483 | (G/A) | 0.8262 | G=G,A=A | 0.0% | G | 0.73 | +++++++- | 6.45E-14 | 1.41 (1.29-1.55) | + | | | | | |
| | | rs80281875 | chr13 | 111038374 | (T/C) | 0.8307 | G=T,A=C | NA | NA | NA | NA | NA | #VALEUR! | + | | | | | |
|  |  | rs75345389 | chr13 | 111038379 | (T/C) | 0.8265 | G=T,A=C | 0.0% | C | 0.26 | ----?-?+ | 4.09E-13 | 0.7 (0.63-0.77) | + |  |  |  |  |  |
| | * | rs9515201 | chr13 | 111040798 | (A/C) | 0.8982 | G=C,A=A | 2.6% | C | 0.71 | ++++-++- | 9.31E-17 | 1.45 (1.33-1.59) | + | + | | | | |
| | | rs4773174 | chr13 | 111041530 | (C/T) | 0.7804 | G=C,A=T | 0.0% | C | 0.77 | +++++++- | 1.45E-12 | 1.42 (1.29-1.56) | + | | | | | |
|  |  | rs55940034 | chr13 | 111043309 | (A/G) | 0.9406 | G=A,A=G | 19.2% | G | 0.28 | -------+ | 1.17E-17 | 0.68 (0.62-0.74) | + |  |  |  |  |  |
| ***FBN1*** | | **rs7174973** | **chr15** | **48763754** | **(A/G)** | **-** | **-** | **99.8%** | **G** | **0.11** | **++++++++** | **1.60E-13** | **1.54 (1.37-1.72)** | | | | | | |
| | | rs61999107 | chr15 | 48691494 | (C/T) | 0.9112 | A=C,G=T | 0.0% | C | 0.89 | --?---?- | 0.0008402 | 0.79 (0.69-0.91) | + | | | | | |
| | | rs2015637 | chr15 | 48716853 | (T/C) | 0.9594 | A=T,G=C | 0.0% | C | 0.11 | ++++++?+ | 3.07E-09 | 1.42 (1.27-1.6) | + | | | | | |
|  |  | rs75225368 | chr15 | 48785206 | (C/T) | 0.9382 | A=C,G=T | 0.0% | C | 0.90 | ?------- | 2.44E-07 | 0.71 (0.62-0.81) | + |  |  |  |  |  |
| | | rs8026752 | chr15 | 48823770 | (A/G) | 0.8999 | A=A,G=G | 0.0% | G | 0.10 | ?+++++++ | 3.39E-07 | 1.4 (1.23-1.6) | + | | + | | | |
|  |  | rs595244 | chr15 | 48840835 | (C/T) | 0.8887 | A=C,G=T | 0.0% | C | 0.89 | -------- | 1.57E-09 | 0.7 (0.62-0.79) | + |  |  |  |  |  |
|  | * | rs1561207 | chr15 | 48858971 | (G/T) | 0.899 | A=G,G=T | 0.0% | G | 0.89 | -------- | 2.41E-09 | 0.7 (0.62-0.79) | + | + |  |  |  |  |
|  | * | rs2437947 | chr15 | 48876881 | (T/A) | 0.899 | A=T,G=A | 0.0% | T | 0.89 | -------- | 2.94E-09 | 0.7 (0.63-0.79) |  | + | + |  |  |  |
| | | rs686861 | chr15 | 48885005 | (A/G) | 0.8896 | A=A,G=G | 0.0% | G | 0.11 | ++++++++ | 4.66E-09 | 1.41 (1.26-1.59) | | + | | | | |
|  |  | rs2456475 | chr15 | 48885877 | (G/A) | 0.8896 | A=G,G=A | 0.0% | G | 0.89 | -------- | 6.30E-09 | 0.71 (0.63-0.8) | + |  |  |  |  |  |
| | | rs2455925 | chr15 | 48893649 | (T/C) | 0.8896 | A=T,G=C | 0.0% | C | 0.11 | ++++++++ | 6.84E-09 | 1.41 (1.25-1.58) | + | | | | | |
| | | rs591519 | chr15 | 48894200 | (A/T) | 0.8896 | A=A,G=T | 0.0% | T | 0.11 | ++++++++ | 6.06E-09 | 1.41 (1.26-1.58) | + | | | | | |
| | | rs1036476 | chr15 | 48914775 | (T/C) | 0.8511 | A=T,G=C | 0.0% | C | 0.11 | ++++++++ | 8.61E-09 | 1.41 (1.25-1.58) | | | | | + | |
|  |  | rs689304 | chr15 | 48922360 | (C/T) | 0.8692 | A=C,G=T | 0.0% | C | 0.89 | -------- | 9.67E-09 | 0.71 (0.63-0.8) | + |  |  |  |  |  |
| | * | rs4775769 | chr15 | 48939888 | (T/G) | 0.8196 | A=G,G=T | 0.0% | G | 0.89 | ---?-??- | 6.12E-07 | 0.7 (0.61-0.8) | + | | | | | |
| ***THSD4*** |  | **rs10851839** | **chr15** | **71628370** | **(T/A)** | **-** | **-** | **30.4%** | **T** | **0.32** | **------+-** | **5.51E-11** | **0.75 (0.69-0.82)** |  |  |  |  |  |  |
|  |  | rs3743111 | chr15 | 71587373 | (G/A) | 0.7033 | T=G,A=A | 0.0% | G | 0.39 | --?----- | 2.06E-06 | 0.8 (0.73-0.88) |  |  |  | + |  |  |
| | * | rs11853359 | chr15 | 71621524 | (G/A) | 0.957 | T=A,A=G | 0.1% | G | 0.67 | ++?+++-+ | 2.96E-08 | 1.31 (1.19-1.44) | + | + | + | + | + | |
| ***LINC00310/KCNE2*** | * | **rs28451064** | **chr21** | **35593827** | **(G/A)** | **-** | **-** | **99.9%** | **G** | **0.88** | **++++++++** | **1.16E-22** | **2.04 (1.77-2.35)** | + | + | | | | |
|  |  | rs9305545 | chr21 | 35595821 | (A/G) | 0.8197 | G=A,A=G | 0.0% | G | 0.15 | --?----- | 1.16E-15 | 0.56 (0.49-0.65) |  |  |  |  |  | + |
| | | rs9982601 | chr21 | 35599128 | (C/T) | 0.7805 | G=C,A=T | 0.0% | C | 0.87 | +++++++- | 9.58E-17 | 1.75 (1.53-1.99) | + | | | | | |

| | | | | | | | | | | | | | | | | | | |
|---|---|---|---|---|---|---|---|---|---|---|---|---|---|---|---|---|---|---|
| | | rs9980618 | chr21 | 35600505 | (C/T) | 0.7805 | G=C,A=T | 0.0% | C | 0.87 | +++++++- | 7.18E-17 | 1.76 (1.54-2) | + | | | | |
| *SYN3* |  | **rs137507** | **chr22** | **33282971** | **(T/C)** | **-** | **-** | **17.7%** | **C** | **0.89** | **-----+--** | **3.30E-08** | **0.73 (0.65-0.81)** |  |  |  |  |  |
|  |  | rs137498 | chr22 | 33276313 | (C/G) | 0.189 | T=C,C=G | 2.5% | G | 0.56 | -------- | 3.32E-07 | 0.82 (0.76-0.88) |  |  |  |  | + |
|  |  | rs137500 | chr22 | 33276686 | (A/G) | 0.189 | T=A,C=G | 2.4% | G | 0.56 | -------- | 3.57E-07 | 0.82 (0.76-0.88) |  | + |  |  |  |
| | * | rs4448 | chr22 | 33276999 | (G/A) | 0.189 | T=G,C=A | 2.5% | G | 0.44 | ++++++++ | 3.27E-07 | 1.22 (1.13-1.32) | | + | + | + | + |
|  | * | rs137503 | chr22 | 33277200 | (C/G) | 0.189 | T=C,C=G | 2.0% | G | 0.56 | -------- | 4.21E-07 | 0.82 (0.76-0.89) |  | + | + | + | + |
|  |  | rs4452 | chr22 | 33283257 | (T/C) | 1 | T=T,C=C | 4.5% | C | 0.89 | -----+-- | 1.57E-07 | 0.74 (0.66-0.83) |  | + |  |  |  |
|  |  | rs137509 | chr22 | 33284221 | (T/G) | 0.8972 | T=T,C=G | 0.2% | G | 0.90 | -----+-- | 5.02E-06 | 0.76 (0.67-0.85) |  | + |  |  |  |
| | | rs80693 | chr22 | 33300347 | (G/T) | 0.9811 | T=G,C=T | 4.3% | G | 0.12 | +++++-++ | 1.62E-07 | 1.36 (1.21-1.52) | | | + | | |
| *FAM47A/TMEM47* | | **rs5973204** | **chrX** | **34489890** | **(C/T)** | **-** | **-** | **18.1%** | **C** | **0.80** | **--?--?--** | **2.97E-08** | **0.74 (0.66-0.82)** | | | | | |
| | | rs12395754 | chrX | 34455709 | (G/A) | 0.7296 | C=G,T=A | 0.0% | G | 0.81 | -??--?-- | 0.00352 | 0.82 (0.72-0.94) | | | | | + |
| | | rs12395785 | chrX | 34455809 | (G/A) | 0.7296 | C=G,T=A | 0.0% | G | 0.81 | -??--?-- | 0.00323 | 0.82 (0.72-0.94) | | | | | + |

**Supplementary Table 5:** Colocalization of association with SCAD and eQTL association.
We display the results of approxiate Bayes factor colocalization analysis for the association with SCAD, on one hand, eQTL association with the indicated eGenes, on the other hand.
NSNPs: number of SNPs used for he analysis. All SNPs present in both studies in a 2Mb window centered on SCAD lead variant were used. H0-H4: Posterior probability that: H0: neither trait has a genetic association in the region; H1: only trait 1 has a genetic association in the region; H2: only trait 2 has a genetic association in the region; H3: both traits are associated, but with different causal variants; H4: both traits are associated and share a single causal variant.

| SCAD lead SNP | eGene | Tissue | SCAD lead SNP eQTL P-value | Best eQTL SNP | Best eQTL P-value | NSNPs | H0 | H1 | H2 | H3 | H4 |
|---|---|---|---|---|---|---|---|---|---|---|---|
| rs34370185 | *FGGY-DT* | Artery_Coronary | 9.6E-16 | rs7543389 | 1.3E-19 | 1780 | 0% | 0% | 0% | 9% | 91% |
| | *FGGY-DT* | Artery_Tibial | 1.4E-61 | rs7543389 | 1.5E-69 | 1780 | 0% | 0% | 0% | 9% | 91% |
| | *FGGY-DT* | Artery_Aorta | 6.6E-53 | rs7543389 | 6.0E-59 | 1780 | 0% | 0% | 0% | 9% | 91% |
| rs1146473 | *F3* | Artery_Tibial | 3.8E-12 | rs60897247 | 2.8E-16 | 2317 | 0% | 0% | 0% | 100% | 0% |
| | *F3* | Artery_Aorta | 1.3E-04 | rs1612481 | 3.6E-08 | 2317 | 0% | 0% | 0% | 6% | 94% |
| rs4970935 | *ECM1* | Cells_Cultured_fibroblasts | 3.8E-20 | rs11801255 | 5.2E-25 | 1555 | 0% | 0% | 0% | 100% | 0% |
| | *ECM1* | Artery_Aorta | 1.7E-07 | rs6693567 | 5.4E-11 | 1555 | 0% | 0% | 0% | 0% | 100% |
| | *ADAMTSL4* | Cells_Cultured_fibroblasts | 2.0E-28 | rs6693567 | 1.5E-34 | 1555 | 0% | 0% | 0% | 100% | 0% |
| | *ADAMTSL4* | Artery_Tibial | 8.9E-24 | rs6693567 | 5.9E-29 | 1555 | 0% | 0% | 0% | 51% | 49% |
| | *ADAMTSL4* | Artery_Aorta | 4.5E-18 | rs6693567 | 1.7E-22 | 1555 | 0% | 0% | 0% | 72% | 28% |
| | *HORMAD1* | Cells_Cultured_fibroblasts | 3.2E-73 | rs7521898 | 1.2E-82 | 1555 | 0% | 0% | 0% | 100% | 0% |
| | *CTSS* | Artery_Tibial | 1.5E-24 | rs41271951 | 5.5E-30 | 1555 | 0% | 0% | 0% | 100% | 0% |
| | *ADAMTSL4-AS2* | Artery_Aorta | 1.1E-17 | rs11204664 | 2.5E-22 | 1555 | 0% | 0% | 0% | 100% | 0% |
| | *ADAMTSL4-AS2* | Artery_Coronary | 2.1E-10 | rs11204664 | 4.7E-14 | 1555 | 0% | 0% | 0% | 100% | 0% |
| | *ADAMTSL4-AS2* | Artery_Tibial | 6.0E-10 | rs11204664 | 5.9E-14 | 1555 | 0% | 0% | 0% | 100% | 0% |
| | *MRPS21* | Artery_Aorta | 3.2E-13 | rs6693697 | 3.2E-17 | 1555 | 0% | 0% | 0% | 100% | 0% |
| | *MRPS21* | Artery_Tibial | 8.4E-09 | rs3818978 | 1.2E-12 | 1555 | 0% | 0% | 0% | 97% | 3% |
| | *MRPS21* | Cells_Cultured_fibroblasts | 7.0E-10 | rs34629240 | 6.6E-14 | 1555 | 0% | 0% | 0% | 100% | 0% |
| rs6828005 | *AFAP1* | Whole_Blood | 1.2E-145 | rs56069442 | 6.9E-159 | 4147 | 0% | 0% | 0% | 100% | 0% |
| rs1507928 | *ZNF827* | Artery_Tibial | 7.2E-17 | rs13128814 | 1.3E-21 | 2004 | 0% | 0% | 0% | 3% | 97% |
| | *ZNF827* | Artery_Aorta | 3.5E-08 | rs13124853 | 6.9E-12 | 2004 | 0% | 0% | 0% | 6% | 94% |
| | *RP11-6L6.4* | Artery_Tibial | 3.7E-19 | rs1865531 | 1.7E-23 | 2004 | 0% | 0% | 0% | 100% | 0% |
| rs73102285 | *ITGA1* | Artery_Tibial | 2.6E-04 | rs10038615 | 8.5E-08 | 3001 | 0% | 0% | 0% | 4% | 96% |
| rs9349379 | *PHACTR1* | Artery_Tibial | 1.1E-35 | rs9349379 | 8.0E-42 | 2207 | 0% | 0% | 0% | 0% | 100% |
| | *PHACTR1* | Artery_Aorta | 5.3E-13 | rs9349379 | 2.0E-17 | 2207 | 0% | 0% | 0% | 0% | 100% |
| | *PHACTR1* | Artery_Coronary | 1.1E-05 | rs9349379 | 3.0E-09 | 2207 | 0% | 0% | 0% | 0% | 100% |
| | *TBC1D7* | Artery_Tibial | 7.8E-09 | rs499818 | 2.1E-11 | 2207 | 0% | 0% | 0% | 98% | 2% |
| | *GFOD1* | Artery_Tibial | 7.9E-12 | rs7742086 | 7.7E-15 | 2041 | 0% | 0% | 0% | 100% | 0% |
| | *RP1-257A7.4* | Artery_Tibial | 2.0E-10 | rs398151 | 1.9E-14 | 2207 | 0% | 0% | 0% | 100% | 0% |
| rs11021221 | *SESN3* | Artery_Tibial | 6.8E-11 | rs11021221 | 5.4E-15 | 2289 | 0% | 0% | 0% | 0% | 100% |
| | *SESN3* | Artery_Coronary | 8.0E-06 | rs10831371 | 2.5E-09 | 2288 | 0% | 0% | 0% | 88% | 12% |
| | *SESN3* | Artery_Aorta | 1.1E-07 | rs11021233 | 2.7E-11 | 2289 | 0% | 0% | 0% | 0% | 100% |
| rs11172113 | *LRP1* | Artery_Aorta | 2.7E-11 | rs11172113 | 3.6E-15 | 1371 | 0% | 0% | 0% | 0% | 100% |
| | *LRP1* | Artery_Tibial | 1.9E-16 | rs11172113 | 9.4E-21 | 1371 | 0% | 0% | 0% | 0% | 100% |
| rs1689040 | *ATP2B1* | Artery_Tibial | 1.7E-14 | rs2681472 | 1.6E-18 | 1682 | 0% | 0% | 0% | 12% | 88% |
| rs11838776 | *COL4A1* | Cells_Cultured_fibroblasts | 3.2E-19 | rs1927342 | 6.6E-24 | 1941 | 0% | 0% | 0% | 100% | 0% |
| rs7174973 | *FBN1* | Whole_Blood | 4.2E-34 | rs924816 | 6.2E-40 | 1243 | 0% | 0% | 0% | 100% | 0% |
| | *RP11-227D13.1* | Artery_Aorta | 8.7E-05 | rs55694948 | 3.1E-08 | 1243 | 0% | 1% | 0% | 14% | 85% |
| | *RP11-227D13.1* | Artery_Tibial | 9.3E-08 | rs6493333 | 2.4E-11 | 1243 | 0% | 0% | 0% | 100% | 0% |
| | *RP11-227D13.1* | Cells_Cultured_fibroblasts | 7.4E-32 | rs6493333 | 7.4E-38 | 1243 | 0% | 0% | 0% | 100% | 0% |
| rs10851839 | *THSD4* | Artery_Aorta | 2.6E-08 | rs11853359 | 4.0E-12 | 2075 | 0% | 0% | 0% | 2% | 98% |
| | *NR2E3* | Artery_Aorta | 2.8E-02 | rs34719283 | 1.6E-05 | 2075 | 0% | 5% | 0% | 4% | 92% |
| rs28451064 | *KCNE2* | Artery_Aorta | 1.9E-04 | rs9980618 | 5.9E-08 | 1992 | 0% | 0% | 0% | 1% | 99% |
| | *SLC5A3* | Artery_Tibial | 1.0E-03 | rs8128536 | 3.0E-07 | 1992 | 0% | 1% | 0% | 25% | 74% |
| | *SLC5A3* | Artery_Aorta | 1.1E-05 | rs28451064 | 3.2E-09 | 1992 | 0% | 0% | 0% | 0% | 100% |
| | *AP000318.2* | Artery_Aorta | 1.8E-04 | rs80284318 | 4.6E-08 | 1992 | 0% | 0% | 0% | 1% | 98% |
| | *LINC00310* | Artery_Aorta | 7.0E-06 | rs9305545 | 1.3E-09 | 1992 | 0% | 0% | 0% | 4% | 96% |
| | *MRPS6* | Artery_Aorta | 6.9E-07 | rs28451064 | 1.2E-10 | 1992 | 0% | 0% | 0% | 0% | 100% |
| | *MRPS6* | Artery_Tibial | 2.7E-03 | rs1018757 | 2.0E-06 | 1992 | 0% | 1% | 0% | 10% | 89% |

**Supplementary Table 6: Transcriptome-wide associated genes with SCAD in 3 artery tissues (GTEx v7 gene expression models).**
Genes with TWAS FDR < 0.05 are shown. Chr: chromosome, P.Bonf : Bonferroni corrected P-value. FDR: False Discovery Rate

| | | | | | Best GWAS SNP | | Best eQTL SNP | | TWAS | | | |
|---|---|---|---|---|---|---|---|---|---|---|---|---|
| **Gene** | **Tissue** | **Chr** | **Start** | **End** | **rsID** | **Z-Score** | **rsID** | **Z-Score** | **Z-score** | **P-value** | **P.Bonf** | **FDR** |
| *ADAMTSL4* | Cells_Transformed_fibroblasts | 1 | 150521884 | 150533413 | rs834238 | 11.8 | rs6693567 | -8.0 | -11.7 | 2.E-31 | 5.E-27 | 5.E-27 |
| *LRP1* | Artery_Tibial | 12 | 57522276 | 57607134 | rs11172113 | -11.5 | rs11172113 | -8.2 | 11.5 | 9.E-31 | 2.E-26 | 1.E-26 |
| *ADAMTSL4* | Artery_Tibial | 1 | 150521884 | 150533413 | rs834238 | 11.8 | rs6693567 | -7.6 | -11.4 | 3.E-30 | 8.E-26 | 3.E-26 |
| *ADAMTSL4* | Artery_Aorta | 1 | 150521884 | 150533413 | rs834238 | 11.8 | rs6693567 | -7.1 | -11.4 | 5.E-30 | 1.E-25 | 3.E-26 |
| *PHACTR1* | Artery_Aorta | 6 | 12717893 | 13288645 | rs9349379 | -11.8 | rs9349379 | -6.2 | 11.2 | 6.E-29 | 2.E-24 | 3.E-25 |
| *PHACTR1* | Artery_Tibial | 6 | 12717893 | 13288645 | rs9349379 | -11.8 | rs9349379 | -9.2 | 10.0 | 2.E-23 | 6.E-19 | 1.E-19 |
| *ECM1* | Artery_Aorta | 1 | 150480538 | 150486265 | rs834238 | 11.8 | rs6693567 | -3.9 | -9.2 | 5.E-20 | 1.E-15 | 2.E-16 |
| *ADAMTSL4-AS2* | Artery_Aorta | 1 | 150521040 | 150530200 | rs834238 | 11.8 | rs6693567 | -4.9 | -8.7 | 4.E-18 | 1.E-13 | 1.E-14 |
| *ADAMTSL4-AS2* | Artery_Coronary | 1 | 150521040 | 150530200 | rs834238 | 11.8 | rs6655975 | -3.9 | -7.4 | 1.E-13 | 3.E-09 | 4.E-10 |
| *ECM1* | Cells_Transformed_fibroblasts | 1 | 150480538 | 150486265 | rs834238 | 11.8 | rs10888585 | 6.8 | -7.1 | 2.E-12 | 5.E-08 | 5.E-09 |
| *C1orf54* | Artery_Coronary | 1 | 150240967 | 150253327 | rs834238 | 11.8 | rs1932934 | 3.6 | 7.0 | 3.E-12 | 8.E-08 | 7.E-09 |
| *SESN3* | Artery_Aorta | 11 | 94898704 | 94965705 | rs2186739 | 7.2 | rs11021233 | -5.4 | -6.4 | 1.E-10 | 3.E-06 | 3.E-07 |
| *APH1A* | Whole_Blood | 1 | 150237804 | 150241980 | rs834238 | 11.8 | rs2275780 | -3.6 | -6.2 | 5.E-10 | 1.E-05 | 1.E-06 |
| *FGGY-DT* | Artery_Aorta | 1 | 59597608 | 59664293 | rs12739770 | 7.1 | rs12758288 | 11.0 | 6.0 | 2.E-09 | 5.E-05 | 4.E-06 |
| *FGGY-DT* | Artery_Coronary | 1 | 59597608 | 59664293 | rs12739770 | 7.1 | rs7543389 | 6.5 | 6.0 | 3.E-09 | 7.E-05 | 5.E-06 |
| *SESN3* | Artery_Tibial | 11 | 94898704 | 94965705 | rs2186739 | 7.2 | rs11021233 | -7.4 | -5.8 | 8.E-09 | 2.E-04 | 1.E-05 |
| *ADAMTSL4-AS2* | Artery_Tibial | 1 | 150521040 | 150530200 | rs834238 | 11.8 | rs9659073 | 3.4 | -5.8 | 8.E-09 | 2.E-04 | 1.E-05 |
| *FGGY-DT* | Artery_Tibial | 1 | 59597608 | 59664293 | rs12739770 | 7.1 | rs12758643 | 12.6 | 5.4 | 9.E-08 | 2.E-03 | 1.E-04 |
| *C1orf54* | Artery_Aorta | 1 | 150240967 | 150253327 | rs834238 | 11.8 | rs751931 | -4.9 | 5.0 | 6.E-07 | 2.E-02 | 9.E-04 |
| *F3* | Artery_Aorta | 1 | 94994781 | 95007356 | rs1146474 | 5.7 | rs1493276 | -3.8 | -4.9 | 1.E-06 | 3.E-02 | 1.E-03 |
| *MRPS6* | Artery_Aorta | 21 | 35496253 | 35515334 | rs9305545 | -8.0 | rs12627001 | 3.9 | -4.9 | 1.E-06 | 3.E-02 | 2.E-03 |
| *MRPS6* | Artery_Tibial | 21 | 35496253 | 35515334 | rs9305545 | -8.0 | rs1018757 | 5.0 | -4.8 | 2.E-06 | 5.E-02 | 2.E-03 |
| *ZNF827* | Artery_Tibial | 4 | 146678779 | 146859787 | rs1507928 | 5.8 | rs1979974 | 7.5 | 4.7 | 3.E-06 | 8.E-02 | 3.E-03 |
| *LTBP3* | Whole_Blood | 11 | 65306276 | 65326401 | rs1783541 | 4.9 | rs17146964 | 5.8 | 4.6 | 4.E-06 | 1.E-01 | 5.E-03 |
| *PHACTR1* | Cells_Transformed_fibroblasts | 6 | 12717893 | 13288645 | rs9349379 | -11.8 | rs7757858 | 4.5 | 4.6 | 4.E-06 | 1.E-01 | 5.E-03 |
| *COL4A1* | Cells_Transformed_fibroblasts | 13 | 110801318 | 110959496 | rs9515201 | -8.3 | rs7991842 | -6.8 | 4.6 | 5.E-06 | 1.E-01 | 5.E-03 |
| *NT5DC1* | Artery_Aorta | 6 | 116422012 | 116570660 | rs12208531 | -4.4 | rs550480 | 7.0 | -4.5 | 8.E-06 | 2.E-01 | 7.E-03 |
| *ATP2B1* | Artery_Tibial | 12 | 89981828 | 90102608 | rs6538195 | -5.7 | rs2681492 | 7.1 | -4.5 | 8.E-06 | 2.E-01 | 7.E-03 |
| *CDC42SE1* | Cells_Transformed_fibroblasts | 1 | 151023897 | 151042801 | rs3820539 | 5.5 | rs4970998 | 4.7 | -4.5 | 8.E-06 | 2.E-01 | 7.E-03 |
| *RPRD2* | Whole_Blood | 1 | 150335567 | 150449042 | rs834238 | 11.8 | rs486275 | -4.1 | 4.4 | 1.E-05 | 3.E-01 | 9.E-03 |
| *ANKRD42* | Artery_Tibial | 11 | 82904837 | 82967141 | rs17519683 | -4.7 | rs565674 | 9.3 | -4.4 | 1.E-05 | 3.E-01 | 9.E-03 |
| *GGCX* | Whole_Blood | 2 | 85772404 | 85788670 | rs6705971 | -4.6 | rs10198569 | 10.2 | -4.4 | 1.E-05 | 4.E-01 | 1.E-02 |
| *ANKRD42* | Artery_Aorta | 11 | 82904837 | 82967141 | rs17519683 | -4.7 | rs647644 | 5.4 | -4.3 | 1.E-05 | 4.E-01 | 1.E-02 |
| *FRK* | Artery_Coronary | 6 | 116252312 | 116381921 | rs12208531 | -4.4 | rs195529 | 4.4 | -4.3 | 2.E-05 | 4.E-01 | 1.E-02 |
| *TARS2* | Artery_Tibial | 1 | 150459887 | 150480078 | rs834238 | 11.8 | rs13294 | -5.3 | 4.3 | 2.E-05 | 5.E-01 | 1.E-02 |

| | | | | | | | | | | | | |
|---|---|---|---|---|---|---|---|---|---|---|---|---|
| *CCDC90B* | Artery_Tibial | 11 | 82970139 | 82997170 | rs17519683 | -4.7 | rs6592108 | 14.5 | -4.3 | 2.E-05 | 5.E-01 | 1.E-02 |
| *AGBL2* | Artery_Aorta | 11 | 47681143 | 47736941 | rs1377416 | 4.5 | rs11828339 | 3.9 | -4.2 | 2.E-05 | 6.E-01 | 2.E-02 |
| *CCDC90B* | Artery_Aorta | 11 | 82970139 | 82997170 | rs17519683 | -4.7 | rs647644 | 11.4 | -4.2 | 2.E-05 | 6.E-01 | 2.E-02 |
| *TBC1D7* | Artery_Tibial | 6 | 13266774 | 13328815 | rs9349379 | -11.8 | rs2458307 | 6.3 | 4.2 | 3.E-05 | 8.E-01 | 2.E-02 |
| *MIA3* | Artery_Aorta | 1 | 222791428 | 222841354 | rs17163303 | 4.0 | rs2133189 | 3.9 | 4.2 | 3.E-05 | 8.E-01 | 2.E-02 |
| *SLC24A3* | Artery_Aorta | 20 | 19193290 | 19703581 | rs4813361 | 4.5 | rs6046169 | -5.8 | -4.2 | 3.E-05 | 9.E-01 | 2.E-02 |
| *GGCX* | Artery_Tibial | 2 | 85772404 | 85788670 | rs6705971 | -4.6 | rs10187424 | 9.3 | -4.1 | 3.E-05 | 9.E-01 | 2.E-02 |
| *CCDC90B* | Artery_Coronary | 11 | 82970139 | 82997170 | rs17519683 | -4.7 | rs7111383 | 7.5 | -4.1 | 4.E-05 | 1.E+00 | 2.E-02 |
| *GJA1* | Artery_Tibial | 6 | 121756838 | 121770873 | rs10499112 | 3.8 | rs9490305 | 5.4 | 4.1 | 4.E-05 | 1.E+00 | 2.E-02 |
| *RAPGEF6* | Artery_Aorta | 5 | 130759614 | 130970929 | rs924434 | 3.7 | rs6873582 | 4.3 | 4.1 | 5.E-05 | 1.E+00 | 3.E-02 |
| *MAT2A* | Artery_Tibial | 2 | 85766288 | 85771845 | rs6705971 | -4.6 | rs3821021 | 4.6 | -4.1 | 5.E-05 | 1.E+00 | 3.E-02 |
| *COL4A2* | Cells_Transformed_fibroblasts | 13 | 110958159 | 111165374 | rs9515201 | -8.3 | rs7991842 | -6.1 | 4.0 | 6.E-05 | 1.E+00 | 3.E-02 |
| *INO80E* | Whole_Blood | 16 | 30006615 | 30016829 | rs3809624 | 5.1 | rs4787491 | 5.5 | 4.0 | 6.E-05 | 1.E+00 | 3.E-02 |
| *SLC24A3* | Artery_Tibial | 20 | 19193290 | 19703581 | rs4813361 | 4.5 | rs3790227 | -5.9 | -4.0 | 7.E-05 | 1.E+00 | 4.E-02 |
| *GGCX* | Artery_Coronary | 2 | 85772404 | 85788670 | rs6705971 | -4.6 | rs2886722 | 4.0 | -4.0 | 7.E-05 | 1.E+00 | 4.E-02 |
| *LRCH1* | Artery_Aorta | 13 | 47127303 | 47327175 | rs17282423 | 4.1 | rs7982810 | 5.0 | 3.9 | 9.E-05 | 1.E+00 | 5.E-02 |

**Supplementary Table 7: Colocalization of association with SCAD and five other traits at SCAD top loci**

We display the results of approxiate Bayes factor colocalization analysis for the association with SCAD, on one hand, and 14 different traits on the other hand.

Top SNP (SCAD): SNP with lowest P-value for SCAD association in the set of common variants betwen the two studies. CHROM: Chromosome. POS: Position (hg19 coordinates). EA: Effect Allele. OA: Other Allele. Z: Z-score of the indicated top SNP. NSNPs: number of SNPs used for he analysis. R2: R2 of linkage desequilibrium between top SCAD SNP and top Trait SNP in the European population of 1000 Genomes reference panel PP.H0-H4.abf: Posterior probability that: H0: neither trait has a genetic association in the region; H1: only trait 1 has a genetic association in the region; H2: only trait 2 has a genetic association in the region; H3: both traits are associated, but with different causal variants; H4: both traits are associated and share a single causal variant.

| | | Top SCAD SNP | | | | | | | Top Trait SNP | | | | | | | | | | | | | |
|---|---|---|---|---|---|---|---|---|---|---|---|---|---|---|---|---|---|---|---|---|---|---|
| **Locus** | **Trait** | **rsID** | **EA** | **OA** | **SCAD Z** | **SCAD P** | **Trait Z** | **Trait P** | **rsID** | **EA** | **OA** | **SCAD Z** | **SCAD P** | **Trait Z** | **Trait P** | **R2** | **NSNPs** | **PP.H0.abf** | **PP.H1.abf** | **PP.H2.abf** | **PP.H3.abf** | **PP.H4.abf** |
| *FGGY-DT* | Intracranial Aneurysm (IA) | rs34370185 | T | G | 7.09 | 1.4E-12 | -1.0 | 3.0E-01 | rs61292499 | A | G | -0.06 | 9.5E-01 | -2.4 | 1.7E-02 | 0.00 | 872 | 0% | 96% | 0% | 2% | 2% |
| *F3* | Intracranial Aneurysm (IA) | rs1146473 | C | T | 5.82 | 5.8E-09 | 1.8 | 7.6E-02 | rs112663538 | T | C | -1.53 | 1.3E-01 | 2.6 | 9.0E-03 | 0.00 | 1288 | 0% | 90% | 0% | 4% | 6% |
| *FALEC/ADAMTSL4-AS2* | Intracranial Aneurysm (IA) | rs4970935 | T | C | -13.04 | 6.1E-39 | 0.9 | 3.5E-01 | rs17599629 | G | A | -1.89 | 5.8E-02 | -2.6 | 1.0E-02 | 0.02 | 831 | 0% | 96% | 0% | 2% | 2% |
| *AFAP1* | Intracranial Aneurysm (IA) | rs6828005 | A | G | -6.67 | 2.6E-11 | -3.0 | 2.4E-03 | rs11724720 | T | C | -5.45 | 5.4E-08 | -4.1 | 4.2E-05 | 0.61 | 1634 | 0% | 55% | 0% | 19% | 26% |
| *ZNF827* | Intracranial Aneurysm (IA) | rs1507928 | C | T | 5.75 | 8.9E-09 | -1.2 | 2.4E-01 | rs28626105 | C | T | 4.99 | 6.2E-07 | -3.6 | 2.9E-04 | 0.48 | 1075 | 0% | 83% | 0% | 7% | 10% |
| *ITGA1* | Intracranial Aneurysm (IA) | rs73102285 | G | A | 5.72 | 1.1E-08 | 2.1 | 3.3E-02 | rs1862191 | T | C | 0.21 | 8.3E-01 | 3.6 | 3.5E-04 | 0.00 | 1743 | 0% | 81% | 0% | 10% | 9% |
| *PHACTR1* | Intracranial Aneurysm (IA) | rs6925904 | G | A | -7.50 | 6.3E-14 | -0.9 | 3.7E-01 | rs2876277 | G | A | 0.19 | 8.5E-01 | -3.1 | 1.9E-03 | 0.00 | 1368 | 0% | 93% | 0% | 6% | 2% |
| *HTRA1* | Intracranial Aneurysm (IA) | rs2736923 | A | G | 5.47 | 4.6E-08 | 1.5 | 1.4E-01 | rs74765115 | G | T | -2.97 | 2.9E-03 | -3.8 | 1.3E-04 | 0.01 | 1152 | 0% | 70% | 0% | 23% | 6% |
| *SESN3/FAM76B* | Intracranial Aneurysm (IA) | rs2186739 | C | T | -7.23 | 4.8E-13 | 0.3 | 7.4E-01 | rs555556 | G | T | -1.49 | 1.4E-01 | -2.3 | 2.3E-02 | 0.00 | 1148 | 0% | 96% | 0% | 3% | 1% |
| *LRP1* | Intracranial Aneurysm (IA) | rs11172113 | C | T | -11.53 | 9.0E-31 | -0.1 | 9.0E-01 | rs147304335 | A | G | 0.45 | 6.5E-01 | 3.2 | 1.4E-03 | 0.00 | 823 | 0% | 95% | 0% | 4% | 1% |
| *ATP2B1* | Intracranial Aneurysm (IA) | rs1689040 | T | C | -6.17 | 7.0E-10 | -2.2 | 2.6E-02 | rs1590008 | T | C | 2.44 | 1.5E-02 | 3.4 | 5.7E-04 | 0.31 | 852 | 0% | 71% | 0% | 6% | 23% |
| *COL4A1* | Intracranial Aneurysm (IA) | rs7326444 | A | G | -6.46 | 1.0E-10 | 0.2 | 8.4E-01 | rs11619453 | A | C | 0.75 | 4.5E-01 | -2.2 | 2.5E-02 | 0.01 | 896 | 0% | 97% | 0% | 2% | 1% |
| *COL4A2* | Intracranial Aneurysm (IA) | rs11838776 | A | G | -8.74 | 2.5E-18 | -1.4 | 1.6E-01 | rs147233316 | A | C | 0.45 | 6.5E-01 | -3.3 | 1.0E-03 | 0.00 | 1012 | 0% | 89% | 0% | 7% | 4% |
| *FBN1* | Intracranial Aneurysm (IA) | rs655015 | C | T | 5.87 | 4.3E-09 | 0.8 | 4.0E-01 | rs12909029 | T | C | 0.88 | 3.8E-01 | 3.3 | 1.1E-03 | 0.04 | 411 | 0% | 95% | 0% | 3% | 2% |
| *THSD4* | Intracranial Aneurysm (IA) | rs28398466 | A | G | -6.46 | 1.1E-10 | -1.5 | 1.4E-01 | rs55940121 | A | G | 0.15 | 8.8E-01 | -3.4 | 6.8E-04 | 0.08 | 922 | 0% | 91% | 0% | 5% | 3% |
| *LINC00310/KCNE2* | Intracranial Aneurysm (IA) | rs9980618 | T | C | -8.35 | 7.2E-17 | 4.4 | 1.1E-05 | rs9977093 | A | G | -7.82 | 5.3E-15 | 4.7 | 3.2E-06 | 0.88 | 1298 | 0% | 1% | 0% | 1% | 99% |
| *SYN3* | Intracranial Aneurysm (IA) | rs137528 | A | G | -5.40 | 6.8E-08 | -0.9 | 3.5E-01 | rs78325826 | G | A | 0.34 | 7.3E-01 | -2.4 | 1.7E-02 | 0.00 | 1558 | 1% | 93% | 0% | 4% | 2% |
| *FGGY-DT* | Cervical Artery Dissection (CeAD) | rs34370185 | T | G | 7.09 | 1.4E-12 | 4.2 | 2.1E-05 | rs7551554 | G | A | 6.47 | 9.6E-11 | 4.6 | 3.6E-06 | 0.84 | 998 | 0% | 1% | 0% | 4% | 95% |
| *F3* | Cervical Artery Dissection (CeAD) | rs1146473 | C | T | 5.82 | 5.8E-09 | 0.3 | 7.3E-01 | rs17400584 | A | G | -1.63 | 1.0E-01 | -3.0 | 3.0E-03 | 0.01 | 1473 | 0% | 87% | 0% | 10% | 3% |
| *FALEC/ADAMTSL4-AS2* | Cervical Artery Dissection (CeAD) | rs1260387 | C | T | -12.21 | 3.2E-34 | -3.2 | 1.4E-03 | rs59898460 | C | T | -9.69 | 3.1E-22 | -4.0 | 5.3E-05 | 0.22 | 970 | 0% | 18% | 0% | 27% | 55% |
| *AFAP1* | Cervical Artery Dissection (CeAD) | rs6828005 | A | G | -6.67 | 2.6E-11 | -0.6 | 5.5E-01 | rs9330346 | C | T | 0.05 | 9.6E-01 | 3.5 | 5.4E-04 | 0.00 | 2028 | 0% | 80% | 0% | 18% | 2% |
| *ZNF827* | Cervical Artery Dissection (CeAD) | rs1507928 | C | T | 5.75 | 8.9E-09 | 1.5 | 1.2E-01 | rs10017224 | T | C | -0.07 | 9.4E-01 | 3.3 | 8.4E-04 | 0.00 | 1159 | 0% | 86% | 0% | 8% | 6% |
| *ITGA1* | Cervical Artery Dissection (CeAD) | rs73102285 | G | A | 5.72 | 1.1E-08 | 0.0 | 9.8E-01 | rs12697099 | A | T | -3.10 | 1.9E-03 | -3.6 | 2.6E-04 | 0.00 | 1979 | 0% | 85% | 0% | 13% | 2% |
| *PHACTR1* | Cervical Artery Dissection (CeAD) | rs6925904 | G | A | -7.50 | 6.3E-14 | -2.9 | 4.0E-03 | rs1937768 | G | A | -4.53 | 5.8E-06 | -3.7 | 1.8E-04 | 0.03 | 1565 | 0% | 33% | 0% | 23% | 44% |
| *HTRA1* | Cervical Artery Dissection (CeAD) | rs2736923 | A | G | 5.47 | 4.6E-08 | 0.9 | 3.4E-01 | rs7085188 | G | C | -2.15 | 3.1E-02 | -3.4 | 7.6E-04 | 0.01 | 1458 | 0% | 83% | 0% | 13% | 4% |
| *SESN3/FAM76B* | Cervical Artery Dissection (CeAD) | rs11021221 | A | T | 7.84 | 4.1E-15 | 0.7 | 4.7E-01 | rs11021389 | A | G | -1.24 | 2.1E-01 | 2.8 | 4.5E-03 | 0.00 | 1260 | 0% | 91% | 0% | 6% | 3% |
| *LRP1* | Cervical Artery Dissection (CeAD) | rs11172113 | C | T | -11.53 | 9.0E-31 | -5.4 | 5.1E-08 | rs11172113 | C | T | -11.53 | 9.0E-31 | -5.4 | 5.1E-08 | 1.00 | 951 | 0% | 0% | 0% | 0% | 100% |
| *ATP2B1* | Cervical Artery Dissection (CeAD) | rs1689040 | T | C | -6.17 | 7.0E-10 | 0.0 | 9.7E-01 | rs112914514 | G | A | 1.32 | 1.9E-01 | 2.7 | 6.2E-03 | 0.01 | 969 | 0% | 92% | 0% | 5% | 3% |
| *COL4A1* | Cervical Artery Dissection (CeAD) | rs7326444 | A | G | -6.46 | 1.0E-10 | -1.7 | 8.1E-02 | rs12584092 | T | C | 0.51 | 6.1E-01 | 3.7 | 2.6E-04 | 0.00 | 1092 | 0% | 76% | 0% | 16% | 7% |
| *COL4A2* | Cervical Artery Dissection (CeAD) | rs11838776 | A | G | -8.74 | 2.5E-18 | -1.1 | 2.8E-01 | rs34905765 | T | C | -1.81 | 7.0E-02 | 2.6 | 9.6E-03 | 0.00 | 1159 | 0% | 91% | 0% | 5% | 4% |
| *FBN1* | Cervical Artery Dissection (CeAD) | rs2437947 | A | T | 5.94 | 2.9E-09 | 0.7 | 4.9E-01 | rs7495591 | T | A | 0.39 | 6.9E-01 | 3.1 | 1.8E-03 | 0.05 | 506 | 0% | 92% | 0% | 4% | 4% |
| *THSD4* | Cervical Artery Dissection (CeAD) | rs10851839 | A | T | 6.56 | 5.5E-11 | 1.1 | 2.9E-01 | rs4357892 | C | A | 0.20 | 8.4E-01 | -2.8 | 5.7E-03 | 0.01 | 1090 | 0% | 91% | 0% | 5% | 3% |
| *LINC00310/KCNE2* | Cervical Artery Dissection (CeAD) | rs9980618 | T | C | -8.35 | 7.2E-17 | -2.9 | 3.5E-03 | rs9980618 | T | C | -8.35 | 7.2E-17 | -2.9 | 3.5E-03 | 1.00 | 1469 | 0% | 40% | 0% | 3% | 57% |
| *SYN3* | Cervical Artery Dissection (CeAD) | rs137528 | A | G | -5.40 | 6.8E-08 | -0.6 | 5.5E-01 | rs74662609 | T | C | -1.75 | 8.0E-02 | -3.2 | 1.6E-03 | 0.00 | 1758 | 1% | 84% | 0% | 12% | 4% |
| *FGGY-DT* | Diastolic Blood Pressure (DBP) | rs34370185 | T | G | 7.09 | 1.4E-12 | 5.0 | 7.0E-07 | rs12730750 | A | G | 5.32 | 1.1E-07 | 6.0 | 1.9E-09 | 0.52 | 1047 | 0% | 0% | 0% | 8% | 91% |
| *F3* | Diastolic Blood Pressure (DBP) | rs1146473 | C | T | 5.82 | 5.8E-09 | 0.9 | 3.7E-01 | rs17396055 | A | G | -0.13 | 9.0E-01 | -6.3 | 4.1E-10 | 0.01 | 1579 | 0% | 0% | 0% | 100% | 0% |
| *FALEC/ADAMTSL4-AS2* | Diastolic Blood Pressure (DBP) | rs4970935 | T | C | -13.04 | 6.1E-39 | -2.9 | 3.6E-03 | rs35392872 | A | G | 0.39 | 6.9E-01 | -3.8 | 1.2E-04 | 0.01 | 1035 | 0% | 89% | 0% | 4% | 8% |
| *AFAP1* | Diastolic Blood Pressure (DBP) | rs6828005 | A | G | -6.67 | 2.6E-11 | 0.4 | 6.7E-01 | rs7655880 | G | A | -1.75 | 8.0E-02 | 4.6 | 4.8E-06 | 0.00 | 2213 | 0% | 89% | 0% | 11% | 0% |
| *ZNF827* | Diastolic Blood Pressure (DBP) | rs1507928 | C | T | 5.75 | 8.9E-09 | -2.7 | 7.7E-03 | rs10022648 | G | A | 1.31 | 1.9E-01 | -5.1 | 3.1E-07 | 0.01 | 1247 | 0% | 51% | 0% | 41% | 8% |
| *ITGA1* | Diastolic Blood Pressure (DBP) | rs73102285 | G | A | 5.72 | 1.1E-08 | -0.5 | 6.5E-01 | rs1645761 | T | C | -1.48 | 1.4E-01 | 4.0 | 7.3E-05 | 0.00 | 2102 | 0% | 93% | 0% | 6% | 0% |
| *PHACTR1* | Diastolic Blood Pressure (DBP) | rs6925904 | G | A | -7.50 | 6.3E-14 | -0.2 | 8.2E-01 | rs17679286 | G | A | 2.74 | 6.2E-03 | 2.8 | 4.6E-03 | 0.04 | 1611 | 0% | 99% | 0% | 1% | 0% |
| *HTRA1* | Diastolic Blood Pressure (DBP) | rs2736923 | A | G | 5.47 | 4.6E-08 | -3.5 | 4.8E-04 | rs10490923 | A | G | -3.00 | 2.7E-03 | 5.9 | 5.0E-09 | 0.02 | 1553 | 0% | 0% | 0% | 99% | 0% |
| *SESN3/FAM76B* | Diastolic Blood Pressure (DBP) | rs11021221 | A | T | 7.84 | 4.1E-15 | -8.1 | 6.9E-16 | rs11021221 | A | T | 7.84 | 4.1E-15 | -8.1 | 6.9E-16 | 1.00 | 1360 | 0% | 0% | 0% | 0% | 100% |
| *LRP1* | Diastolic Blood Pressure (DBP) | rs11172113 | C | T | -11.53 | 9.0E-31 | -4.0 | 5.6E-05 | rs2958124 | A | C | 2.00 | 4.5E-02 | 7.7 | 2.0E-14 | 0.01 | 1002 | 0% | 0% | 0% | 100% | 0% |
| *ATP2B1* | Diastolic Blood Pressure (DBP) | rs1689040 | T | C | -6.17 | 7.0E-10 | -16.5 | 2.8E-61 | rs2681485 | A | G | 5.99 | 2.1E-09 | 16.7 | 1.3E-62 | 1.00 | 1023 | 0% | 0% | 0% | 2% | 98% |
| *COL4A1* | Diastolic Blood Pressure (DBP) | rs7326444 | A | G | -6.46 | 1.0E-10 | 5.5 | 5.0E-08 | rs9559749 | A | G | 0.73 | 4.6E-01 | 5.7 | 1.4E-08 | 0.00 | 1147 | 0% | 0% | 0% | 5% | 95% |
| *COL4A2* | Diastolic Blood Pressure (DBP) | rs11838776 | A | G | -8.74 | 2.5E-18 | 5.1 | 2.6E-07 | rs55940034 | G | A | -8.56 | 1.2E-17 | 5.3 | 1.3E-07 | 0.94 | 1222 | 0% | 0% | 0% | 0% | 100% |
| *FBN1* | Diastolic Blood Pressure (DBP) | rs2437947 | A | T | 5.94 | 2.9E-09 | 3.7 | 2.5E-04 | rs2245752 | A | C | -2.61 | 9.0E-03 | -5.4 | 8.3E-08 | 0.31 | 527 | 0% | 2% | 0% | 95% | 3% |
| *THSD4* | Diastolic Blood Pressure (DBP) | rs10851839 | A | T | 6.56 | 5.5E-11 | 8.7 | 2.4E-18 | rs11853359 | A | G | -5.54 | 3.0E-08 | -9.1 | 1.3E-19 | 0.96 | 1140 | 0% | 0% | 0% | 3% | 97% |
| *LINC00310/KCNE2* | Diastolic Blood Pressure (DBP) | rs9980618 | T | C | -8.35 | 7.2E-17 | 4.2 | 2.5E-05 | rs71326977 | A | C | 1.29 | 2.0E-01 | -5.9 | 2.8E-09 | 0.01 | 1517 | 0% | 1% | 0% | 92% | 7% |
| *SYN3* | Diastolic Blood Pressure (DBP) | rs137528 | A | G | -5.40 | 6.8E-08 | 0.0 | 9.8E-01 | rs16990642 | A | T | 0.29 | 7.7E-01 | 3.2 | 1.4E-03 | 0.00 | 1861 | 1% | 98% | 0% | 1% | 0% |

| | | | | | | | | | | | | | | | | | | | | | | |
|---|---|---|---|---|---|---|---|---|---|---|---|---|---|---|---|---|---|---|---|---|---|---|
| *FGGY-DT* | Systolic Blood Pressure (PP) | rs34370185 | T | G | 7.09 | 1.4E-12 | -8.0 | 1.3E-15 | rs12758643 | T | C | 5.06 | 4.0E-07 | -8.3 | 1.1E-16 | 0.96 | 1045 | 0% | 0% | 0% | 8% | 92% |
| *F3* | Systolic Blood Pressure (PP) | rs1146473 | C | T | 5.82 | 5.8E-09 | 0.9 | 3.8E-01 | rs75088623 | G | A | -0.60 | 5.5E-01 | -5.8 | 5.1E-09 | 0.02 | 1577 | 0% | 1% | 0% | 99% | 0% |
| *FALEC/ADAMTSL4-AS2* | Systolic Blood Pressure (PP) | rs4970935 | T | C | -13.04 | 6.1E-39 | 2.0 | 5.1E-02 | rs11585169 | A | T | -1.39 | 1.6E-01 | 5.8 | 5.3E-09 | 0.04 | 1035 | 0% | 0% | 0% | 100% | 0% |
| *AFAP1* | Systolic Blood Pressure (PP) | rs6828005 | A | G | -6.67 | 2.6E-11 | 3.3 | 8.7E-04 | rs4478172 | C | A | 5.02 | 5.3E-07 | -4.8 | 1.5E-06 | 0.04 | 2202 | 0% | 31% | 0% | 30% | 39% |
| *ZNF827* | Systolic Blood Pressure (PP) | rs1507928 | C | T | 5.75 | 8.9E-09 | -6.3 | 3.8E-10 | rs13128814 | A | G | 5.53 | 3.2E-08 | -7.1 | 1.2E-12 | 0.71 | 1244 | 0% | 0% | 0% | 2% | 98% |
| *ITGA1* | Systolic Blood Pressure (PP) | rs73102285 | G | A | 5.72 | 1.1E-08 | 3.3 | 9.2E-04 | rs74344272 | T | A | 1.27 | 2.0E-01 | 4.3 | 1.9E-05 | 0.00 | 2098 | 0% | 62% | 0% | 14% | 24% |
| *PHACTR1* | Systolic Blood Pressure (PP) | rs6925904 | G | A | -7.50 | 6.3E-14 | -6.7 | 1.7E-11 | rs2876301 | T | C | -7.24 | 4.2E-13 | -6.9 | 4.2E-12 | 0.99 | 1610 | 0% | 0% | 0% | 3% | 97% |
| *HTRA1* | Systolic Blood Pressure (PP) | rs2736923 | A | G | 5.47 | 4.6E-08 | -1.8 | 6.9E-02 | rs72834453 | G | T | -2.73 | 6.4E-03 | 7.0 | 3.0E-12 | 0.02 | 1549 | 0% | 0% | 0% | 99% | 0% |
| *SESN3/FAM76B* | Systolic Blood Pressure (PP) | rs11021221 | A | T | 7.84 | 4.1E-15 | 0.0 | 9.6E-01 | rs1047731 | G | A | 3.36 | 7.7E-04 | 2.6 | 8.2E-03 | 0.00 | 1357 | 0% | 99% | 0% | 1% | 0% |
| *LRP1* | Systolic Blood Pressure (PP) | rs11172113 | C | T | -11.53 | 9.0E-31 | 1.6 | 1.2E-01 | rs1296041 | C | T | -0.35 | 7.2E-01 | 4.8 | 1.4E-06 | 0.00 | 1002 | 0% | 31% | 0% | 69% | 0% |
| *ATP2B1* | Systolic Blood Pressure (PP) | rs1689040 | T | C | -6.17 | 7.0E-10 | -18.5 | 3.7E-76 | rs17249754 | A | G | -5.61 | 2.0E-08 | -21.0 | 1.3E-97 | 0.28 | 1021 | 0% | 0% | 0% | 8% | 92% |
| *COL4A1* | Systolic Blood Pressure (PP) | rs7326444 | A | G | -6.46 | 1.0E-10 | 0.7 | 4.9E-01 | rs4771653 | T | C | 0.48 | 6.3E-01 | -4.9 | 9.9E-07 | 0.00 | 1145 | 0% | 47% | 0% | 53% | 0% |
| *COL4A2* | Systolic Blood Pressure (PP) | rs11838776 | A | G | -8.74 | 2.5E-18 | 1.1 | 2.6E-01 | rs9515270 | T | C | -2.47 | 1.4E-02 | 5.3 | 1.0E-07 | 0.00 | 1220 | 0% | 8% | 0% | 92% | 0% |
| *FBN1* | Systolic Blood Pressure (PP) | rs2437947 | A | T | 5.94 | 2.9E-09 | -7.9 | 2.6E-15 | rs2437947 | A | T | 5.94 | 2.9E-09 | -7.9 | 2.6E-15 | 1.00 | 527 | 0% | 0% | 0% | 3% | 97% |
| *THSD4* | Systolic Blood Pressure (PP) | rs10851839 | A | T | 6.56 | 5.5E-11 | 0.7 | 4.6E-01 | rs985868 | G | A | -0.61 | 5.4E-01 | 3.4 | 7.8E-04 | 0.00 | 1138 | 0% | 99% | 0% | 1% | 0% |
| *LINC00310/KCNE2* | Systolic Blood Pressure (PP) | rs9980618 | T | C | -8.35 | 7.2E-17 | -2.4 | 1.4E-02 | rs2834257 | A | G | -1.34 | 1.8E-01 | 5.1 | 2.8E-07 | 0.00 | 1516 | 0% | 21% | 0% | 78% | 1% |
| *SYN3* | Systolic Blood Pressure (PP) | rs137528 | A | G | -5.40 | 6.8E-08 | 1.3 | 2.0E-01 | rs2049948 | G | A | 1.20 | 2.3E-01 | -3.2 | 1.2E-03 | 0.00 | 1857 | 1% | 98% | 0% | 1% | 0% |
| *FGGY-DT* | Fibromuscular Dysplasia | rs34370185 | T | G | 7.09 | 1.4E-12 | 1.4 | 1.5E-01 | rs10493259 | C | T | -0.65 | 5.2E-01 | -2.9 | 3.6E-03 | 0.02 | 978 | 0% | 86% | 0% | 8% | 6% |
| *F3* | Fibromuscular Dysplasia | rs1146473 | C | T | 5.82 | 5.8E-09 | 0.8 | 4.0E-01 | rs859068 | T | C | -1.84 | 6.5E-02 | -2.8 | 5.1E-03 | 0.00 | 1523 | 0% | 86% | 0% | 10% | 3% |
| *FALEC/ADAMTSL4-AS2* | Fibromuscular Dysplasia | rs4446975 | T | C | -11.38 | 5.3E-30 | -3.1 | 2.3E-03 | rs11204662 | G | T | -10.65 | 1.7E-26 | -3.5 | 4.5E-04 | 0.89 | 808 | 0% | 30% | 0% | 7% | 63% |
| *AFAP1* | Fibromuscular Dysplasia | rs6828005 | A | G | -6.67 | 2.6E-11 | -0.8 | 4.2E-01 | rs56411722 | C | T | -5.04 | 4.5E-07 | -3.5 | 5.0E-04 | 0.27 | 2031 | 0% | 72% | 0% | 25% | 3% |
| *ZNF827* | Fibromuscular Dysplasia | rs1507928 | C | T | 5.75 | 8.9E-09 | 3.3 | 8.1E-04 | rs4544728 | T | C | 3.88 | 1.1E-04 | 4.1 | 3.6E-05 | 0.26 | 1079 | 0% | 14% | 0% | 17% | 69% |
| *ITGA1* | Fibromuscular Dysplasia | rs73102285 | G | A | 5.72 | 1.1E-08 | 1.0 | 3.3E-01 | rs62357202 | A | G | -2.35 | 1.9E-02 | -2.8 | 4.8E-03 | 0.00 | 2040 | 0% | 83% | 0% | 13% | 4% |
| *PHACTR1* | Fibromuscular Dysplasia | rs6925904 | G | A | -7.50 | 6.3E-14 | -4.6 | 4.3E-06 | rs7751826 | T | C | -7.10 | 1.3E-12 | -4.7 | 2.1E-06 | 0.97 | 1540 | 0% | 0% | 0% | 2% | 97% |
| *HTRA1* | Fibromuscular Dysplasia | rs2736923 | A | G | 5.47 | 4.6E-08 | 2.2 | 2.5E-02 | rs3817285 | C | T | -0.12 | 9.1E-01 | -3.5 | 5.3E-04 | 0.00 | 1428 | 0% | 71% | 0% | 10% | 19% |
| *SESN3/FAM76B* | Fibromuscular Dysplasia | rs11021221 | A | T | 7.84 | 4.1E-15 | -0.3 | 8.0E-01 | rs75887387 | T | G | -0.81 | 4.2E-01 | -2.5 | 1.1E-02 | 0.00 | 1202 | 0% | 91% | 0% | 6% | 3% |
| *LRP1* | Fibromuscular Dysplasia | rs11172113 | C | T | -11.53 | 9.0E-31 | -6.4 | 2.0E-10 | rs11172113 | C | T | -11.53 | 9.0E-31 | -6.4 | 2.0E-10 | 1.00 | 746 | 0% | 0% | 0% | 0% | 100% |
| *ATP2B1* | Fibromuscular Dysplasia | rs1689040 | T | C | -6.17 | 7.0E-10 | -3.6 | 3.1E-04 | rs2681492 | C | T | -5.51 | 3.6E-08 | -5.6 | 1.7E-08 | 0.27 | 762 | 0% | 0% | 0% | 10% | 90% |
| *COL4A1* | Fibromuscular Dysplasia | rs7326444 | A | G | -6.46 | 1.0E-10 | -1.3 | 1.9E-01 | rs80343422 | G | A | 0.09 | 9.3E-01 | -2.4 | 1.5E-02 | 0.00 | 1040 | 0% | 91% | 0% | 5% | 5% |
| *COL4A2* | Fibromuscular Dysplasia | rs11838776 | A | G | -8.74 | 2.5E-18 | -3.7 | 1.8E-04 | rs55940034 | G | A | -8.56 | 1.2E-17 | -3.8 | 1.4E-04 | 0.94 | 1130 | 0% | 5% | 0% | 1% | 94% |
| *FBN1* | Fibromuscular Dysplasia | rs2437947 | A | T | 5.94 | 2.9E-09 | 1.2 | 2.4E-01 | rs9972344 | G | T | -1.92 | 5.4E-02 | -3.0 | 2.5E-03 | 0.00 | 393 | 0% | 90% | 0% | 4% | 6% |
| *THSD4* | Fibromuscular Dysplasia | rs10851839 | A | T | 6.56 | 5.5E-11 | 3.2 | 1.3E-03 | rs11632680 | G | T | 2.91 | 3.6E-03 | 4.0 | 6.3E-05 | 0.21 | 1089 | 0% | 26% | 0% | 7% | 66% |
| *LINC00310/KCNE2* | Fibromuscular Dysplasia | rs9980618 | T | C | -8.35 | 7.2E-17 | -3.0 | 2.4E-03 | rs3746861 | T | C | 2.17 | 3.0E-02 | 3.3 | 1.1E-03 | 0.01 | 1481 | 0% | 34% | 0% | 6% | 60% |
| *SYN3* | Fibromuscular Dysplasia | rs137528 | A | G | -5.40 | 6.8E-08 | -1.1 | 2.7E-01 | rs142100390 | A | C | -3.28 | 1.1E-03 | -3.2 | 1.2E-03 | 0.19 | 1734 | 1% | 81% | 0% | 15% | 4% |
| *FGGY-DT* | Coronary Artery Disease (CAD) | rs34370185 | T | G | 7.09 | 1.4E-12 | -2.7 | 6.1E-03 | rs932773 | C | G | 2.06 | 4.0E-02 | 4.1 | 4.1E-05 | 0.01 | 947 | 0% | 76% | 0% | 12% | 12% |
| *F3* | Coronary Artery Disease (CAD) | rs1146473 | C | T | 5.82 | 5.8E-09 | -0.1 | 9.4E-01 | rs17398377 | C | T | 1.43 | 1.5E-01 | 2.7 | 7.1E-03 | 0.01 | 1440 | 0% | 98% | 0% | 1% | 0% |
| *FALEC/ADAMTSL4-AS2* | Coronary Artery Disease (CAD) | rs4970935 | T | C | -13.04 | 6.1E-39 | 4.2 | 2.2E-05 | rs7549723 | T | C | -3.31 | 9.5E-04 | 5.2 | 1.8E-07 | 0.10 | 868 | 0% | 1% | 0% | 60% | 39% |
| *AFAP1* | Coronary Artery Disease (CAD) | rs6828005 | A | G | -6.67 | 2.6E-11 | 1.8 | 7.4E-02 | rs9968354 | C | T | 1.01 | 3.1E-01 | -3.7 | 2.1E-04 | 0.08 | 1658 | 0% | 92% | 0% | 6% | 1% |
| *ZNF827* | Coronary Artery Disease (CAD) | rs1507928 | C | T | 5.75 | 8.9E-09 | -3.7 | 2.4E-04 | rs4345206 | C | T | 5.01 | 5.6E-07 | -4.1 | 4.1E-05 | 0.63 | 871 | 0% | 43% | 0% | 3% | 53% |
| *ITGA1* | Coronary Artery Disease (CAD) | rs73102285 | G | A | 5.72 | 1.1E-08 | 0.3 | 7.8E-01 | rs62357230 | A | G | -1.37 | 1.7E-01 | 3.9 | 1.2E-04 | 0.00 | 1719 | 0% | 85% | 0% | 15% | 0% |
| *PHACTR1* | Coronary Artery Disease (CAD) | rs6925904 | G | A | -7.50 | 6.3E-14 | 11.5 | 2.4E-30 | rs6925904 | G | A | -7.50 | 6.3E-14 | 11.5 | 2.4E-30 | 1.00 | 1227 | 0% | 0% | 0% | 0% | 100% |
| *HTRA1* | Coronary Artery Disease (CAD) | rs72631113 | T | C | 5.42 | 5.9E-08 | -4.9 | 1.1E-06 | rs77494534 | T | C | -0.05 | 9.6E-01 | 5.6 | 1.7E-08 | 0.02 | 1332 | 0% | 0% | 0% | 16% | 84% |
| *SESN3/FAM76B* | Coronary Artery Disease (CAD) | rs11021221 | A | T | 7.84 | 4.1E-15 | -2.2 | 3.0E-02 | rs7938589 | A | G | -0.76 | 4.5E-01 | -3.6 | 3.0E-04 | 0.00 | 1107 | 0% | 92% | 0% | 4% | 4% |
| *LRP1* | Coronary Artery Disease (CAD) | rs11172113 | C | T | -11.53 | 9.0E-31 | 3.2 | 1.2E-03 | rs507562 | G | C | 1.76 | 7.9E-02 | 4.7 | 3.0E-06 | 0.00 | 775 | 0% | 27% | 0% | 57% | 16% |
| *ATP2B1* | Coronary Artery Disease (CAD) | rs1689040 | T | C | -6.17 | 7.0E-10 | 3.3 | 8.9E-04 | rs12579302 | G | A | -5.62 | 2.0E-08 | 6.4 | 1.2E-10 | 0.28 | 962 | 0% | 0% | 0% | 9% | 91% |
| *COL4A1* | Coronary Artery Disease (CAD) | rs7326444 | A | G | -6.46 | 1.0E-10 | 6.5 | 9.4E-11 | rs1000989 | C | T | -5.08 | 3.8E-07 | 7.0 | 2.3E-12 | 0.89 | 1076 | 0% | 0% | 0% | 15% | 85% |
| *COL4A2* | Coronary Artery Disease (CAD) | rs11838776 | A | G | -8.74 | 2.5E-18 | 6.6 | 3.6E-11 | rs11838776 | A | G | -8.74 | 2.5E-18 | 6.6 | 3.6E-11 | 1.00 | 1053 | 0% | 0% | 0% | 0% | 100% |
| *FBN1* | Coronary Artery Disease (CAD) | rs2437947 | A | T | 5.94 | 2.9E-09 | -1.9 | 5.2E-02 | rs11638254 | A | T | -0.11 | 9.2E-01 | 2.5 | 1.4E-02 | 0.01 | 367 | 0% | 96% | 0% | 0% | 4% |
| *THSD4* | Coronary Artery Disease (CAD) | rs10851839 | A | T | 6.56 | 5.5E-11 | -1.6 | 1.2E-01 | rs4300598 | T | C | 0.01 | 9.9E-01 | 2.5 | 1.1E-02 | 0.00 | 1050 | 0% | 98% | 0% | 1% | 1% |
| *LINC00310/KCNE2* | Coronary Artery Disease (CAD) | rs9980618 | T | C | -8.35 | 7.2E-17 | 9.7 | 3.0E-22 | rs9980618 | T | C | -8.35 | 7.2E-17 | 9.7 | 3.0E-22 | 1.00 | 1351 | 0% | 0% | 0% | 0% | 100% |
| *SYN3* | Coronary Artery Disease (CAD) | rs137528 | A | G | -5.40 | 6.8E-08 | 2.7 | 7.7E-03 | rs5754240 | A | G | 2.01 | 4.4E-02 | -4.3 | 2.1E-05 | 0.05 | 1652 | 0% | 46% | 0% | 9% | 44% |
| *FGGY-DT* | Any stroke (AS) | rs34370185 | T | G | 7.09 | 1.4E-12 | 0.0 | 9.8E-01 | rs12738848 | G | T | -0.21 | 8.4E-01 | 3.0 | 2.9E-03 | 0.00 | 1040 | 0% | 98% | 0% | 2% | 1% |
| *F3* | Any stroke (AS) | rs1146473 | C | T | 5.82 | 5.8E-09 | 0.3 | 8.0E-01 | rs77568287 | A | G | 0.16 | 8.7E-01 | -3.3 | 8.6E-04 | 0.00 | 1560 | 0% | 96% | 0% | 4% | 1% |
| *FALEC/ADAMTSL4-AS2* | Any stroke (AS) | rs1260387 | C | T | -12.21 | 3.2E-34 | 1.2 | 2.5E-01 | rs12403795 | T | C | 8.04 | 8.9E-16 | -2.3 | 2.0E-02 | 0.41 | 1028 | 0% | 98% | 0% | 1% | 1% |
| *AFAP1* | Any stroke (AS) | rs6828005 | A | G | -6.67 | 2.6E-11 | 0.5 | 6.5E-01 | rs67667723 | C | A | -0.80 | 4.3E-01 | -3.5 | 5.8E-04 | 0.00 | 2204 | 0% | 94% | 0% | 5% | 1% |
| *ZNF827* | Any stroke (AS) | rs1507928 | C | T | 5.75 | 8.9E-09 | 0.2 | 8.2E-01 | rs11733094 | A | G | 0.44 | 6.6E-01 | 3.0 | 2.6E-03 | 0.01 | 1232 | 0% | 96% | 0% | 3% | 0% |
| *ITGA1* | Any stroke (AS) | rs73102285 | G | A | 5.72 | 1.1E-08 | -3.2 | 1.5E-03 | rs16880353 | G | A | 5.63 | 1.8E-08 | -3.4 | 6.0E-04 | 0.96 | 2088 | 0% | 52% | 0% | 3% | 44% |
| *PHACTR1* | Any stroke (AS) | rs6925904 | G | A | -7.50 | 6.3E-14 | -0.9 | 3.9E-01 | rs6900427 | A | G | 2.80 | 5.1E-03 | 2.1 | 3.2E-02 | 0.02 | 1608 | 0% | 97% | 0% | 2% | 1% |
| *HTRA1* | Any stroke (AS) | rs2736923 | A | G | 5.47 | 4.6E-08 | 0.0 | 9.7E-01 | rs2284665 | T | G | 4.95 | 7.6E-07 | -5.4 | 6.0E-08 | 0.02 | 1549 | 0% | 0% | 0% | 18% | 82% |
| *SESN3/FAM76B* | Any stroke (AS) | rs11021221 | A | T | 7.84 | 4.1E-15 | -1.8 | 7.4E-02 | rs7110456 | C | T | 0.05 | 9.6E-01 | -2.8 | 5.3E-03 | 0.00 | 1353 | 0% | 95% | 0% | 2% | 3% |
| *LRP1* | Any stroke (AS) | rs11172113 | C | T | -11.53 | 9.0E-31 | 1.9 | 6.0E-02 | rs2229717 | T | G | -1.73 | 8.3E-02 | 4.0 | 7.4E-05 | 0.00 | 993 | 0% | 92% | 0% | 5% | 3% |

| | | | | | | | | | | | | | | | | | | | | | | |
|---|---|---|---|---|---|---|---|---|---|---|---|---|---|---|---|---|---|---|---|---|---|---|
| *ATP2B1* | Any stroke (AS) | rs1689040 | T | C | -6.17 | 7.0E-10 | -1.9 | 5.9E-02 | rs7294375 | G | T | 3.92 | 9.1E-05 | 5.1 | 3.3E-07 | 0.10 | 1019 | 0% | 3% | 0% | 93% | 4% |
| *COL4A1* | Any stroke (AS) | rs7326444 | A | G | -6.46 | 1.0E-10 | 3.9 | 8.6E-05 | rs9521635 | C | T | -6.05 | 1.4E-09 | 4.5 | 9.4E-06 | 0.92 | 1142 | 0% | 5% | 0% | 5% | 90% |
| *COL4A2* | Any stroke (AS) | rs11838776 | A | G | -8.74 | 2.5E-18 | 2.4 | 1.6E-02 | rs12865767 | T | A | 0.32 | 7.5E-01 | 2.9 | 3.4E-03 | 0.00 | 1210 | 0% | 90% | 0% | 2% | 8% |
| *FBN1* | Any stroke (AS) | rs2437947 | A | T | 5.94 | 2.9E-09 | -0.4 | 7.0E-01 | rs7169672 | T | C | -0.53 | 6.0E-01 | 2.0 | 4.9E-02 | 0.01 | 522 | 0% | 99% | 0% | 0% | 1% |
| *THSD4* | Any stroke (AS) | rs10851839 | A | T | 6.56 | 5.5E-11 | 0.8 | 4.3E-01 | rs1866920 | C | T | 2.84 | 4.5E-03 | 3.0 | 2.8E-03 | 0.12 | 1131 | 0% | 98% | 0% | 2% | 1% |
| *LINC00310/KCNE2* | Any stroke (AS) | rs9980618 | T | C | -8.35 | 7.2E-17 | -1.6 | 1.1E-01 | rs34545319 | G | A | 0.19 | 8.5E-01 | 2.9 | 4.3E-03 | 0.01 | 1516 | 0% | 95% | 0% | 3% | 2% |
| *SYN3* | Any stroke (AS) | rs137528 | A | G | -5.40 | 6.8E-08 | 0.1 | 9.3E-01 | rs7289631 | A | G | -1.55 | 1.2E-01 | -3.8 | 1.6E-04 | 0.18 | 1852 | 1% | 87% | 0% | 12% | 1% |
| *FGGY-DT* | Cardioembolic Stroke (CES) | rs34370185 | T | G | 7.09 | 1.4E-12 | 0.0 | 9.8E-01 | rs6664131 | A | C | 1.31 | 1.9E-01 | -3.1 | 1.9E-03 | 0.00 | 1040 | 0% | 94% | 0% | 5% | 1% |
| *F3* | Cardioembolic Stroke (CES) | rs1146473 | C | T | 5.82 | 5.8E-09 | 0.3 | 7.7E-01 | rs79658099 | C | T | -0.60 | 5.5E-01 | 3.0 | 3.0E-03 | 0.00 | 1560 | 0% | 93% | 0% | 6% | 1% |
| *FALEC/ADAMTSL4-AS2* | Cardioembolic Stroke (CES) | rs1260387 | C | T | -12.21 | 3.2E-34 | 1.8 | 6.7E-02 | rs12134682 | C | G | -2.14 | 3.3E-02 | 3.3 | 8.0E-04 | 0.05 | 1028 | 0% | 82% | 0% | 13% | 5% |
| *AFAP1* | Cardioembolic Stroke (CES) | rs6828005 | A | G | -6.67 | 2.6E-11 | 0.2 | 8.5E-01 | rs6848741 | G | A | -0.89 | 3.7E-01 | -3.2 | 1.5E-03 | 0.01 | 2204 | 0% | 91% | 0% | 7% | 1% |
| *ZNF827* | Cardioembolic Stroke (CES) | rs1507928 | C | T | 5.75 | 8.9E-09 | 1.1 | 2.5E-01 | rs72729816 | A | C | -0.34 | 7.4E-01 | 3.0 | 2.4E-03 | 0.00 | 1232 | 0% | 94% | 0% | 4% | 2% |
| *ITGA1* | Cardioembolic Stroke (CES) | rs73102285 | G | A | 5.72 | 1.1E-08 | -1.8 | 7.4E-02 | rs2126952 | C | T | -2.75 | 6.0E-03 | 3.8 | 1.4E-04 | 0.44 | 2088 | 0% | 83% | 0% | 11% | 5% |
| *PHACTR1* | Cardioembolic Stroke (CES) | rs6925904 | G | A | -7.50 | 6.3E-14 | -0.4 | 7.1E-01 | rs116314410 | A | T | 1.69 | 9.1E-02 | 2.9 | 3.3E-03 | 0.00 | 1608 | 0% | 95% | 0% | 4% | 1% |
| *HTRA1* | Cardioembolic Stroke (CES) | rs2736923 | A | G | 5.47 | 4.6E-08 | -1.2 | 2.3E-01 | rs1891114 | G | A | 0.35 | 7.3E-01 | 3.0 | 2.9E-03 | 0.00 | 1549 | 0% | 91% | 0% | 5% | 4% |
| *SESN3/FAM76B* | Cardioembolic Stroke (CES) | rs11021221 | A | T | 7.84 | 4.1E-15 | 0.6 | 5.5E-01 | rs10831314 | T | C | 2.67 | 7.5E-03 | 3.5 | 4.1E-04 | 0.00 | 1353 | 0% | 91% | 0% | 7% | 1% |
| *LRP1* | Cardioembolic Stroke (CES) | rs11172113 | C | T | -11.53 | 9.0E-31 | -0.3 | 7.7E-01 | rs2229717 | T | G | -1.73 | 8.3E-02 | 3.3 | 1.1E-03 | 0.00 | 993 | 0% | 97% | 0% | 2% | 1% |
| *ATP2B1* | Cardioembolic Stroke (CES) | rs1689040 | T | C | -6.17 | 7.0E-10 | -1.3 | 1.8E-01 | rs7960337 | C | T | 4.87 | 1.1E-06 | 2.4 | 1.4E-02 | 0.09 | 1019 | 0% | 95% | 0% | 3% | 2% |
| *COL4A1* | Cardioembolic Stroke (CES) | rs7326444 | A | G | -6.46 | 1.0E-10 | 0.6 | 5.6E-01 | rs681777 | C | G | 0.63 | 5.3E-01 | -2.9 | 3.3E-03 | 0.00 | 1142 | 0% | 94% | 0% | 4% | 1% |
| *COL4A2* | Cardioembolic Stroke (CES) | rs11838776 | A | G | -8.74 | 2.5E-18 | 0.7 | 4.9E-01 | rs9588247 | T | G | -1.81 | 7.0E-02 | 2.9 | 3.6E-03 | 0.00 | 1210 | 0% | 95% | 0% | 4% | 1% |
| *FBN1* | Cardioembolic Stroke (CES) | rs2437947 | A | T | 5.94 | 2.9E-09 | 1.6 | 1.0E-01 | rs74773702 | C | T | -0.43 | 6.7E-01 | 2.0 | 4.4E-02 | 0.01 | 522 | 0% | 94% | 0% | 1% | 5% |
| *THSD4* | Cardioembolic Stroke (CES) | rs10851839 | A | T | 6.56 | 5.5E-11 | -0.9 | 3.5E-01 | rs28693764 | A | C | 0.19 | 8.4E-01 | -2.5 | 1.1E-02 | 0.00 | 1131 | 0% | 96% | 0% | 2% | 2% |
| *LINC00310/KCNE2* | Cardioembolic Stroke (CES) | rs9980618 | T | C | -8.35 | 7.2E-17 | 1.0 | 3.0E-01 | rs41314677 | G | A | -0.86 | 3.9E-01 | 2.9 | 3.6E-03 | 0.00 | 1516 | 0% | 93% | 0% | 5% | 2% |
| *SYN3* | Cardioembolic Stroke (CES) | rs137528 | A | G | -5.40 | 6.8E-08 | 1.0 | 3.0E-01 | rs9621514 | T | C | 0.75 | 4.5E-01 | 3.6 | 3.8E-04 | 0.00 | 1852 | 1% | 85% | 0% | 13% | 2% |
| *FGGY-DT* | Small Vessel Stroke (SVS) | rs34370185 | T | G | 7.09 | 1.4E-12 | -0.1 | 9.3E-01 | rs114186484 | T | C | -1.01 | 3.1E-01 | 2.6 | 9.6E-03 | 0.01 | 1040 | 0% | 96% | 0% | 3% | 1% |
| *F3* | Small Vessel Stroke (SVS) | rs1146473 | C | T | 5.82 | 5.8E-09 | 0.2 | 8.4E-01 | rs6675132 | T | G | -1.21 | 2.3E-01 | -2.7 | 7.1E-03 | 0.00 | 1560 | 0% | 93% | 0% | 6% | 1% |
| *FALEC/ADAMTSL4-AS2* | Small Vessel Stroke (SVS) | rs1260387 | C | T | -12.21 | 3.2E-34 | -0.4 | 7.0E-01 | rs4970966 | T | G | 2.56 | 1.1E-02 | 2.4 | 1.8E-02 | 0.01 | 1028 | 0% | 97% | 0% | 2% | 1% |
| *AFAP1* | Small Vessel Stroke (SVS) | rs6828005 | A | G | -6.67 | 2.6E-11 | -0.1 | 9.5E-01 | rs4643793 | T | C | 0.01 | 9.9E-01 | 3.8 | 1.4E-04 | 0.00 | 2204 | 0% | 85% | 0% | 14% | 1% |
| *ZNF827* | Small Vessel Stroke (SVS) | rs1507928 | C | T | 5.75 | 8.9E-09 | -0.5 | 6.4E-01 | rs7694604 | G | T | -0.57 | 5.7E-01 | 4.5 | 8.6E-06 | 0.00 | 1232 | 0% | 21% | 0% | 79% | 0% |
| *ITGA1* | Small Vessel Stroke (SVS) | rs73102285 | G | A | 5.72 | 1.1E-08 | -2.5 | 1.3E-02 | rs72752528 | T | C | 0.54 | 5.9E-01 | 2.8 | 5.1E-03 | 0.00 | 2088 | 0% | 73% | 0% | 8% | 19% |
| *PHACTR1* | Small Vessel Stroke (SVS) | rs6925904 | G | A | -7.50 | 6.3E-14 | -1.8 | 7.7E-02 | rs9471949 | C | T | -0.03 | 9.7E-01 | 3.0 | 3.0E-03 | 0.00 | 1608 | 0% | 91% | 0% | 5% | 4% |
| *HTRA1* | Small Vessel Stroke (SVS) | rs2736923 | A | G | 5.47 | 4.6E-08 | -1.6 | 1.2E-01 | rs2142308 | C | G | 4.35 | 1.3E-05 | -4.5 | 5.6E-06 | 0.02 | 1549 | 0% | 3% | 0% | 10% | 87% |
| *SESN3/FAM76B* | Small Vessel Stroke (SVS) | rs11021221 | A | T | 7.84 | 4.1E-15 | -2.5 | 1.3E-02 | rs4384353 | C | T | -4.92 | 8.9E-07 | 3.2 | 1.2E-03 | 0.40 | 1353 | 0% | 70% | 0% | 6% | 24% |
| *LRP1* | Small Vessel Stroke (SVS) | rs11172113 | C | T | -11.53 | 9.0E-31 | 0.3 | 7.3E-01 | rs142888721 | A | G | 1.00 | 3.2E-01 | 3.7 | 2.2E-04 | 0.03 | 993 | 0% | 88% | 0% | 10% | 1% |
| *ATP2B1* | Small Vessel Stroke (SVS) | rs1689040 | T | C | -6.17 | 7.0E-10 | 0.0 | 9.6E-01 | rs1895711 | T | A | 3.98 | 7.2E-05 | 3.7 | 2.1E-04 | 0.10 | 1019 | 0% | 80% | 0% | 19% | 1% |
| *COL4A1* | Small Vessel Stroke (SVS) | rs7326444 | A | G | -6.46 | 1.0E-10 | 3.5 | 4.6E-04 | rs9521635 | C | T | -6.05 | 1.4E-09 | 4.1 | 3.6E-05 | 0.92 | 1142 | 0% | 12% | 0% | 6% | 82% |
| *COL4A2* | Small Vessel Stroke (SVS) | rs11838776 | A | G | -8.74 | 2.5E-18 | 4.6 | 4.9E-06 | rs55940034 | G | A | -8.56 | 1.2E-17 | 4.6 | 4.9E-06 | 0.94 | 1210 | 0% | 0% | 0% | 0% | 100% |
| *FBN1* | Small Vessel Stroke (SVS) | rs2437947 | A | T | 5.94 | 2.9E-09 | -0.2 | 8.6E-01 | rs2009404 | T | G | 0.49 | 6.2E-01 | -2.6 | 8.5E-03 | 0.00 | 522 | 0% | 96% | 0% | 2% | 2% |
| *THSD4* | Small Vessel Stroke (SVS) | rs10851839 | A | T | 6.56 | 5.5E-11 | 0.0 | 1.0E+00 | rs77103696 | A | G | 1.05 | 2.9E-01 | 2.3 | 2.2E-02 | 0.00 | 1131 | 0% | 96% | 0% | 3% | 1% |
| *LINC00310/KCNE2* | Small Vessel Stroke (SVS) | rs9980618 | T | C | -8.35 | 7.2E-17 | -1.3 | 2.0E-01 | rs7282240 | T | C | -3.80 | 1.5E-04 | -3.4 | 8.0E-04 | 0.28 | 1516 | 0% | 90% | 0% | 6% | 4% |
| *SYN3* | Small Vessel Stroke (SVS) | rs137528 | A | G | -5.40 | 6.8E-08 | -0.5 | 6.0E-01 | rs5994625 | G | A | 0.39 | 7.0E-01 | -3.9 | 9.6E-05 | 0.02 | 1852 | 1% | 78% | 0% | 19% | 3% |
| *FGGY-DT* | Forced expiratory volume in 1 sec (FEV1) | rs34370185 | T | G | 7.09 | 1.4E-12 | 0.0 | 9.8E-01 | rs114186484 | T | C | -1.01 | 3.1E-01 | -2.7 | 6.4E-03 | 0.01 | 1045 | 0% | 99% | 0% | 0% | 0% |
| *F3* | Forced expiratory volume in 1 sec (FEV1) | rs1146473 | C | T | 5.82 | 5.8E-09 | -0.5 | 6.0E-01 | rs112292390 | C | G | 0.62 | 5.3E-01 | 3.2 | 1.6E-03 | 0.00 | 1576 | 0% | 98% | 0% | 2% | 0% |
| *FALEC/ADAMTSL4-AS2* | Forced expiratory volume in 1 sec (FEV1) | rs4970935 | T | C | -13.04 | 6.1E-39 | -3.8 | 1.5E-04 | rs10888385 | C | T | -0.98 | 3.2E-01 | -7.6 | 3.5E-14 | 0.04 | 1033 | 0% | 0% | 0% | 100% | 0% |
| *AFAP1* | Forced expiratory volume in 1 sec (FEV1) | rs6828005 | A | G | -6.67 | 2.6E-11 | 0.3 | 7.8E-01 | rs4478172 | C | A | 5.02 | 5.3E-07 | -5.8 | 6.0E-09 | 0.04 | 2214 | 0% | 0% | 0% | 97% | 3% |
| *ZNF827* | Forced expiratory volume in 1 sec (FEV1) | rs1507928 | C | T | 5.75 | 8.9E-09 | 2.7 | 6.2E-03 | rs2135963 | G | C | 1.62 | 1.0E-01 | 4.7 | 2.9E-06 | 0.03 | 1241 | 0% | 68% | 0% | 28% | 4% |
| *ITGA1* | Forced expiratory volume in 1 sec (FEV1) | rs73102285 | G | A | 5.72 | 1.1E-08 | -1.8 | 7.0E-02 | rs9791046 | A | T | -0.42 | 6.8E-01 | 5.6 | 2.1E-08 | 0.00 | 2096 | 0% | 1% | 0% | 99% | 0% |
| *PHACTR1* | Forced expiratory volume in 1 sec (FEV1) | rs6925904 | G | A | -7.50 | 6.3E-14 | 0.3 | 7.6E-01 | rs1931546 | C | T | 1.29 | 2.0E-01 | -2.6 | 1.0E-02 | 0.00 | 1613 | 0% | 99% | 0% | 1% | 0% |
| *HTRA1* | Forced expiratory volume in 1 sec (FEV1) | rs2736923 | A | G | 5.47 | 4.6E-08 | -0.9 | 3.9E-01 | rs12571363 | A | C | -3.60 | 3.2E-04 | -7.7 | 1.6E-14 | 0.00 | 1553 | 0% | 0% | 0% | 97% | 3% |
| *SESN3/FAM76B* | Forced expiratory volume in 1 sec (FEV1) | rs11021221 | A | T | 7.84 | 4.1E-15 | -2.9 | 3.9E-03 | rs8181535 | T | C | 4.26 | 2.0E-05 | -3.5 | 5.6E-04 | 0.47 | 1359 | 0% | 86% | 0% | 2% | 12% |
| *LRP1* | Forced expiratory volume in 1 sec (FEV1) | rs11172113 | C | T | -11.53 | 9.0E-31 | 0.2 | 8.1E-01 | rs146529565 | C | A | 0.35 | 7.3E-01 | 4.2 | 2.3E-05 | 0.03 | 991 | 0% | 60% | 0% | 40% | 0% |
| *ATP2B1* | Forced expiratory volume in 1 sec (FEV1) | rs1689040 | T | C | -6.17 | 7.0E-10 | 1.3 | 1.9E-01 | rs17456901 | C | T | -1.48 | 1.4E-01 | 4.4 | 1.3E-05 | 0.00 | 1017 | 0% | 76% | 0% | 24% | 0% |
| *COL4A1* | Forced expiratory volume in 1 sec (FEV1) | rs7326444 | A | G | -6.46 | 1.0E-10 | -0.5 | 6.3E-01 | rs9301442 | C | T | 1.86 | 6.2E-02 | -3.3 | 8.4E-04 | 0.00 | 1148 | 0% | 99% | 0% | 1% | 0% |
| *COL4A2* | Forced expiratory volume in 1 sec (FEV1) | rs11838776 | A | G | -8.74 | 2.5E-18 | 0.9 | 3.7E-01 | rs9515279 | T | A | 1.62 | 1.1E-01 | -3.9 | 8.3E-05 | 0.00 | 1223 | 0% | 93% | 0% | 7% | 0% |
| *FBN1* | Forced expiratory volume in 1 sec (FEV1) | rs2437947 | A | T | 5.94 | 2.9E-09 | -0.9 | 3.8E-01 | rs8040242 | C | A | -0.20 | 8.5E-01 | -2.9 | 3.6E-03 | 0.01 | 527 | 0% | 99% | 0% | 1% | 0% |
| *THSD4* | Forced expiratory volume in 1 sec (FEV1) | rs10851839 | A | T | 6.56 | 5.5E-11 | 6.1 | 1.4E-09 | rs8033889 | T | G | -5.31 | 1.1E-07 | -6.8 | 1.4E-11 | 0.30 | 1135 | 0% | 0% | 0% | 34% | 66% |
| *LINC00310/KCNE2* | Forced expiratory volume in 1 sec (FEV1) | rs9980618 | T | C | -8.35 | 7.2E-17 | -2.5 | 1.4E-02 | rs2236610 | G | C | 0.99 | 3.2E-01 | -4.4 | 1.3E-05 | 0.00 | 1518 | 0% | 66% | 0% | 31% | 3% |
| *SYN3* | Forced expiratory volume in 1 sec (FEV1) | rs137528 | A | G | -5.40 | 6.8E-08 | 2.6 | 1.0E-02 | rs5998705 | G | A | 2.81 | 5.0E-03 | -4.4 | 1.3E-05 | 0.20 | 1863 | 1% | 84% | 0% | 10% | 5% |
| *FGGY-DT* | HDL | rs34370185 | T | G | 7.09 | 1.4E-12 | -0.4 | 6.7E-01 | rs2492323 | T | G | 0.05 | 9.6E-01 | 3.0 | 2.3E-03 | 0.00 | 1045 | 0% | 99% | 0% | 1% | 0% |
| *F3* | HDL | rs1146473 | C | T | 5.82 | 5.8E-09 | -0.1 | 9.1E-01 | rs4147820 | T | C | -1.11 | 2.7E-01 | -4.4 | 1.1E-05 | 0.00 | 1576 | 0% | 94% | 0% | 6% | 0% |
| *FALEC/ADAMTSL4-AS2* | HDL | rs4970935 | T | C | -13.04 | 6.1E-39 | 2.9 | 3.8E-03 | rs41271951 | G | A | -1.31 | 1.9E-01 | 7.7 | 1.2E-14 | 0.01 | 1033 | 0% | 0% | 0% | 100% | 0% |

| | | | | | | | | | | | | | | | | | | | | | | |
|---|---|---|---|---|---|---|---|---|---|---|---|---|---|---|---|---|---|---|---|---|---|---|
| *AFAP1* | HDL | rs6828005 | A | G | -6.67 | 2.6E-11 | 1.1 | 2.6E-01 | rs2002574 | C | T | 0.20 | 8.5E-01 | -4.0 | 5.7E-05 | 0.00 | 2214 | 0% | 97% | 0% | 2% | 0% |
| *ZNF827* | HDL | rs1507928 | C | T | 5.75 | 8.9E-09 | 1.3 | 1.8E-01 | rs13134992 | T | C | 0.40 | 6.9E-01 | 5.6 | 1.8E-08 | 0.01 | 1241 | 0% | 2% | 0% | 98% | 0% |
| *ITGA1* | HDL | rs73102285 | G | A | 5.72 | 1.1E-08 | 0.8 | 4.2E-01 | rs61448800 | A | G | 0.98 | 3.3E-01 | -3.6 | 2.8E-04 | 0.00 | 2096 | 0% | 95% | 0% | 4% | 0% |
| *PHACTR1* | HDL | rs6925904 | G | A | -7.50 | 6.3E-14 | 1.4 | 1.5E-01 | rs4714946 | A | G | 3.74 | 1.8E-04 | -2.9 | 3.6E-03 | 0.41 | 1613 | 0% | 98% | 0% | 1% | 0% |
| *HTRA1* | HDL | rs2736923 | A | G | 5.47 | 4.6E-08 | -1.7 | 9.7E-02 | rs9423289 | T | C | 0.09 | 9.3E-01 | 4.0 | 7.6E-05 | 0.00 | 1553 | 0% | 96% | 0% | 3% | 1% |
| *SESN3/FAM76B* | HDL | rs11021221 | A | T | 7.84 | 4.1E-15 | 0.6 | 5.4E-01 | rs11021232 | C | T | -1.53 | 1.3E-01 | -6.0 | 1.7E-09 | 0.05 | 1359 | 0% | 0% | 0% | 100% | 0% |
| *LRP1* | HDL | rs11172113 | C | T | -11.53 | 9.0E-31 | 3.8 | 1.3E-04 | rs61352607 | T | G | -1.66 | 9.7E-02 | 12.3 | 6.9E-35 | 0.02 | 991 | 0% | 0% | 0% | 100% | 0% |
| *ATP2B1* | HDL | rs1689040 | T | C | -6.17 | 7.0E-10 | 0.5 | 6.3E-01 | rs12371151 | T | A | 2.02 | 4.3E-02 | -3.3 | 8.5E-04 | 0.00 | 1017 | 0% | 99% | 0% | 1% | 0% |
| *COL4A1* | HDL | rs7326444 | A | G | -6.46 | 1.0E-10 | -0.5 | 6.3E-01 | rs11841319 | T | C | -1.30 | 1.9E-01 | -3.5 | 5.0E-04 | 0.00 | 1148 | 0% | 99% | 0% | 1% | 0% |
| *COL4A2* | HDL | rs11838776 | A | G | -8.74 | 2.5E-18 | 0.9 | 3.5E-01 | rs9521977 | C | T | -0.66 | 5.1E-01 | -4.3 | 1.6E-05 | 0.00 | 1223 | 0% | 91% | 0% | 9% | 0% |
| *FBN1* | HDL | rs2437947 | A | T | 5.94 | 2.9E-09 | -2.3 | 2.4E-02 | rs117540572 | A | G | 0.92 | 3.6E-01 | 3.0 | 2.9E-03 | 0.00 | 527 | 0% | 96% | 0% | 1% | 3% |
| *THSD4* | HDL | rs10851839 | A | T | 6.56 | 5.5E-11 | 2.2 | 3.0E-02 | rs75125772 | C | G | -0.64 | 5.2E-01 | 2.8 | 5.0E-03 | 0.00 | 1135 | 0% | 98% | 0% | 1% | 2% |
| *LINC00310/KCNE2* | HDL | rs9980618 | T | C | -8.35 | 7.2E-17 | 1.6 | 1.0E-01 | rs9978142 | T | A | -5.53 | 3.3E-08 | 2.8 | 5.7E-03 | 0.67 | 1518 | 0% | 98% | 0% | 1% | 1% |
| *SYN3* | HDL | rs137528 | A | G | -5.40 | 6.8E-08 | -0.4 | 6.6E-01 | rs58930195 | C | T | 0.31 | 7.6E-01 | 2.2 | 2.5E-02 | 0.00 | 1863 | 1% | 98% | 0% | 1% | 0% |
| *FGGY-DT* | LDL | rs34370185 | T | G | 7.09 | 1.4E-12 | 2.6 | 8.7E-03 | rs6678642 | G | T | 6.12 | 9.7E-10 | 3.0 | 2.4E-03 | 0.70 | 1045 | 0% | 93% | 0% | 2% | 6% |
| *F3* | LDL | rs1146473 | C | T | 5.82 | 5.8E-09 | 2.0 | 4.9E-02 | rs11807145 | T | C | 0.73 | 4.6E-01 | -3.6 | 3.4E-04 | 0.04 | 1576 | 0% | 97% | 0% | 2% | 1% |
| *FALEC/ADAMTSL4-AS2* | LDL | rs4970935 | T | C | -13.04 | 6.1E-39 | -2.2 | 2.9E-02 | rs267733 | G | A | -2.99 | 2.8E-03 | -5.2 | 2.2E-07 | 0.02 | 1033 | 0% | 20% | 0% | 79% | 0% |
| *AFAP1* | LDL | rs6828005 | A | G | -6.67 | 2.6E-11 | -0.8 | 4.4E-01 | rs2002574 | C | T | 0.20 | 8.5E-01 | -4.6 | 3.8E-06 | 0.00 | 2214 | 0% | 91% | 0% | 8% | 1% |
| *ZNF827* | LDL | rs1507928 | C | T | 5.75 | 8.9E-09 | 0.1 | 8.9E-01 | rs6819403 | A | C | -0.44 | 6.6E-01 | 3.0 | 2.6E-03 | 0.00 | 1241 | 0% | 99% | 0% | 1% | 0% |
| *ITGA1* | LDL | rs73102285 | G | A | 5.72 | 1.1E-08 | -0.6 | 5.4E-01 | rs116734477 | T | C | 1.50 | 1.3E-01 | -8.7 | 4.5E-18 | 0.00 | 2096 | 0% | 0% | 0% | 100% | 0% |
| *PHACTR1* | LDL | rs6925904 | G | A | -7.50 | 6.3E-14 | -1.4 | 1.7E-01 | rs36102538 | G | A | 0.61 | 5.4E-01 | 2.6 | 8.3E-03 | 0.01 | 1613 | 0% | 99% | 0% | 1% | 0% |
| *HTRA1* | LDL | rs2736923 | A | G | 5.47 | 4.6E-08 | -2.0 | 4.6E-02 | rs9423289 | T | C | 0.09 | 9.3E-01 | 6.9 | 6.0E-12 | 0.00 | 1553 | 0% | 0% | 0% | 100% | 0% |
| *SESN3/FAM76B* | LDL | rs11021221 | A | T | 7.84 | 4.1E-15 | 2.9 | 4.0E-03 | rs3858379 | C | T | -0.44 | 6.6E-01 | -4.3 | 1.6E-05 | 0.00 | 1359 | 0% | 82% | 0% | 7% | 11% |
| *LRP1* | LDL | rs11172113 | C | T | -11.53 | 9.0E-31 | 0.8 | 4.0E-01 | rs2122982 | A | G | -1.85 | 6.5E-02 | -6.0 | 1.5E-09 | 0.02 | 991 | 0% | 0% | 0% | 100% | 0% |
| *ATP2B1* | LDL | rs1689040 | T | C | -6.17 | 7.0E-10 | 2.3 | 2.3E-02 | rs12306780 | T | A | 1.05 | 2.9E-01 | 4.5 | 5.6E-06 | 0.00 | 1017 | 0% | 44% | 0% | 55% | 1% |
| *COL4A1* | LDL | rs7326444 | A | G | -6.46 | 1.0E-10 | -1.0 | 3.4E-01 | rs532625 | T | A | 0.43 | 6.7E-01 | -3.9 | 8.0E-05 | 0.01 | 1148 | 0% | 98% | 0% | 2% | 0% |
| *COL4A2* | LDL | rs11838776 | A | G | -8.74 | 2.5E-18 | -5.3 | 1.2E-07 | rs55940034 | G | A | -8.56 | 1.2E-17 | -5.4 | 6.9E-08 | 0.94 | 1223 | 0% | 0% | 0% | 1% | 99% |
| *FBN1* | LDL | rs2437947 | A | T | 5.94 | 2.9E-09 | 1.7 | 9.3E-02 | rs363820 | C | T | 5.16 | 2.5E-07 | 1.9 | 6.0E-02 | 0.90 | 527 | 0% | 99% | 0% | 0% | 1% |
| *THSD4* | LDL | rs10851839 | A | T | 6.56 | 5.5E-11 | -1.5 | 1.3E-01 | rs13379731 | C | G | -1.47 | 1.4E-01 | 3.4 | 5.9E-04 | 0.09 | 1135 | 0% | 98% | 0% | 1% | 0% |
| *LINC00310/KCNE2* | LDL | rs9980618 | T | C | -8.35 | 7.2E-17 | -1.3 | 1.8E-01 | rs34323310 | T | C | 0.73 | 4.7E-01 | -4.0 | 6.5E-05 | 0.00 | 1518 | 0% | 93% | 0% | 6% | 0% |
| *SYN3* | LDL | rs137528 | A | G | -5.40 | 6.8E-08 | -0.8 | 4.0E-01 | rs11703454 | A | G | -2.36 | 1.8E-02 | -2.7 | 7.2E-03 | 0.00 | 1863 | 1% | 98% | 0% | 1% | 0% |
| *FGGY-DT* | Migraine | rs34370185 | T | G | 7.09 | 1.4E-12 | 0.2 | 8.2E-01 | rs932770 | A | G | 1.81 | 7.1E-02 | 3.2 | 1.6E-03 | 0.01 | 1045 | 0% | 100% | 0% | 0% | 0% |
| *F3* | Migraine | rs1146473 | C | T | 5.82 | 5.8E-09 | -0.3 | 7.9E-01 | rs560426 | T | C | 0.76 | 4.5E-01 | -2.8 | 4.9E-03 | 0.01 | 1576 | 0% | 100% | 0% | 0% | 0% |
| *FALEC/ADAMTSL4-AS2* | Migraine | rs4970935 | T | C | -13.04 | 6.1E-39 | -4.6 | 5.0E-06 | rs6693567 | T | C | -11.13 | 9.8E-29 | -5.2 | 1.9E-07 | 0.90 | 1033 | 0% | 11% | 0% | 2% | 86% |
| *AFAP1* | Migraine | rs6828005 | A | G | -6.67 | 2.6E-11 | -2.1 | 3.3E-02 | rs4696794 | G | A | -2.92 | 3.5E-03 | -3.7 | 2.1E-04 | 0.17 | 2214 | 0% | 99% | 0% | 0% | 0% |
| *ZNF827* | Migraine | rs1507928 | C | T | 5.75 | 8.9E-09 | -0.1 | 9.0E-01 | rs10029927 | T | C | 0.33 | 7.4E-01 | 2.8 | 5.3E-03 | 0.01 | 1241 | 0% | 100% | 0% | 0% | 0% |
| *ITGA1* | Migraine | rs73102285 | G | A | 5.72 | 1.1E-08 | 0.0 | 9.9E-01 | rs78464352 | A | G | -1.55 | 1.2E-01 | -3.2 | 1.3E-03 | 0.00 | 2096 | 0% | 100% | 0% | 0% | 0% |
| *PHACTR1* | Migraine | rs6925904 | G | A | -7.50 | 6.3E-14 | -6.4 | 1.2E-10 | rs1332844 | T | C | -7.38 | 1.7E-13 | -6.5 | 8.1E-11 | 1.00 | 1613 | 0% | 0% | 0% | 2% | 98% |
| *HTRA1* | Migraine | rs2736923 | A | G | 5.47 | 4.6E-08 | 2.9 | 3.2E-03 | rs139101261 | C | G | -3.15 | 1.6E-03 | -3.5 | 4.0E-04 | 0.01 | 1553 | 0% | 97% | 0% | 1% | 2% |
| *SESN3/FAM76B* | Migraine | rs11021221 | A | T | 7.84 | 4.1E-15 | 2.1 | 3.3E-02 | rs148907559 | T | A | 0.03 | 9.8E-01 | 3.0 | 2.5E-03 | 0.01 | 1359 | 0% | 100% | 0% | 0% | 0% |
| *LRP1* | Migraine | rs11172113 | C | T | -11.53 | 9.0E-31 | -10.1 | 4.0E-24 | rs11172113 | C | T | -11.53 | 9.0E-31 | -10.1 | 4.0E-24 | 1.00 | 991 | 0% | 0% | 0% | 0% | 100% |
| *ATP2B1* | Migraine | rs1689040 | T | C | -6.17 | 7.0E-10 | -2.8 | 5.7E-03 | rs6538222 | A | G | 0.90 | 3.7E-01 | 3.0 | 3.1E-03 | 0.00 | 1017 | 0% | 99% | 0% | 0% | 1% |
| *COL4A1* | Migraine | rs7326444 | A | G | -6.46 | 1.0E-10 | -1.9 | 5.3E-02 | rs7995770 | G | A | -0.33 | 7.4E-01 | -4.0 | 6.4E-05 | 0.00 | 1148 | 0% | 99% | 0% | 0% | 0% |
| *COL4A2* | Migraine | rs11838776 | A | G | -8.74 | 2.5E-18 | -2.5 | 1.4E-02 | rs9521762 | A | G | 0.96 | 3.4E-01 | -3.7 | 2.6E-04 | 0.05 | 1223 | 0% | 99% | 0% | 0% | 0% |
| *FBN1* | Migraine | rs2437947 | A | T | 5.94 | 2.9E-09 | -0.2 | 8.1E-01 | rs28730795 | G | T | 1.69 | 9.1E-02 | 2.2 | 2.7E-02 | 0.24 | 527 | 0% | 100% | 0% | 0% | 0% |
| *THSD4* | Migraine | rs10851839 | A | T | 6.56 | 5.5E-11 | 0.2 | 8.1E-01 | rs138365904 | G | A | -1.95 | 5.1E-02 | 3.3 | 9.5E-04 | 0.11 | 1135 | 0% | 100% | 0% | 0% | 0% |
| *LINC00310/KCNE2* | Migraine | rs9980618 | T | C | -8.35 | 7.2E-17 | -3.9 | 8.5E-05 | rs8131284 | C | T | -7.61 | 2.8E-14 | -4.0 | 6.1E-05 | 0.80 | 1518 | 0% | 57% | 0% | 1% | 42% |
| *SYN3* | Migraine | rs137528 | A | G | -5.40 | 6.8E-08 | -1.0 | 2.9E-01 | rs8138938 | A | G | -0.03 | 9.8E-01 | -3.3 | 9.6E-04 | 0.02 | 1863 | 1% | 99% | 0% | 0% | 0% |
| *FGGY-DT* | Hemoglobin (HGB) | rs34370185 | T | G | 7.09 | 1.4E-12 | 5.1 | 4.3E-07 | rs34370185 | T | G | 7.09 | 1.4E-12 | 5.1 | 4.3E-07 | 1.00 | 1046 | 0% | 0% | 0% | 2% | 97% |
| *F3* | Hemoglobin (HGB) | rs1146473 | C | T | 5.82 | 5.8E-09 | 0.8 | 4.5E-01 | rs871662 | G | A | -1.30 | 1.9E-01 | -3.8 | 1.5E-04 | 0.04 | 1576 | 0% | 94% | 0% | 5% | 1% |
| *FALEC/ADAMTSL4-AS2* | Hemoglobin (HGB) | rs4970935 | T | C | -13.04 | 6.1E-39 | -0.3 | 7.8E-01 | rs72700806 | G | A | -1.81 | 7.0E-02 | 3.5 | 3.9E-04 | 0.06 | 1037 | 0% | 88% | 0% | 12% | 0% |
| *AFAP1* | Hemoglobin (HGB) | rs6828005 | A | G | -6.67 | 2.6E-11 | 3.1 | 2.2E-03 | rs12501350 | T | C | 2.21 | 2.7E-02 | -3.6 | 3.5E-04 | 0.47 | 2215 | 0% | 93% | 0% | 2% | 5% |
| *ZNF827* | Hemoglobin (HGB) | rs1507928 | C | T | 5.75 | 8.9E-09 | -0.6 | 5.2E-01 | rs714195 | C | T | -0.10 | 9.2E-01 | 6.3 | 2.8E-10 | 0.01 | 1242 | 0% | 0% | 0% | 100% | 0% |
| *ITGA1* | Hemoglobin (HGB) | rs73102285 | G | A | 5.72 | 1.1E-08 | -0.5 | 6.5E-01 | rs60925406 | C | T | -1.10 | 2.7E-01 | -6.6 | 3.0E-11 | 0.00 | 2096 | 0% | 0% | 0% | 100% | 0% |
| *PHACTR1* | Hemoglobin (HGB) | rs6925904 | G | A | -7.50 | 6.3E-14 | -0.5 | 6.3E-01 | rs62386724 | C | T | 0.09 | 9.3E-01 | 3.4 | 7.1E-04 | 0.00 | 1613 | 0% | 97% | 0% | 3% | 0% |
| *HTRA1* | Hemoglobin (HGB) | rs2736923 | A | G | 5.47 | 4.6E-08 | 0.6 | 5.4E-01 | rs4980169 | G | A | 0.00 | 1.0E+00 | -4.4 | 9.2E-06 | 0.00 | 1556 | 0% | 71% | 0% | 29% | 0% |
| *SESN3/FAM76B* | Hemoglobin (HGB) | rs11021221 | A | T | 7.84 | 4.1E-15 | -2.7 | 6.3E-03 | rs12800440 | T | C | -0.34 | 7.3E-01 | -6.2 | 5.7E-10 | 0.00 | 1360 | 0% | 0% | 0% | 100% | 0% |
| *LRP1* | Hemoglobin (HGB) | rs11172113 | C | T | -11.53 | 9.0E-31 | -3.2 | 1.2E-03 | rs3741414 | T | C | -0.64 | 5.2E-01 | 9.1 | 8.4E-20 | 0.02 | 999 | 0% | 0% | 0% | 100% | 0% |
| *ATP2B1* | Hemoglobin (HGB) | rs1689040 | T | C | -6.17 | 7.0E-10 | -2.0 | 4.6E-02 | rs2615834 | A | T | -1.28 | 2.0E-01 | -4.9 | 1.0E-06 | 0.00 | 1024 | 0% | 30% | 0% | 70% | 0% |
| *COL4A1* | Hemoglobin (HGB) | rs7326444 | A | G | -6.46 | 1.0E-10 | 0.8 | 4.4E-01 | rs79692857 | A | C | -0.33 | 7.4E-01 | 6.3 | 2.9E-10 | 0.00 | 1148 | 0% | 0% | 0% | 100% | 0% |
| *COL4A2* | Hemoglobin (HGB) | rs11838776 | A | G | -8.74 | 2.5E-18 | -3.7 | 2.2E-04 | rs4103 | T | C | -0.36 | 7.2E-01 | 5.2 | 1.9E-07 | 0.06 | 1225 | 0% | 16% | 0% | 64% | 20% |

| | | | | | | | | | | | | | | | | | | | | | | |
|---|---|---|---|---|---|---|---|---|---|---|---|---|---|---|---|---|---|---|---|---|---|---|
| *FBN1* | Hemoglobin (HGB) | rs2437947 | A | T | 5.94 | 2.9E-09 | -0.6 | 5.7E-01 | rs17462641 | T | C | -1.89 | 5.8E-02 | 4.9 | 1.1E-06 | 0.02 | 527 | 0% | 6% | 0% | 94% | 0% |
| *THSD4* | Hemoglobin (HGB) | rs10851839 | A | T | 6.56 | 5.5E-11 | -4.9 | 9.2E-07 | rs2625529 | C | G | 0.36 | 7.2E-01 | -7.8 | 8.9E-15 | 0.00 | 1139 | 0% | 0% | 0% | 100% | 0% |
| *LINC00310/KCNE2* | Hemoglobin (HGB) | rs9980618 | T | C | -8.35 | 7.2E-17 | 0.6 | 5.7E-01 | rs2834317 | A | G | 2.33 | 2.0E-02 | -12.5 | 8.0E-36 | 0.00 | 1519 | 0% | 0% | 0% | 100% | 0% |
| *SYN3* | Hemoglobin (HGB) | rs137528 | A | G | -5.40 | 6.8E-08 | 0.9 | 3.6E-01 | rs9606967 | C | G | 1.22 | 2.2E-01 | -4.7 | 3.0E-06 | 0.00 | 1864 | 1% | 75% | 0% | 24% | 0% |

**Supplementary Table 8: Genetic correlation between SCAD and cardiovascular and neurovascular diseases and traits**

rg: genetic correlation, se: standard error, P: P-value for the genetic correlation.

CAD: coronary artery disease, MI: myocardial infraction, FMD: fibromuscular dysplasia, CeAD: cervical artery, dissection, IA: intracranial aneurysm, SAH: subarachnoid hemorrhage , uIA: unreptured intracranial aneurysm ,AS: any stoke, AIS: any ischemic stroke, LAS: , CES: cardioembolic stroke, SVS: small vessel stroke, MIG: Migraine, T2D: type 2 diabete, SMK: smoking (never/ever), SBP: systolic blood pressure, DBP: diastolic blood pressure, PP: pulse pressure, HTN: Hypertension, BMI: body mass index , HDL: high density lipoprotein, LDL: low density lipoprotein, TC: total cholesterol, TG: triglycerides, HGB: haemoglobin, HCT: haematocrit, NEU : Neutrophil count, MONO : Monocyte count, LYMPHO : Lymphocyte count and PLT: Platelet count

details on used GWAS summary statistics of disases and traits are available in Supplementary table 15

| | Unadjusted | | | | | | SBP mtCOJO | | | | | | DBP mtCOJO | | | | | |
|---|---|---|---|---|---|---|---|---|---|---|---|---|---|---|---|---|---|---|
| **Disease or trait** | **rg** | **se** | **L95_rg** | **U95_rg** | **P** | **P.adj** | **rg** | **se** | **L95_rg** | **U95_rg** | **P** | **P.adj** | **rg** | **se** | **L95_rg** | **U95_rg** | **P** | **P.adj** |
| **CAD** | -0.12 | 0.04 | -0.21 | -0.04 | 3.7E-03 | 1.0E-01 | -0.19 | 0.04 | -0.28 | -0.11 | 4.6E-06 | 1.2E-04 | -0.19 | 0.04 | -0.27 | -0.10 | 1.3E-05 | 3.4E-04 |
| **MI** | -0.14 | 0.05 | -0.24 | -0.04 | 6.6E-03 | 1.8E-01 | -0.21 | 0.05 | -0.31 | -0.10 | 1.5E-04 | 3.9E-03 | -0.21 | 0.06 | -0.32 | -0.10 | 1.6E-04 | 4.2E-03 |
| **FMD** | 0.38 | 0.18 | 0.03 | 0.74 | 3.5E-02 | 9.8E-01 | 0.30 | 0.15 | 0.00 | 0.60 | 5.3E-02 | 1.0E+00 | 0.30 | 0.15 | 0.00 | 0.60 | 5.2E-02 | 1.0E+00 |
| **CeAD** | 0.61 | 0.20 | 0.21 | 1.00 | 2.4E-03 | 6.7E-02 | 0.52 | 0.20 | 0.13 | 0.92 | 9.3E-03 | 2.4E-01 | 0.51 | 0.20 | 0.11 | 0.90 | 1.2E-02 | 3.1E-01 |
| **IA** | 0.22 | 0.06 | 0.10 | 0.33 | 2.0E-04 | 5.6E-03 | 0.16 | 0.06 | 0.04 | 0.27 | 6.9E-03 | 1.8E-01 | 0.16 | 0.06 | 0.04 | 0.27 | 8.2E-03 | 2.1E-01 |
| **SAH** | 0.27 | 0.07 | 0.14 | 0.40 | 6.4E-05 | 1.8E-03 | 0.20 | 0.07 | 0.07 | 0.33 | 2.8E-03 | 7.3E-02 | 0.20 | 0.07 | 0.07 | 0.33 | 3.3E-03 | 8.6E-02 |
| **uIA** | 0.14 | 0.08 | -0.02 | 0.30 | 9.0E-02 | 1.0E+00 | 0.09 | 0.08 | -0.07 | 0.26 | 2.6E-01 | 1.0E+00 | 0.09 | 0.08 | -0.08 | 0.26 | 2.9E-01 | 1.0E+00 |
| **AS** | 0.17 | 0.06 | 0.05 | 0.29 | 4.5E-03 | 1.3E-01 | 0.10 | 0.06 | -0.02 | 0.22 | 1.1E-01 | 1.0E+00 | 0.10 | 0.06 | -0.02 | 0.23 | 1.1E-01 | 1.0E+00 |
| **AIS** | 0.17 | 0.06 | 0.05 | 0.28 | 4.6E-03 | 1.3E-01 | 0.09 | 0.06 | -0.03 | 0.22 | 1.3E-01 | 1.0E+00 | 0.10 | 0.06 | -0.03 | 0.22 | 1.3E-01 | 1.0E+00 |
| **LAS** | 0.68 | 0.55 | -0.39 | 1.76 | 2.1E-01 | 1.0E+00 | 0.46 | 0.28 | -0.08 | 1.01 | 9.4E-02 | 1.0E+00 | 0.51 | 0.30 | -0.07 | 1.10 | 8.4E-02 | 1.0E+00 |
| **CES** | 0.34 | 0.09 | 0.17 | 0.51 | 1.0E-04 | 2.8E-03 | 0.34 | 0.09 | 0.16 | 0.52 | 2.0E-04 | 5.2E-03 | 0.34 | 0.09 | 0.16 | 0.53 | 2.0E-04 | 5.2E-03 |
| **SVS** | -0.04 | 0.10 | -0.24 | 0.16 | 7.2E-01 | 1.0E+00 | -0.09 | 0.11 | -0.31 | 0.13 | 4.1E-01 | 1.0E+00 | -0.11 | 0.11 | -0.32 | 0.11 | 3.2E-01 | 1.0E+00 |
| **MIG** | 0.18 | 0.06 | 0.07 | 0.30 | 1.3E-03 | 3.6E-02 | 0.18 | 0.05 | 0.08 | 0.29 | 5.0E-04 | 1.3E-02 | 0.16 | 0.05 | 0.06 | 0.27 | 2.5E-03 | 6.5E-02 |
| **T2D** | 0.02 | 0.08 | -0.15 | 0.18 | 8.5E-01 | 1.0E+00 | -0.03 | 0.07 | -0.17 | 0.12 | 7.3E-01 | 1.0E+00 | 0.00 | 0.07 | -0.15 | 0.14 | 9.5E-01 | 1.0E+00 |
| **SMK** | 0.00 | 0.03 | -0.06 | 0.06 | 9.0E-01 | 1.0E+00 | 0.02 | 0.03 | -0.04 | 0.08 | 5.0E-01 | 1.0E+00 | 0.02 | 0.03 | -0.04 | 0.07 | 5.5E-01 | 1.0E+00 |
| **SBP** | 0.12 | 0.03 | 0.06 | 0.19 | 1.0E-04 | 2.8E-03 | - | - | - | - | - | - | - | - | - | - | - | - |
| **DBP** | 0.17 | 0.03 | 0.11 | 0.24 | 2.6E-07 | 7.3E-06 | - | - | - | - | - | - | - | - | - | - | - | - |
| **BMI** | -0.02 | 0.03 | -0.08 | 0.03 | 4.1E-01 | 1.0E+00 | -0.02 | 0.03 | -0.07 | 0.03 | 4.9E-01 | 1.0E+00 | -0.02 | 0.03 | -0.07 | 0.04 | 5.3E-01 | 1.0E+00 |
| **HDL** | -0.09 | 0.03 | -0.15 | -0.03 | 1.8E-03 | 5.0E-02 | -0.08 | 0.03 | -0.14 | -0.03 | 2.1E-03 | 5.5E-02 | -0.08 | 0.03 | -0.13 | -0.03 | 3.6E-03 | 9.4E-02 |
| **LDL** | -0.07 | 0.03 | -0.14 | 0.00 | 4.5E-02 | 1.0E+00 | -0.04 | 0.03 | -0.09 | 0.02 | 2.0E-01 | 1.0E+00 | -0.04 | 0.03 | -0.10 | 0.02 | 1.6E-01 | 1.0E+00 |
| **TC** | -0.08 | 0.03 | -0.14 | -0.02 | 1.0E-02 | 2.8E-01 | -0.06 | 0.03 | -0.11 | 0.00 | 3.5E-02 | 9.1E-01 | -0.06 | 0.03 | -0.11 | -0.01 | 3.1E-02 | 8.1E-01 |
| **TG** | 0.06 | 0.03 | -0.01 | 0.12 | 8.6E-02 | 1.0E+00 | 0.04 | 0.03 | -0.02 | 0.09 | 2.0E-01 | 1.0E+00 | 0.03 | 0.03 | -0.02 | 0.09 | 2.3E-01 | 1.0E+00 |
| **HGB** | 0.12 | 0.03 | 0.06 | 0.17 | 2.7E-05 | 7.6E-04 | 0.08 | 0.03 | 0.03 | 0.14 | 1.2E-03 | 3.1E-02 | 0.05 | 0.03 | 0.00 | 0.10 | 3.9E-02 | 1.0E+00 |
| **HCT** | 0.12 | 0.03 | 0.06 | 0.17 | 4.9E-05 | 1.4E-03 | 0.09 | 0.03 | 0.04 | 0.14 | 1.0E-03 | 2.6E-02 | 0.06 | 0.03 | 0.01 | 0.11 | 2.9E-02 | 7.5E-01 |
| **NEU** | 0.02 | 0.03 | -0.04 | 0.08 | 6.1E-01 | 1.0E+00 | -0.01 | 0.03 | -0.07 | 0.04 | 6.4E-01 | 1.0E+00 | -0.01 | 0.03 | -0.06 | 0.05 | 8.2E-01 | 1.0E+00 |
| **MONO** | 0.00 | 0.03 | -0.06 | 0.07 | 9.0E-01 | 1.0E+00 | -0.01 | 0.03 | -0.06 | 0.04 | 7.4E-01 | 1.0E+00 | -0.01 | 0.03 | -0.06 | 0.04 | 7.4E-01 | 1.0E+00 |
| **LYMPHO** | -0.05 | 0.04 | -0.13 | 0.02 | 1.6E-01 | 1.0E+00 | -0.08 | 0.03 | -0.14 | -0.02 | 5.3E-03 | 1.4E-01 | -0.08 | 0.03 | -0.14 | -0.02 | 5.6E-03 | 1.5E-01 |
| **PLT** | -0.06 | 0.03 | -0.11 | -0.01 | 1.4E-02 | 3.9E-01 | -0.06 | 0.02 | -0.10 | -0.01 | 1.6E-02 | 4.2E-01 | -0.05 | 0.02 | -0.10 | -0.01 | 2.3E-02 | 6.0E-01 |

**Supplementary Table 9: Comparaison of SCAD meta-analysis results, unstratified and stratified on FMD status**

EA: Effect Allele, OA: Other Allele, OR: Odds Ratio, CI: Confidence Interval ,EAF: Effect Allele Frequency, P: P-value of genetic association, Het_P: P-value of heterogeneity

| CHR | POS | LOCUS | TOP SNP SCAD | EA | OA | Unstratified [1268-1917] Cases / [6896-9291] Controls | | | | SCAD with FMD [203-409] Cases / [3036-4961] Controls | | | | SCAD without FMD [305-614] Cases / [3400-5776] Controls | | | |
|---|---|---|---|---|---|---|---|---|---|---|---|---|---|---|---|---|---|
| | | | | | | OR (95% CI) | P | EAF | Het_P | OR (95% CI) | P | EAF | Het_P | OR (95% CI) | P | EAF | Het_P |
| 1 | 59656909 | *FGGY-DT* | rs34370185 | T | G | 1.34 (1.24 - 1.46) | 1.4E-12 | 0.29 | 0.044 | 1.23 (1.05 - 1.46) | 1.2E-02 | 0.28 | 0.396 | 1.35 (1.18 - 1.55) | 1.5E-05 | 0.29 | 0.076 |
| 1 | 95050472 | *F3* | rs1146473 | C | T | 1.32 (1.2 - 1.45) | 5.8E-09 | 0.19 | 0.098 | 1.31 (1.09 - 1.56) | 3.5E-03 | 0.19 | 0.256 | 1.32 (1.13 - 1.55) | 4.2E-04 | 0.19 | 0.001 |
| 1 | 150504062 | *FALEC/ ADAMTSL4-AS2* | rs4970935 | C | T | 1.72 (1.59 - 1.87) | 6.1E-39 | 0.28 | 0.636 | 1.77 (1.51 - 2.08) | 3.7E-12 | 0.27 | 0.655 | 1.78 (1.55 - 2.04) | 9.0E-17 | 0.27 | 0.808 |
| 4 | 7774352 | *AFAP1* | rs6828005 | G | A | 1.29 (1.2 - 1.4) | 2.6E-11 | 0.45 | 0.816 | 1.41 (1.21 - 1.65) | 9.6E-06 | 0.44 | 0.193 | 1.25 (1.1 - 1.42) | 4.9E-04 | 0.44 | 0.009 |
| 4 | 146788035 | *ZNF827* | rs1507928 | C | T | 1.25 (1.16 - 1.35) | 8.9E-09 | 0.48 | 0.385 | 1.34 (1.15 - 1.56) | 1.8E-04 | 0.47 | 0.920 | 1.25 (1.1 - 1.42) | 6.1E-04 | 0.47 | 0.305 |
| 5 | 52155642 | *ITGA1* | rs73102285 | G | A | 1.27 (1.17 - 1.38) | 1.1E-08 | 0.27 | 0.312 | 1.3 (1.1 - 1.54) | 1.8E-03 | 0.27 | 0.598 | 1.36 (1.18 - 1.56) | 1.7E-05 | 0.27 | 0.377 |
| 6 | 12903957 | *PHACTR1* | rs9349379 | A | G | 1.64 (1.51 - 1.78) | 2.9E-32 | 0.62 | 0.193 | 1.67 (1.42 - 1.97) | 1.3E-09 | 0.61 | 0.244 | 1.63 (1.42 - 1.87) | 6.3E-12 | 0.61 | 0.184 |
| 10 | 124259062 | *HTRA1* | rs2736923 | A | G | 1.44 (1.26 - 1.64) | 4.6E-08 | 0.89 | 0.600 | 1.53 (1.17 - 2.01) | 2.1E-03 | 0.88 | 0.488 | 1.4 (1.13 - 1.73) | 2.2E-03 | 0.88 | 0.424 |
| 11 | 95308854 | *SESN3/FAM76B* | rs11021221 | A | T | 1.47 (1.33 - 1.61) | 4.1E-15 | 0.17 | 0.192 | 1.62 (1.35 - 1.95) | 3.1E-07 | 0.17 | 0.462 | 1.3 (1.1 - 1.52) | 1.9E-03 | 0.17 | 0.320 |
| 12 | 57527283 | *LRP1* | rs11172113 | T | C | 1.62 (1.49 - 1.76) | 9.0E-31 | 0.62 | 0.702 | 1.61 (1.36 - 1.9) | 1.6E-08 | 0.61 | 0.477 | 1.6 (1.39 - 1.83) | 2.6E-11 | 0.61 | 0.280 |
| 12 | 89978233 | *ATP2B1* | rs1689040 | C | T | 1.28 (1.18 - 1.39) | 7.0E-10 | 0.59 | 0.660 | 1.22 (1.04 - 1.42) | 1.3E-02 | 0.58 | 0.361 | 1.3 (1.14 - 1.48) | 6.9E-05 | 0.59 | 0.612 |
| 13 | 110838236 | *COL4A1* | rs7326444 | G | A | 1.31 (1.21 - 1.42) | 1.0E-10 | 0.64 | 0.517 | 1.55 (1.31 - 1.84) | 4.2E-07 | 0.65 | 0.456 | 1.26 (1.1 - 1.44) | 9.3E-04 | 0.64 | 0.974 |
| 13 | 111040681 | *COL4A2* | rs11838776 | G | A | 1.5 (1.37 - 1.65) | 2.5E-18 | 0.73 | 0.420 | 1.63 (1.35 - 1.97) | 3.2E-07 | 0.72 | 0.841 | 1.54 (1.32 - 1.8) | 3.7E-08 | 0.73 | 0.890 |
| 15 | 48763754 | *FBN1* | rs7174973 | G | A | 1.54 (1.37 - 1.72) | 1.6E-13 | 0.11 | 0.030 | 1.9 (1.52 - 2.37) | 1.1E-08 | 0.11 | 0.232 | 1.69 (1.41 - 2.02) | 1.1E-08 | 0.11 | 0.485 |
| 15 | 71628370 | *THSD4* | rs10851839 | A | T | 1.32 (1.22 - 1.44) | 5.5E-11 | 0.68 | 0.237 | 1.34 (1.12 - 1.59) | 9.7E-04 | 0.68 | 0.142 | 1.34 (1.17 - 1.55) | 4.1E-05 | 0.67 | 0.550 |
| 21 | 35593827 | *LINC00310/KCNE2* | rs28451064 | G | A | 2.04 (1.77 - 2.35) | 1.2E-22 | 0.88 | 0.497 | 1.71 (1.29 - 2.26) | 1.7E-04 | 0.87 | 0.889 | 2.74 (2.08 - 3.61) | 7.0E-13 | 0.88 | 0.125 |
| 22 | 33282971 | *SYN3* | rs137507 | T | C | 1.38 (1.23 - 1.55) | 3.3E-08 | 0.11 | 0.019 | 1.38 (1.1 - 1.73) | 5.2E-03 | 0.11 | 0.065 | 1.42 (1.17 - 1.71) | 3.0E-04 | 0.11 | 0.248 |
| X | 34489890 | *FAM47A/TMEM47* | rs5973204 | T | C | 1.36 (1.22 - 1.51) | 3.0E-08 | 0.20 | 0.155 | 1.81 (1.41 - 2.32) | 2.9E-06 | 0.19 | 0.378 | 1.47 (1.19 - 1.82) | 2.9E-04 | 0.19 | 0.308 |

**Supplementary Table 10: Genetic correlation between SCAD and CAD conditioned on cardiovascular risk factors and blood traits (mtCOJO)**

rg: genetic correlation, se: standard error, P: P-value for the genetic correlation.

SBP: systolic blood pressure, DBP: diastolic blood pressure, BMI: body mass index , HDL: high density lipoprotein, LDL: low density lipoprotein, TC: total cholesterol, TG: triglycerides, SMK: smoking (never/ever), HGB: haemoglobin, HCT: haematocrit

| Disease or trait | rg | se | L95_rg | U95_rg | z | P |
|---|---|---|---|---|---|---|
| Unadjusted | -0.122 | 0.042 | -0.20 | -0.04 | -2.91 | 3.6E-03 |
| SBP | -0.195 | 0.043 | -0.28 | -0.11 | -4.58 | 4.6E-06 |
| DBP | -0.188 | 0.043 | -0.27 | -0.10 | -4.37 | 1.3E-05 |
| BMI | -0.128 | 0.041 | -0.21 | -0.05 | -3.16 | 1.6E-03 |
| HDL | -0.149 | 0.041 | -0.23 | -0.07 | -3.66 | 2.0E-04 |
| LDL | -0.123 | 0.041 | -0.20 | -0.04 | -3.04 | 2.4E-03 |
| TC | -0.129 | 0.041 | -0.21 | -0.05 | -3.18 | 1.5E-03 |
| TG | -0.141 | 0.041 | -0.22 | -0.06 | -3.46 | 5.0E-04 |
| SMK | -0.123 | 0.041 | -0.20 | -0.04 | -3.04 | 2.4E-03 |
| HGB | -0.127 | 0.041 | -0.21 | -0.05 | -3.08 | 2.1E-03 |
| HCT | -0.126 | 0.041 | -0.21 | -0.05 | -3.07 | 2.2E-03 |

**Supplementary Table 11: Mendelian randomisation (MR) analysis between SCAD or CAD and cardiovascular risk factors and blood traits**

N_SNP : count of SNP used as instrumental variables in the MR analysis, BETA: effect size obtained from MR analysis (IVW: Inverse variance weighted, MR-egger, weighted median), SE: standard error of effect size, P: P-value for the association. P.adj: Bonferonni adjusted P-value.

CAD: coronary artery disease, SCAD : spontaneous coronary artery dissection, SBP: systolic blood pressure, DBP: diastolic blood pressure, BMI: body mass index , HDL: high density lipoprotein, LDL: low density lipoprotein, TC: total cholesterol, TG: triglycerides, SMK: smoking (never/ever), T2D: type 2 diabete, HGB: haemoglobin, HCT: haematocrit

details on used GWAS summary statistics of exposures are available in supplementry table S16

| | Outcome = SCAD | | | | | | | | | | | Outcome = CAD | | | | | | | | | | |
|---|---|---|---|---|---|---|---|---|---|---|---|---|---|---|---|---|---|---|---|---|---|---|
| | | IVW | | | | MR-Egger | | | Weighted median | | | | IVW | | | | Egger | | | Weighted median | | |
| Exposure (risk factor) | N_SNP | BETA | SE | P | P.adj | BETA | SE | P | BETA | SE | P | N_SNP | BETA | SE | P | P.adj | BETA | SE | P | BETA | SE | P |
| SBP | 433 | 0.05 | 0.01 | 7.6E-06 | 8.4E-05 | 0.12 | 0.03 | 2.1E-05 | 0.04 | 0.01 | 1.8E-03 | 497 | 0.04 | 0.002 | 8.6E-49 | 9.5E-48 | 0.04 | 0.01 | 2.8E-09 | 0.03 | 0.002 | 1.9E-50 |
| DBP | 444 | 0.10 | 0.02 | 1.9E-08 | 2.1E-07 | 0.23 | 0.05 | 2.7E-06 | 0.09 | 0.02 | 2.1E-05 | 510 | 0.06 | 0.004 | 1.6E-44 | 1.8E-43 | 0.06 | 0.01 | 1.3E-08 | 0.06 | 0.004 | 9.6E-46 |
| BMI | 276 | 0.00 | 0.03 | 8.9E-01 | 1 | 0.21 | 0.09 | 2.6E-02 | 0.03 | 0.05 | 4.7E-01 | 319 | 0.09 | 0.01 | 3.0E-35 | 3.3E-34 | 0.08 | 0.02 | 4.9E-04 | 0.10 | 0.01 | 5.2E-35 |
| HDL | 175 | -0.89 | 0.36 | 1.4E-02 | 0.154 | -0.01 | 0.67 | 9.9E-01 | -0.55 | 0.52 | 2.9E-01 | 186 | -0.66 | 0.10 | 1.8E-10 | 2E-09 | -0.50 | 0.18 | 5.0E-03 | -0.46 | 0.12 | 1.1E-04 |
| LDL | 98 | -0.23 | 0.22 | 3.0E-01 | 1 | -0.10 | 0.41 | 8.0E-01 | 0.07 | 0.32 | 8.2E-01 | 107 | 0.64 | 0.09 | 7.4E-13 | 8.1E-12 | 1.00 | 0.17 | 3.2E-08 | 0.67 | 0.06 | 1.3E-25 |
| TC | 100 | -0.22 | 0.14 | 1.2E-01 | 1 | -0.08 | 0.24 | 7.3E-01 | -0.08 | 0.19 | 6.8E-01 | 118 | 0.31 | 0.06 | 1.2E-08 | 1.3E-07 | 0.44 | 0.10 | 1.3E-05 | 0.35 | 0.04 | 2.0E-20 |
| TG | 131 | 0.22 | 0.13 | 9.7E-02 | 1 | 0.16 | 0.21 | 4.6E-01 | 0.18 | 0.19 | 3.5E-01 | 150 | 0.31 | 0.05 | 3.9E-11 | 4.3E-10 | 0.19 | 0.07 | 6.5E-03 | 0.24 | 0.04 | 2.8E-10 |
| SMK | 28 | -0.57 | 1.09 | 6.0E-01 | 1 | 4.81 | 6.65 | 4.8E-01 | -0.53 | 1.51 | 7.2E-01 | 35 | 0.37 | 0.18 | 4.4E-02 | 0.484 | 0.43 | 0.79 | 5.9E-01 | 0.43 | 0.24 | 6.7E-02 |
| T2D | 153 | 0.08 | 0.07 | 2.7E-01 | 1 | 0.01 | 0.20 | 9.5E-01 | 0.07 | 0.10 | 5.0E-01 | 162 | 0.12 | 0.02 | 1.6E-10 | 1.8E-09 | 0.03 | 0.04 | 4.8E-01 | 0.09 | 0.02 | 1.5E-07 |
| HGB | 296 | 0.56 | 0.14 | 3.3E-05 | 3.63E-04 | 0.39 | 0.28 | 1.6E-01 | 0.53 | 0.21 | 1.2E-02 | 210 | 0.05 | 0.04 | 2.3E-01 | 1 | -0.14 | 0.06 | 2.6E-02 | -0.03 | 0.04 | 4.4E-01 |
| HCT | 273 | 0.54 | 0.14 | 1.5E-04 | 1.65E-03 | 0.30 | 0.30 | 3.3E-01 | 0.29 | 0.22 | 2.0E-01 | 329 | 0.06 | 0.03 | 7.9E-02 | 0.869 | -0.07 | 0.07 | 2.7E-01 | 0.01 | 0.04 | 7.1E-01 |

**Supplementary Table 12: Mendelian randomisation (MR) analysis between SCAD or CAD and cardiovascular risk factors and blood traits stratified on sex**

N_SNP : count of SNP used as instrumental variables in the MR analysis, BETA: effect size obtained from IVW MR analysis, SE: standard error of effect size, P: P-value for the association.

CAD: coronary artery disease, SCAD : spontaneous coronary artery dissection, SBP: systolic blood pressure, DBP: diastolic blood pressure, BMI: body mass index , HDL: high density lipoprotein, LDL: low density lipoprotein, TC: total cholesterol, TG: triglycerides, SMK: smoking (never/ever), T2D: type 2 diabete, HGB: haemoglobin, HCT: haematocrit

| | Women | | | | | | | | Men | | | |
|---|---|---|---|---|---|---|---|---|---|---|---|---|
| | Outcome = SCAD | | | | Outcome = CAD | | | | Outcome = CAD | | | |
| Exposure (risk factor) | N_SNP | BETA | SE | P | N_SNP | BETA | SE | P | N_SNP | BETA | SE | P |
| SBP | 159 | 0.04 | 0.01 | 5.8E-03 | 185 | 0.04 | 0.004 | 7.9E-23 | 174 | 0.03 | 0.01 | 6.3E-11 |
| DBP | 137 | 0.09 | 0.03 | 7.7E-04 | 163 | 0.06 | 0.01 | 6.4E-17 | 158 | 0.05 | 0.01 | 4.1E-10 |
| BMI | 277 | 0.01 | 0.03 | 8.3E-01 | 333 | 0.09 | 0.01 | 9.9E-26 | 317 | 0.10 | 0.01 | 8.1E-28 |
| HDL | 176 | -0.81 | 0.34 | 1.8E-02 | 198 | -0.59 | 0.11 | 1.1E-07 | 181 | -0.84 | 0.13 | 4.0E-10 |
| LDL | 100 | -0.12 | 0.21 | 5.7E-01 | 109 | 0.48 | 0.09 | 4.4E-08 | 102 | 0.69 | 0.10 | 4.3E-11 |
| TC | 100 | -0.20 | 0.14 | 1.5E-01 | 122 | 0.24 | 0.06 | 1.9E-05 | 111 | 0.39 | 0.07 | 2.9E-09 |
| TG | 128 | 0.25 | 0.15 | 1.0E-01 | 153 | 0.38 | 0.06 | 5.3E-11 | 142 | 0.26 | 0.04 | 2.1E-09 |
| SMK | 28 | -0.23 | 1.07 | 8.3E-01 | 37 | 0.44 | 0.27 | 1.0E-01 | 35 | 0.29 | 0.22 | 1.8E-01 |
| T2D | 153 | 0.09 | 0.07 | 2.2E-01 | 161 | 0.13 | 0.02 | 2.1E-09 | 156 | 0.12 | 0.02 | 2.9E-08 |
| HGB | 172 | 0.53 | 0.19 | 5.6E-03 | 215 | 0.05 | 0.05 | 3.6E-01 | 200 | 0.04 | 0.04 | 4.3E-01 |
| HCT | 168 | 0.15 | 0.06 | 1.3E-02 | 207 | 0.01 | 0.02 | 5.4E-01 | 193 | -0.02 | 0.02 | 3.5E-01 |

**Supplementary Table 13**: **Details of genotyping, pre-imputation and post-imputation quality control steps per study.**
N : number of samples, IBD: Identity by descent , PCA: Principal component analysis.

*UK Biobank controls randomly selected after exclusion for relatedness (K>0.044), heterozygosity and self-reported ethnicity
**Only include UBC samples from CanSCAD
**Only include UBC samples from CanSCAD, exclude U.S
**Age,sex, and ancestry (PC1-PC3) matched controls.

| | | | | | QC steps | | | | | | | | |
|---|---|---|---|---|---|---|---|---|---|---|---|---|---|
| **Country** | **Cohort** | **Status** | **Genotyping array** | **N (genotyped)** | **sub-Cohort selection filter** | **Heterozygosity / Call Rate** | **Relatdness (IBD>0.185)** | **Non European excluded. (PCA analysis)** | **Non SCAD or unknown** | **genetic syndrome cases** | **Sex check** | **N included in GWAS** | |
| FRA | DISCO | Cases | Infinium OmniExpressExome-8v1.3 | 412 | - | 12 | 1 | 41 | 45 | - | - | 313 | |
| | 3 Cities study | Controls | Illumina-Human660W-Quad v1.0 | 1487 | - | 0 | 0 | 0 | NR | - | - | 1487 | |
| UK | SCAD-UK Study I | Cases | Sequenced | 383 | | | | 0 | | | | 383 | |
| | | Controls* | Affymetrix Axiom UK Biobank array* | 1487 | | 0 | 0 | 0 | NR | | | 1919 | |
| UK | SCAD-UK Study II | Cases | Illumina-Infinium Global Screening Array-24 v2.0 +MD | 163 | | 20 | 0 | 0 | 0 | | | 143 | |
| | | Controls* | Affymetrix Axiom UK Biobank array* | 815 | | 0 | 0 | 0 | NR | | | 815 | |
| USA | MAYO | Cases | Infinium OmniExpress-24 v1.2 | 506 | | 0 | 0 | 0 | 0 | | | 506 | |
| | | Controls | Infinium HumanHap550, 610, 660; OmniExpress | 1549 | | 0 | 0 | 0 | NR | | | 1549 | |
| USA | CanSCAD | Case | illumina-Infinium CoreExome-24v1.1 BeadArray with 607,778 SNP markers (UM_HUNT_Biobank_v1-1_20006200_A) | 502 | 49 | 1 | 1 | 88 | 2 | 2 | 2 | 357** | |
| | MGI | Controls | | 13756 | | 0 | 0 | 11631 | NR | | | 2125*** | |
| USA | DEFINE-SCAD Study | Cases | Infinium OmniExpressExome-8v1.6 | 47 | | 1 | 0 | 0 | 30 | | | 16 | 42 |
| | | Cases | Illumina-Human-Omni-Express-Exome | 172 | | 3 | 0 | 0 | 143 | | | 26 | |
| | | Controls | Infinium OmniExpressExome-8v1.6 | 1 | | 0 | 0 | 0 | 0 | | | 1 | 153 |
| | | Controls | Illumina-Human-Omni-Express-Exome | 164 | | 2 | 6 | 0 | 1 | | 3 | 152 | |
| AU | VCCRI | Cases | Whole Genome Sequencing | 88 | | 0 | 0 | 0 | 0 | | | 88 | |
| | | Controls | | 1127 | | 0 | 0 | 0 | NR | | | 1127 | |
| AU | VCCRI2 | Cases | Axiom UK Biobank array-cases Axiom PMDA-controls | 91 | | 0 | 2 | 4 | 0 | | | 85 | |
| | | Controls | | 111 | | 0 | 0 | 0 | NR | | | 111 | |

**Supplementary Table 14: Epigenomic datasets used for annotations of SCAD associated variants.**

| Tissue enrichment (Supplementary figure S3) | | | | |
|---|---|---|---|---|
| **File accession** | **File format** | **Assay** | **Biosample term name** | **Links to Download** |
| ENCFF559GDW | bed narrowPeak | H3K27ac ChIP-seq | spleen | |
| ENCFF714TUQ | bed narrowPeak | H3K27ac ChIP-seq | gastrocnemius medialis | |
| ENCFF271OHD | bed narrowPeak | H3K27ac ChIP-seq | body of pancreas | |
| ENCFF200VYQ | bed narrowPeak | H3K27ac ChIP-seq | upper lobe of left lung | |
| ENCFF635VBV | bed narrowPeak | H3K27ac ChIP-seq | heart left ventricle | |
| ENCFF382EMP | bed narrowPeak | H3K27ac ChIP-seq | right lobe of liver | |
| ENCFF033TXD | bed narrowPeak | H3K27ac ChIP-seq | ascending aorta | |
| ENCFF267KHM | bed narrowPeak | H3K27ac ChIP-seq | gastrocnemius medialis | |
| ENCFF057CRD | bed narrowPeak | H3K27ac ChIP-seq | esophagus squamous epithelium | |
| ENCFF670PZG | bed narrowPeak | H3K27ac ChIP-seq | gastroesophageal sphincter | |
| ENCFF814RAE | bed narrowPeak | H3K27ac ChIP-seq | Peyer's patch | |
| ENCFF731GWD | bed narrowPeak | H3K27ac ChIP-seq | body of pancreas | |
| ENCFF266NWL | bed narrowPeak | H3K27ac ChIP-seq | transverse colon | |
| ENCFF538QNE | bed narrowPeak | H3K27ac ChIP-seq | adrenal gland | |
| ENCFF524AKO | bed narrowPeak | H3K27ac ChIP-seq | ascending aorta | |
| ENCFF231SUE | bed narrowPeak | H3K27ac ChIP-seq | uterus | |
| ENCFF214NTL | bed narrowPeak | H3K27ac ChIP-seq | spleen | |
| ENCFF208DJA | bed narrowPeak | H3K27ac ChIP-seq | sigmoid colon | |
| ENCFF561EZS | bed narrowPeak | H3K27ac ChIP-seq | right atrium auricular region | |
| ENCFF776YOM | bed narrowPeak | H3K27ac ChIP-seq | gastrocnemius medialis | |
| ENCFF739QLX | bed narrowPeak | H3K27ac ChIP-seq | tibial nerve | |
| ENCFF004ANL | bed narrowPeak | H3K27ac ChIP-seq | esophagus muscularis mucosa | |
| ENCFF703WKZ | bed narrowPeak | H3K27ac ChIP-seq | tibial nerve | |
| ENCFF394NUN | bed narrowPeak | H3K27ac ChIP-seq | adrenal gland | |
| ENCFF417GYW | bed narrowPeak | H3K27ac ChIP-seq | thoracic aorta | |
| ENCFF289XOR | bed narrowPeak | H3K27ac ChIP-seq | skin epidermis | |
| ENCFF087WUV | bed narrowPeak | H3K27ac ChIP-seq | skin epidermis | |
| ENCFF489GFM | bed narrowPeak | H3K27ac ChIP-seq | skin epidermis | |
| ENCFF683QWQ | bed narrowPeak | H3K27ac ChIP-seq | left lung | |
| ENCFF758DTU | bed narrowPeak | H3K27ac ChIP-seq | spleen | |
| ENCFF877IEN | bed narrowPeak | H3K27ac ChIP-seq | adrenal gland | |
| ENCFF317ENJ | bed narrowPeak | H3K27ac ChIP-seq | adrenal gland | |
| ENCFF770HCQ | bed narrowPeak | H3K27ac ChIP-seq | pancreas | |
| ENCFF862CEW | bed narrowPeak | H3K27ac ChIP-seq | spleen | |

| | | | |
|---|---|---|---|
| ENCFF875LMK | bed narrowPeak | H3K27ac ChIP-seq | skin epidermis |
| ENCFF987KRH | bed narrowPeak | H3K27ac ChIP-seq | skin epidermis |
| ENCFF677BHR | bed narrowPeak | H3K27ac ChIP-seq | chorionic villus |
| ENCFF101BIB | bed narrowPeak | H3K27ac ChIP-seq | gastroesophageal sphincter |
| ENCFF988ZYW | bed narrowPeak | H3K27ac ChIP-seq | breast epithelium |
| ENCFF322YIK | bed narrowPeak | H3K27ac ChIP-seq | spleen |
| ENCFF111SYA | bed narrowPeak | H3K27ac ChIP-seq | sigmoid colon |
| ENCFF787TWQ | bed narrowPeak | H3K27ac ChIP-seq | gastroesophageal sphincter |
| ENCFF379RWI | bed narrowPeak | H3K27ac ChIP-seq | skin epidermis |
| ENCFF257VPC | bed narrowPeak | H3K27ac ChIP-seq | stomach |
| ENCFF285RPL | bed narrowPeak | H3K27ac ChIP-seq | ovary |
| ENCFF219SMW | bed narrowPeak | H3K27ac ChIP-seq | chorionic villus |
| ENCFF477MXB | bed narrowPeak | H3K27ac ChIP-seq | placental basal plate |
| ENCFF105SAZ | bed narrowPeak | H3K27ac ChIP-seq | temporal lobe |
| ENCFF722LRM | bed narrowPeak | H3K27ac ChIP-seq | stomach smooth muscle |
| ENCFF874GTL | bed narrowPeak | H3K27ac ChIP-seq | placenta |
| ENCFF645UIY | bed narrowPeak | H3K27ac ChIP-seq | chorionic villus |
| ENCFF286ERH | bed narrowPeak | H3K27ac ChIP-seq | chorion |
| ENCFF382PYP | bed narrowPeak | H3K27ac ChIP-seq | mucosa of rectum |
| ENCFF177IAO | bed narrowPeak | H3K27ac ChIP-seq | trophoblast |
| ENCFF398EEO | bed narrowPeak | H3K27ac ChIP-seq | suprapubic skin |
| ENCFF310DII | bed narrowPeak | H3K27ac ChIP-seq | Peyer's patch |
| ENCFF337KNQ | bed narrowPeak | H3K27ac ChIP-seq | coronary artery |
| ENCFF230KBG | bed narrowPeak | H3K27ac ChIP-seq | skin epidermis |
| ENCFF860TAY | bed narrowPeak | H3K27ac ChIP-seq | middle frontal area 46 |
| ENCFF169QSG | bed narrowPeak | H3K27ac ChIP-seq | middle frontal area 46 |
| ENCFF165SPP | bed narrowPeak | H3K27ac ChIP-seq | vagina |
| ENCFF423CQD | bed narrowPeak | H3K27ac ChIP-seq | skin epidermis |
| ENCFF337DEA | bed narrowPeak | H3K27ac ChIP-seq | middle frontal area 46 |
| ENCFF899KVD | bed narrowPeak | H3K27ac ChIP-seq | spleen |
| ENCFF507WEL | bed narrowPeak | H3K27ac ChIP-seq | breast epithelium |
| ENCFF730HDE | bed narrowPeak | H3K27ac ChIP-seq | middle frontal area 46 |
| ENCFF078CLH | bed narrowPeak | H3K27ac ChIP-seq | thoracic aorta |
| ENCFF595JKW | bed narrowPeak | H3K27ac ChIP-seq | middle frontal area 46 |
| ENCFF280JLL | bed narrowPeak | H3K27ac ChIP-seq | adrenal gland |
| ENCFF970DMJ | bed narrowPeak | H3K27ac ChIP-seq | right atrium auricular region |
| ENCFF007OIY | bed narrowPeak | H3K27ac ChIP-seq | heart left ventricle |
| ENCFF797XLL | bed narrowPeak | H3K27ac ChIP-seq | heart right ventricle |
| ENCFF014ZMR | bed narrowPeak | H3K27ac ChIP-seq | sigmoid colon |
| ENCFF751RTT | bed narrowPeak | H3K27ac ChIP-seq | thyroid gland |

| | | | |
|---|---|---|---|
| ENCFF910HDI | bed narrowPeak | H3K27ac ChIP-seq | stomach |
| ENCFF577QNA | bed narrowPeak | H3K27ac ChIP-seq | vagina |
| ENCFF987GGX | bed narrowPeak | H3K27ac ChIP-seq | body of pancreas |
| ENCFF283HUT | bed narrowPeak | H3K27ac ChIP-seq | middle frontal area 46 |
| ENCFF270GEZ | bed narrowPeak | H3K27ac ChIP-seq | thyroid gland |
| ENCFF336ETH | bed narrowPeak | H3K27ac ChIP-seq | heart left ventricle |
| ENCFF945XND | bed narrowPeak | H3K27ac ChIP-seq | tibial artery |
| ENCFF933DUA | bed narrowPeak | H3K27ac ChIP-seq | middle frontal area 46 |
| ENCFF736SWI | bed narrowPeak | H3K27ac ChIP-seq | stomach |
| ENCFF108SAD | bed narrowPeak | H3K27ac ChIP-seq | uterus |
| ENCFF216XRU | bed narrowPeak | H3K27ac ChIP-seq | middle frontal area 46 |
| ENCFF124JXP | bed narrowPeak | H3K27ac ChIP-seq | middle frontal area 46 |
| ENCFF448PFH | bed narrowPeak | H3K27ac ChIP-seq | lower lobe of left lung |
| ENCFF346GZD | bed narrowPeak | H3K27ac ChIP-seq | skin epidermis |
| ENCFF924OGR | bed narrowPeak | H3K27ac ChIP-seq | skin epidermis |
| ENCFF600QTP | bed narrowPeak | H3K27ac ChIP-seq | heart right ventricle |
| ENCFF045ADI | bed narrowPeak | H3K27ac ChIP-seq | middle frontal area 46 |
| ENCFF714JDQ | bed narrowPeak | H3K27ac ChIP-seq | pancreas |
| ENCFF721ZGP | bed narrowPeak | H3K27ac ChIP-seq | middle frontal area 46 |
| ENCFF462XVH | bed narrowPeak | H3K27ac ChIP-seq | skin epidermis |
| ENCFF861YME | bed narrowPeak | H3K27ac ChIP-seq | middle frontal area 46 |
| ENCFF654QRM | bed narrowPeak | H3K27ac ChIP-seq | body of pancreas |
| ENCFF114SOB | bed narrowPeak | H3K27ac ChIP-seq | esophagus muscularis mucosa |
| ENCFF882VLE | bed narrowPeak | H3K27ac ChIP-seq | upper lobe of left lung |
| ENCFF003TGC | bed narrowPeak | H3K27ac ChIP-seq | skin epidermis |
| ENCFF214YFB | bed narrowPeak | H3K27ac ChIP-seq | heart left ventricle |
| ENCFF229NLK | bed narrowPeak | H3K27ac ChIP-seq | middle frontal area 46 |
| ENCFF847EPP | bed narrowPeak | H3K27ac ChIP-seq | esophagus squamous epithelium |
| ENCFF024XNY | bed narrowPeak | H3K27ac ChIP-seq | middle frontal area 46 |
| ENCFF712CHB | bed narrowPeak | H3K27ac ChIP-seq | middle frontal area 46 |
| ENCFF528AIU | bed narrowPeak | H3K27ac ChIP-seq | middle frontal area 46 |
| ENCFF496RDA | bed narrowPeak | H3K27ac ChIP-seq | upper lobe of left lung |
| ENCFF253VVF | bed narrowPeak | H3K27ac ChIP-seq | esophagus squamous epithelium |
| ENCFF032CSL | bed narrowPeak | H3K27ac ChIP-seq | testis |
| ENCFF474VLR | bed narrowPeak | H3K27ac ChIP-seq | middle frontal area 46 |
| ENCFF581YQU | bed narrowPeak | H3K27ac ChIP-seq | middle frontal area 46 |
| ENCFF960JWJ | bed narrowPeak | H3K27ac ChIP-seq | esophagus muscularis mucosa |
| ENCFF549AXK | bed narrowPeak | H3K27ac ChIP-seq | colonic mucosa |
| ENCFF189UQY | bed narrowPeak | H3K27ac ChIP-seq | middle frontal area 46 |
| ENCFF380YWT | bed narrowPeak | H3K27ac ChIP-seq | heart left ventricle |

| | | | | |
|---|---|---|---|---|
| ENCFF283LVU | bed narrowPeak | H3K27ac ChIP-seq | middle frontal area 46 | |
| ENCFF693JGW | bed narrowPeak | H3K27ac ChIP-seq | middle frontal area 46 | |
| ENCFF017NRE | bed narrowPeak | H3K27ac ChIP-seq | gastroesophageal sphincter | |
| ENCFF381SFJ | bed narrowPeak | H3K27ac ChIP-seq | upper lobe of left lung | |
| ENCFF756JDB | bed narrowPeak | H3K27ac ChIP-seq | middle frontal area 46 | |
| ENCFF521HKS | bed narrowPeak | H3K27ac ChIP-seq | heart right ventricle | |
| ENCFF110LKJ | bed narrowPeak | H3K27ac ChIP-seq | parathyroid adenoma | |
| ENCFF010VMY | bed narrowPeak | H3K27ac ChIP-seq | middle frontal area 46 | |
| ENCFF089YVE | bed narrowPeak | H3K27ac ChIP-seq | middle frontal area 46 | |
| ENCFF300SDH | bed narrowPeak | H3K27ac ChIP-seq | middle frontal area 46 | |
| ENCFF570DDI | bed narrowPeak | H3K27ac ChIP-seq | heart right ventricle | |
| ENCFF319KZD | bed narrowPeak | H3K27ac ChIP-seq | middle frontal area 46 | |
| ENCFF717AYR | bed narrowPeak | H3K27ac ChIP-seq | tibial nerve | |
| ENCFF761QUK | bed narrowPeak | H3K27ac ChIP-seq | middle frontal area 46 | |
| ENCFF936SMV | bed narrowPeak | H3K27ac ChIP-seq | Peyer's patch | |
| ENCFF540YUQ | bed narrowPeak | H3K27ac ChIP-seq | testis | |
| ENCFF970PFD | bed narrowPeak | H3K27ac ChIP-seq | middle frontal area 46 | |
| ENCFF749JEY | bed narrowPeak | H3K27ac ChIP-seq | heart right ventricle | |
| ENCFF142COP | bed narrowPeak | H3K27ac ChIP-seq | transverse colon | |
| ENCFF487FOU | bed narrowPeak | H3K27ac ChIP-seq | adrenal gland | |
| ENCFF128AQO | bed narrowPeak | H3K27ac ChIP-seq | middle frontal area 46 | |
| ENCFF787FST | bed narrowPeak | H3K27ac ChIP-seq | skin epidermis | https://www.encodeproject.org/ |
| ENCFF240EGF | bed narrowPeak | H3K27ac ChIP-seq | skin epidermis | |
| ENCFF499HFY | bed narrowPeak | H3K27ac ChIP-seq | heart left ventricle | |
| ENCFF579OOI | bed narrowPeak | H3K27ac ChIP-seq | Peyer's patch | |
| ENCFF600DLN | bed narrowPeak | H3K27ac ChIP-seq | skin epidermis | |
| ENCFF895PGR | bed narrowPeak | H3K27ac ChIP-seq | skin epidermis | |
| ENCFF441AHV | bed narrowPeak | H3K27ac ChIP-seq | tibial nerve | |
| ENCFF676LJR | bed narrowPeak | H3K27ac ChIP-seq | middle frontal area 46 | |
| ENCFF523WDP | bed narrowPeak | H3K27ac ChIP-seq | tibial artery | |
| ENCFF205LFV | bed narrowPeak | H3K27ac ChIP-seq | middle frontal area 46 | |
| ENCFF978AOD | bed narrowPeak | H3K27ac ChIP-seq | stomach | |
| ENCFF294GNO | bed narrowPeak | H3K27ac ChIP-seq | transverse colon | |
| ENCFF673PCP | bed narrowPeak | H3K27ac ChIP-seq | esophagus squamous epithelium | |
| ENCFF639KHG | bed narrowPeak | H3K27ac ChIP-seq | skin epidermis | |
| ENCFF805FFP | bed narrowPeak | H3K27ac ChIP-seq | spleen | |
| ENCFF913SWW | bed narrowPeak | H3K27ac ChIP-seq | heart right ventricle | |
| ENCFF065ZDX | bed narrowPeak | H3K27ac ChIP-seq | thyroid gland | |
| ENCFF984YUQ | bed narrowPeak | H3K27ac ChIP-seq | coronary artery | |
| ENCFF457YRY | bed narrowPeak | H3K27ac ChIP-seq | placental basal plate | |

| | | | |
|---|---|---|---|
| ENCFF095OXH | bed narrowPeak | H3K27ac ChIP-seq | heart left ventricle |
| ENCFF083HKS | bed narrowPeak | H3K27ac ChIP-seq | skin epidermis |
| ENCFF551MWX | bed narrowPeak | H3K27ac ChIP-seq | prostate gland |
| ENCFF087FDQ | bed narrowPeak | H3K27ac ChIP-seq | gastrocnemius medialis |
| ENCFF368OSK | bed narrowPeak | H3K27ac ChIP-seq | transverse colon |
| ENCFF414SKU | bed narrowPeak | H3K27ac ChIP-seq | sigmoid colon |
| ENCFF594VVB | bed narrowPeak | H3K27ac ChIP-seq | skin epidermis |
| ENCFF143DRH | bed narrowPeak | H3K27ac ChIP-seq | skin epidermis |
| ENCFF138WNV | bed narrowPeak | H3K27ac ChIP-seq | thyroid gland |
| ENCFF498SSN | bed narrowPeak | H3K27ac ChIP-seq | pancreas |
| ENCFF788DLD | bed narrowPeak | H3K27ac ChIP-seq | middle frontal area 46 |
| ENCFF610VHP | bed narrowPeak | H3K27ac ChIP-seq | pancreas |
| ENCFF245RGA | bed narrowPeak | H3K27ac ChIP-seq | heart right ventricle |
| ENCFF034GZW | bed narrowPeak | H3K27ac ChIP-seq | skeletal muscle tissue |
| ENCFF173ZPC | bed narrowPeak | H3K27ac ChIP-seq | mucosa of rectum |
| ENCFF937WTH | bed narrowPeak | H3K27ac ChIP-seq | caudate nucleus |
| ENCFF737HGQ | bed narrowPeak | H3K27ac ChIP-seq | kidney |
| ENCFF236DDT | bed narrowPeak | H3K27ac ChIP-seq | temporal lobe |
| ENCFF066PAQ | bed narrowPeak | H3K27ac ChIP-seq | layer of hippocampus |
| ENCFF703WCD | bed narrowPeak | H3K27ac ChIP-seq | middle frontal area 46 |
| ENCFF056SCA | bed narrowPeak | H3K27ac ChIP-seq | rectal smooth muscle tissue |
| ENCFF089APD | bed narrowPeak | H3K27ac ChIP-seq | colonic mucosa |
| ENCFF778VTG | bed narrowPeak | H3K27ac ChIP-seq | placenta |
| ENCFF745JHJ | bed narrowPeak | H3K27ac ChIP-seq | trophoblast |
| ENCFF016AAS | bed narrowPeak | H3K27ac ChIP-seq | skin epidermis |
| ENCFF014OZD | bed narrowPeak | H3K27ac ChIP-seq | lower lobe of left lung |
| ENCFF729IPL | bed narrowPeak | H3K27ac ChIP-seq | skin epidermis |
| ENCFF925RBS | bed narrowPeak | H3K27ac ChIP-seq | middle frontal area 46 |
| ENCFF658NHX | bed narrowPeak | H3K27ac ChIP-seq | subcutaneous abdominal adipose tissue |
| ENCFF552UIM | bed narrowPeak | H3K27ac ChIP-seq | chorionic villus |
| ENCFF805YRQ | bed narrowPeak | H3K27ac ChIP-seq | liver |
| ENCFF155FWO | bed narrowPeak | H3K27ac ChIP-seq | middle frontal area 46 |
| ENCFF201VZW | bed narrowPeak | H3K27ac ChIP-seq | prostate gland |
| ENCFF275GAS | bed narrowPeak | H3K27ac ChIP-seq | large intestine |
| ENCFF982LEG | bed narrowPeak | H3K27ac ChIP-seq | esophagus muscularis mucosa |
| ENCFF341BPG | bed narrowPeak | H3K27ac ChIP-seq | colonic mucosa |
| ENCFF150MNN | bed narrowPeak | H3K27ac ChIP-seq | layer of hippocampus |
| ENCFF551LSF | bed narrowPeak | H3K27ac ChIP-seq | layer of hippocampus |
| ENCFF857JHM | bed narrowPeak | H3K27ac ChIP-seq | muscle layer of duodenum |
| ENCFF516MFW | bed narrowPeak | H3K27ac ChIP-seq | spinal cord |

| | | | |
|---|---|---|---|
| ENCFF659NCA | bed narrowPeak | H3K27ac ChIP-seq | endocrine pancreas |
| ENCFF340JNX | bed narrowPeak | H3K27ac ChIP-seq | amnion |
| ENCFF062NKK | bed narrowPeak | H3K27ac ChIP-seq | pancreas |
| ENCFF592GQB | bed narrowPeak | H3K27ac ChIP-seq | esophagus |
| ENCFF066ROS | bed narrowPeak | H3K27ac ChIP-seq | stomach |
| ENCFF153OUB | bed narrowPeak | H3K27ac ChIP-seq | right cardiac atrium |
| ENCFF287VIA | bed narrowPeak | H3K27ac ChIP-seq | liver |
| ENCFF906EQK | bed narrowPeak | H3K27ac ChIP-seq | heart right ventricle |
| ENCFF718QPZ | bed narrowPeak | H3K27ac ChIP-seq | thymus |
| ENCFF859UCD | bed narrowPeak | H3K27ac ChIP-seq | muscle of trunk |
| ENCFF618CUQ | bed narrowPeak | H3K27ac ChIP-seq | small intestine |
| ENCFF466EWZ | bed narrowPeak | H3K27ac ChIP-seq | muscle layer of colon |
| ENCFF208GHP | bed narrowPeak | H3K27ac ChIP-seq | urinary bladder |
| ENCFF450OXP | bed narrowPeak | H3K27ac ChIP-seq | pancreas |
| ENCFF672SGH | bed narrowPeak | H3K27ac ChIP-seq | middle frontal area 46 |
| ENCFF610WJV | bed narrowPeak | H3K27ac ChIP-seq | heart left ventricle |
| ENCFF401XVC | bed narrowPeak | H3K27ac ChIP-seq | psoas muscle |
| ENCFF824TUN | bed narrowPeak | H3K27ac ChIP-seq | heart right ventricle |
| ENCFF445XCF | bed narrowPeak | H3K27ac ChIP-seq | esophagus |
| ENCFF329CDU | bed narrowPeak | H3K27ac ChIP-seq | heart right ventricle |
| ENCFF037RUM | bed narrowPeak | H3K27ac ChIP-seq | heart left ventricle |
| ENCFF231VWX | bed narrowPeak | H3K27ac ChIP-seq | sigmoid colon |
| ENCFF346BOQ | bed narrowPeak | H3K27ac ChIP-seq | adrenal gland |
| ENCFF654RND | bed narrowPeak | H3K27ac ChIP-seq | heart left ventricle |
| ENCFF872ZJD | bed narrowPeak | H3K27ac ChIP-seq | middle frontal area 46 |
| ENCFF129DPW | bed narrowPeak | H3K27ac ChIP-seq | adipose tissue |
| ENCFF994ZGB | bed narrowPeak | H3K27ac ChIP-seq | aorta |
| ENCFF145NKN | bed narrowPeak | H3K27ac ChIP-seq | spleen |
| ENCFF372OTI | bed narrowPeak | H3K27ac ChIP-seq | stomach |
| ENCFF231NZU | bed narrowPeak | H3K27ac ChIP-seq | adrenal gland |
| ENCFF154ZCF | bed narrowPeak | H3K27ac ChIP-seq | muscle of leg |
| ENCFF872XZM | bed narrowPeak | H3K27ac ChIP-seq | aorta |
| ENCFF340SAM | bed narrowPeak | H3K27ac ChIP-seq | left ventricle myocardium inferior |
| ENCFF954CRD | bed narrowPeak | H3K27ac ChIP-seq | middle frontal area 46 |
| ENCFF059ERB | bed narrowPeak | H3K27ac ChIP-seq | heart left ventricle |
| ENCFF668BZJ | bed narrowPeak | H3K27ac ChIP-seq | thymus |
| ENCFF023HKA | bed narrowPeak | H3K27ac ChIP-seq | psoas muscle |
| ENCFF311BUM | bed narrowPeak | H3K27ac ChIP-seq | middle frontal area 46 |
| ENCFF610ZPW | bed narrowPeak | H3K27ac ChIP-seq | middle frontal area 46 |
| ENCFF307QYO | bed narrowPeak | H3K27ac ChIP-seq | middle frontal area 46 |

| | | | | |
|---|---|---|---|---|
| ENCFF104GTG | bed narrowPeak | H3K27ac ChIP-seq | heart left ventricle | |
| ENCFF442MTS | bed narrowPeak | H3K27ac ChIP-seq | heart right ventricle | |
| ENCFF646VQX | bed narrowPeak | H3K27ac ChIP-seq | parathyroid adenoma | |
| ENCFF681YKY | bed narrowPeak | H3K27ac ChIP-seq | middle frontal area 46 | |
| ENCFF846LAZ | bed narrowPeak | H3K27ac ChIP-seq | left lung | |
| ENCFF927GAF | bed narrowPeak | H3K27ac ChIP-seq | spleen | |
| ENCFF745ROB | bed narrowPeak | H3K27ac ChIP-seq | middle frontal area 46 | |
| ENCFF508PQT | bed narrowPeak | H3K27ac ChIP-seq | middle frontal area 46 | |
| ENCFF272ROP | bed narrowPeak | H3K27ac ChIP-seq | middle frontal area 46 | |
| ENCFF384SYD | bed narrowPeak | H3K27ac ChIP-seq | middle frontal area 46 | |
| ENCFF595CLG | bed narrowPeak | H3K27ac ChIP-seq | middle frontal area 46 | |
| ENCFF297NSU | bed narrowPeak | H3K27ac ChIP-seq | placenta | |
| ENCFF922GGA | bed narrowPeak | H3K27ac ChIP-seq | duodenal mucosa | |
| ENCFF444CZR | bed narrowPeak | H3K27ac ChIP-seq | caudate nucleus | |
| ENCFF418XDA | bed narrowPeak | H3K27ac ChIP-seq | kidney | |
| ENCFF072GKG | bed narrowPeak | H3K27ac ChIP-seq | angular gyrus | |
| ENCFF193GDV | bed narrowPeak | H3K27ac ChIP-seq | liver | |
| ENCFF100WAF | bed narrowPeak | H3K27ac ChIP-seq | cingulate gyrus | |
| ENCFF650YIZ | bed narrowPeak | H3K27ac ChIP-seq | cingulate gyrus | |
| ENCFF976WTG | bed narrowPeak | H3K27ac ChIP-seq | endocrine pancreas | |
| ENCFF008PSH | bed narrowPeak | H3K27ac ChIP-seq | stomach | |
| ENCFF860MVH | bed narrowPeak | H3K27ac ChIP-seq | middle frontal area 46 | |
| ENCFF791TAO | bed narrowPeak | H3K27ac ChIP-seq | substantia nigra | |
| ENCFF026YOC | bed narrowPeak | H3K27ac ChIP-seq | chorion | |
| ENCFF684COM | bed narrowPeak | H3K27ac ChIP-seq | angular gyrus | |
| ENCFF273NFQ | bed narrowPeak | H3K27ac ChIP-seq | spleen | |
| ENCFF761PXE | bed narrowPeak | H3K27ac ChIP-seq | adrenal gland | |
| ENCFF574FYX | bed narrowPeak | H3K27ac ChIP-seq | small intestine | |
| ENCFF328KKO | bed narrowPeak | H3K27ac ChIP-seq | ovary | |
| ENCFF464RXV | bed narrowPeak | H3K27ac ChIP-seq | psoas muscle | |
| ENCFF015DHA | bed narrowPeak | H3K27ac ChIP-seq | spleen | |
| ENCFF759DJM | bed narrowPeak | H3K27ac ChIP-seq | sigmoid colon | |
| ENCFF965GSQ | bed narrowPeak | H3K27ac ChIP-seq | stomach | |
| ENCFF898XDK | bed narrowPeak | H3K27ac ChIP-seq | small intestine | |
| ENCFF074WDV | bed narrowPeak | H3K27ac ChIP-seq | small intestine | |
| ENCFF314QTE | bed narrowPeak | H3K27ac ChIP-seq | lung | |
| ENCFF652JPJ | bed narrowPeak | H3K27ac ChIP-seq | lung | |
| **snATAC-Seq enrichment (Figure 2, Supplementary Figure 4)** | | | | |
| **Cell type** | **File format** | **Type** | **closest Cell Ontology term(s)** | **Link to download** |
| Fibroblast (General) | bed | snATAC-Seq | fibroblast | |

| | | | |
|---|---|---|---|
| Fibroblast (Epithelial) | bed | snATAC-Seq | skin fibroblast |
| Fibroblast (Gastrointestinal) | bed | snATAC-Seq | fibroblast |
| Adipocyte | bed | snATAC-Seq | fat cell |
| Mesothelial Cell | bed | snATAC-Seq | mesothelial cell |
| Fibroblast (Peripheral Nerve) | bed | snATAC-Seq | fibroblast |
| Fibroblast (Sk Muscle Associated) | bed | snATAC-Seq | skeletal muscle fibroblast |
| Satellite Cell | bed | snATAC-Seq | skeletal muscle satellite cell |
| Fibroblast (Liver Adrenal) | bed | snATAC-Seq | fibroblast |
| Thyroid Follicular Cell | bed | snATAC-Seq | thyroid follicular cell |
| Pancreatic Acinar Cell | bed | snATAC-Seq | pancreatic acinar cell |
| T Lymphocyte 1 (CD8+) | bed | snATAC-Seq | CD8-positive, alpha-beta T cell |
| T lymphocyte 2 (CD4+) | bed | snATAC-Seq | CD4-positive, alpha-beta T cell |
| Natural Killer T Cell | bed | snATAC-Seq | mature NK T cell |
| Naive T cell | bed | snATAC-Seq | naive t cell |
| Cardiac Pericyte 1 | bed | snATAC-Seq | pericyte cell |
| Pericyte (General) 1 | bed | snATAC-Seq | pericyte cell |
| Pericyte (General) 2 | bed | snATAC-Seq | pericyte cell |
| Pericyte (General) 3 | bed | snATAC-Seq | pericyte cell |
| Cardiac Pericyte 2 | bed | snATAC-Seq | pericyte cell |
| Pericyte (Esophageal Muscularis) | bed | snATAC-Seq | pericyte cell |
| Pericyte (General) 4 | bed | snATAC-Seq | pericyte cell |
| Cardiac Pericyte 3 | bed | snATAC-Seq | pericyte cell |
| Cardiac Pericyte 4 | bed | snATAC-Seq | pericyte cell |
| Esophageal Epithelial Cell | bed | snATAC-Seq | non keratinizing barrier epithelial cell |
| Pancreatic Beta Cell 1 | bed | snATAC-Seq | type B pancreatic cell |
| Pancreatic Alpha Cell 1 | bed | snATAC-Seq | pancreatic A cell |
| Pancreatic Beta Cell 2 | bed | snATAC-Seq | type B pancreatic cell |
| Pancreatic Delta,Gamma cell | bed | snATAC-Seq | pancreatic D cell, pancreatic PP cell |
| Pancreatic Alpha Cell 2 | bed | snATAC-Seq | pancreatic A cell |
| Gastric Neuroendocrine Cell | bed | snATAC-Seq | stomach neuroendocrine cell |
| Alveolar Type 2 (AT2) Cell | bed | snATAC-Seq | type II pneumocyte |
| Alveolar Type 1 (AT1) Cell | bed | snATAC-Seq | type I pneumocyte |
| Alverolar Type 2,Immune | bed | snATAC-Seq | type II pneumocyte |
| Club Cell | bed | snATAC-Seq | club cell |
| Cilliated Cell | bed | snATAC-Seq | ciliated cell |
| Mammary Luminal Epithelial Cell 1 | bed | snATAC-Seq | luminal epithelial cell of mammary glan |
| Basal Epithelial (Mammary) | bed | snATAC-Seq | basal cell |
| Granular Epidermal (Skin) | bed | snATAC-Seq | granular cell of epidermis |
| Mammary Luminal Epithelial Cell 2 | bed | snATAC-Seq | luminal epithelial cell of mammary glan |
| Eccrine Epidermal (Skin) | bed | snATAC-Seq | eccrine cell |

| | | | | |
|---|---|---|---|---|
| Airway Goblet Cell | bed | snATAC-Seq | respiratory goblet cell | |
| Basal Epidermal (Skin) | bed | snATAC-Seq | basal cell of epidermis | |
| Chief Cell | bed | snATAC-Seq | peptic cell | |
| Parietal Cell | bed | snATAC-Seq | parietal cell | |
| Foveolar Cell | bed | snATAC-Seq | foveolar cell of stomach | |
| Schwann Cell (General) | bed | snATAC-Seq | schwann cell | |
| Melanocyte | bed | snATAC-Seq | melanocyte | |
| Macrophage (General) | bed | snATAC-Seq | macrophage | |
| Macrophage (General,Alveolar) | bed | snATAC-Seq | alveolar macrophage | |
| Microglia | bed | snATAC-Seq | microglial cell | |
| Glutamatergic Neuron 1 | bed | snATAC-Seq | glutamatergic neuron | |
| Glutamatergic Neuron 2 | bed | snATAC-Seq | glutamatergic neuron | |
| GABAergic Neuron 1 | bed | snATAC-Seq | gabaergic neuron | |
| GABAergic Neuron 2 | bed | snATAC-Seq | gabaergic neuron | |
| CNS,Enteric Neuron | bed | snATAC-Seq | enteric neuron | http://renlab.sdsc.edu/kai/Key_Processed_Data/Peaks/ |
| Keratinocyte 1 | bed | snATAC-Seq | keratinocyte | |
| Keratinocyte 2 | bed | snATAC-Seq | keratinocyte | |
| Transitional Zone Cortical Cell | bed | snATAC-Seq | cortical cell of adrenal gland | |
| Zona Fasciculata Cortical Cell | bed | snATAC-Seq | type II cell of adrenal cortex | |
| Zona Glomerulosa Cortical Cell | bed | snATAC-Seq | type I cell of adrenal cortex | |
| Cortical Epithelial-like | bed | snATAC-Seq | cortical cell of adrenal gland | |
| Mammary Epithelial | bed | snATAC-Seq | mammary gland epithelial cell | |
| Myoepithelial (Skin) | bed | snATAC-Seq | myoepithelial cell | |
| Luteal Cell (Ovarian) | bed | snATAC-Seq | luteal cell | |
| Plasma Cell | bed | snATAC-Seq | plasma cell | |
| Memory B Cell | bed | snATAC-Seq | memory b cell | |
| Hepatocyte | bed | snATAC-Seq | hepatocyte | |
| Oligodendrocyte Precursor | bed | snATAC-Seq | oligodendrocyte precursor cell | |
| Astrocyte 1 | bed | snATAC-Seq | astrocyte | |
| Astrocyte 2 | bed | snATAC-Seq | astrocyte | |
| Ductal Cell (Pancreatic) | bed | snATAC-Seq | pancreatic ductal cell | |
| Mast Cell | bed | snATAC-Seq | mast cell | |
| Ventricular Cardiomyocyte | bed | snATAC-Seq | ventricular cardiac muscle cell | |
| Atrial Cardiomyocyte | bed | snATAC-Seq | regular atrial cardiac myocyte | |
| Oligodendrocyte | bed | snATAC-Seq | oligodendrocyte | |
| Endothelial Cell (General) 1 | bed | snATAC-Seq | endothelial cell | |
| Endothelial Cell (Myocardial) | bed | snATAC-Seq | cardiac endothelial cell | |
| Lymphatic Endothelial Cell | bed | snATAC-Seq | endothelial cell of lymphatic vessel | |
| Endothelial Cell (General) 2 | bed | snATAC-Seq | endothelial cell | |
| Alveolar Capillary Endothelial Cell | bed | snATAC-Seq | endothelial cell | |

| | | | | |
|---|---|---|---|---|
| Endothelial Cell (General) 3 | bed | snATAC-Seq | endothelial cell | |
| Endocardial Cell | bed | snATAC-Seq | endocardial cell | |
| Endothelial (Exocrine Tissues) | bed | snATAC-Seq | endothelial cell | |
| Blood Brain Barrier Endothelial Cell | bed | snATAC-Seq | endothelial cell | |
| Smooth Muscle (Esophageal Muscul | bed | snATAC-Seq | smooth muscle cell of the esophagus | |
| Smooth Muscle (Vaginal) | bed | snATAC-Seq | smooth muscle cell | |
| Smooth Muscle (Esophageal Mucosa | bed | snATAC-Seq | smooth muscle cell | |
| Smooth Muscle (Colon) 1 | bed | snATAC-Seq | enteric smooth muscle cell | |
| Smooth Muscle (Esophageal Muscul | bed | snATAC-Seq | smooth muscle cell of the esophagus | |
| Smooth Muscle (Uterine) | bed | snATAC-Seq | uterine smooth muscle cell | |
| Smooth Muscle (General) | bed | snATAC-Seq | smooth muscle cell | |
| Smooth Muscle (GE Junction) | bed | snATAC-Seq | smooth muscle cell | |
| Smooth Muscle (Colon) 2 | bed | snATAC-Seq | enteric smooth muscle cell | |
| Smooth Muscle (Esophageal Muscul | bed | snATAC-Seq | smooth muscle cell | |
| Smooth Muscle (General Gastrointe | bed | snATAC-Seq | smooth muscle cell | |
| Type I Skeletal Myocyte | bed | snATAC-Seq | type I muscle cell | |
| Type II Skeletal Myocyte | bed | snATAC-Seq | type II muscle cell | |
| Vascular Smooth Muscle 1 | bed | snATAC-Seq | blood vessel smooth muscle cell | |
| Vascular Smooth Muscle 2 | bed | snATAC-Seq | blood vessel smooth muscle cell | |
| Cardiac Fibroblasts | bed | snATAC-Seq | fibroblast of cardiac tissue | |
| Peripheral Nerve Stromal | bed | snATAC-Seq | stromal cell | |
| Colon Epithelial Cell 1 | bed | snATAC-Seq | colon epithelial cell | |
| Small Intestinal Enterocyte | bed | snATAC-Seq | enterocyte of epithelium of small intest | |
| Colonic Goblet Cell | bed | snATAC-Seq | colon goblet cell | |
| Small Intestinal Goblet Cell | bed | snATAC-Seq | small intestine goblet cell | |
| Colon Epithelial Cell 2 | bed | snATAC-Seq | colon epithelial cell | |
| Colon Epithelial Cell 3 | bed | snATAC-Seq | colon epithelial cell | |
| Enterochromaffin Cell | bed | snATAC-Seq | enterochromaffin-like cell | |
| Tuft Cell | bed | snATAC-Seq | intestinal tuft cell | |
| Paneth Cell | bed | snATAC-Seq | paneth cell | |
| **Additional datasets for SNP annotation (Supplementary Figure 5, Supplementary Table 4)** | | | | |
| Coronary artery-1 | fastq | ATAC-Seq | coronary artery | https://www.ncbi.nlm.nih.gov/sra/?term=SRR2378591 |
| Coronary artery-2 | fastq | ATAC-Seq | coronary artery | https://www.ncbi.nlm.nih.gov/sra/?term=SRR2378592 |
| Coronary artery-3 | fastq | ATAC-Seq | coronary artery | https://www.ncbi.nlm.nih.gov/sra/?term=SRR2378593 |
| scATAC | fragments.bed | snATAC-Seq | 25 tissues | https://www.ncbi.nlm.nih.gov/geo/query/acc.cgi?acc=GSE184462 |

**Supplementary Table 15:** details of summary statistics used in to calculate the genetic correlation with SCAD
for continuous traits Neff= sample size of study
for binary traits Neff was calculated as follow : neff= 4/(1/Ncases+1/Nctrls)
* : range.

| Trait | abreveation | Type | Strata | Neff | N Cases | N controls | Consortia / study | study reference | Source of summary statistics |
|---|---|---|---|---|---|---|---|---|---|
| Coronary Artery Disease | CAD | binary | Bothsex, Female and male | 613021 | 181522 | 984168 | CARDIoGRAMplusC4D Consortium (Aragam et al) | https://doi.org/10.1101/2021.05.24.21257377 | shared by authors |
| Myocardial Infarction | MI | binary | bothsex | 126612 | 42561 | 123504 | CARDIoGRAMplusC4D Consortium (Nikpay et al) | PMID: 26343387 | http://www.cardiogramplusc4d.org/data-downloads/ |
| Fibromuscular dysplasia | FMD | binary | Bothsex and Female | 5105 | 1556 | 7100 | Georges et al | PMID: 34654805 | http://ftp.ebi.ac.uk/pub/databases/gwas/summary_statistics/GCST90026001-GCST90027000/GCST90026612 |
| Cervical Artery Dissection | CeAD | binary | Bothsex | 5081 | 1393 | 14416 | Stéphanie Debette et al | PMID: 25420145 | shared by authors |
| Intracranial Aneurysm | IA | binary | Bothsex | [14576 - 24253]* | 7495 | 71934 | Mark K Bakker et al | PMID: 33199917 | shared by authors |
| Subarachnoid Haemorrhage | SAH | binary | Bothsex | [10203 - 17019]* | 5140 | 71934 | Mark K Bakker et al | PMID: 33199917 | shared by authors |
| unruptured Intracranial Aneurysm (uIA) | uIA | binary | Bothsex | [5225 - 7721]* | 2070 | 71934 | Mark K Bakker et al | PMID: 33199917 | shared by authors |
| Any stroke | AS | binary | Bothsex | 147590 | 40585 | 406111 | MEGASTROKE consortium | PMID:29531354 | https://www.megastroke.org/ |
| Any Ischemic Stroke | AIS | binary | Bothsex | 126232 | 34217 | 406111 | MEGASTROKE consortium | PMID:29531354 | https://www.megastroke.org/ |
| Large Artery Stroke | LAS | binary | Bothsex | 16985 | 4373 | 146392 | MEGASTROKE consortium | PMID:29531354 | https://www.megastroke.org/ |
| Cardioembolic Stroke | CES | binary | Bothsex | 27795 | 7193 | 204570 | MEGASTROKE consortium | PMID:29531354 | https://www.megastroke.org/ |
| Small Vessel Stroke | SVS | binary | Bothsex | 20958 | 5386 | 192662 | MEGASTROKE consortium | PMID:29531354 | https://www.megastroke.org/ |
| Migraine | MIG | binary | Bothsex, Female and male | 41332 | 10647 | 350494 | UK BioBank Neale lab summary statistics | - | http://www.nealelab.is/uk-biobank |
| Type 2 diabetes | T2D | binary | Bothsex | 272026 | 74124 | 824006 | Mahajan et al | PMID: 30297969 | https://diagram-consortium.org/downloads.html |
| Smoking (ever) | SMK | binary | Bothsex, Female and male | 357132 | 164638 | 195068 | UK BioBank Neale lab summary statistics | - | http://www.nealelab.is/uk-biobank |
| Systolic blood pressure | SBP | continuous | Bothsex, Female and male | 340159 | - | - | UK BioBank Neale lab summary statistics | - | http://www.nealelab.is/uk-biobank |
| Diastolic blood pressure | DBP | continuous | Bothsex, Female and male | 340162 | - | - | UK BioBank Neale lab summary statistics | - | http://www.nealelab.is/uk-biobank |
| Systolic blood pressure | SBP | continuous | Bothsex | [637578 - 745820]* | - | - | Evangelou et al | PMID: 30224653 | ftp://ftp.ebi.ac.uk/pub/databases/gwas/summary_statistics/EvangelouE_30224653_GCST006624 |
| Diastolic blood pressure | DBP | continuous | Bothsex | [647307 - 757601]* | - | - | Evangelou et al | PMID: 30224653 | ftp://ftp.ebi.ac.uk/pub/databases/gwas/summary_statistics/EvangelouE_30224653_GCST006630 |
| Body mass index | BMI | continuous | Bothsex, Female and male | 359983 | - | - | UK BioBank Neale lab summary statistics | - | http://www.nealelab.is/uk-biobank |
| High density | HDL | continuous | Bothsex, Female and male | 315133 | - | - | UK BioBank Neale lab summary statistics | - | http://www.nealelab.is/uk-biobank |
| Low density | LDL | continuous | Bothsex, Female and male | 343621 | - | - | UK BioBank Neale lab summary statistics | - | http://www.nealelab.is/uk-biobank |
| Total cholesterol | TC | continuous | Bothsex, Female and male | 344278 | - | - | UK BioBank Neale lab summary statistics | - | http://www.nealelab.is/uk-biobank |
| Trygliceryde | TG | continuous | Bothsex, Female and male | 343992 | - | - | UK BioBank Neale lab summary statistics | - | http://www.nealelab.is/uk-biobank |
| Hemoglobin | HGB | continuous | Bothsex | 408112 | - | - | Vuckovic et al | PMID: 32888494 | http://ftp.ebi.ac.uk/pub/databases/gwas/summary_statistics/GCST90002001-GCST90003000/GCST90002384/ |
| Hematocrit | HCT | continuous | Bothsex | 408112 | - | - | Vuckovic et al | PMID: 32888494 | http://ftp.ebi.ac.uk/pub/databases/gwas/summary_statistics/GCST90002001-GCST90003000/GCST90002383/ |

| | | | | | | | | | |
|---|---|---|---|---|---|---|---|---|---|
| Neutrophil count | NEU | continuous | Bothsex | 408112 | - | - | Vuckovic et al | PMID: 32888494 | http://ftp.ebi.ac.uk/pub/databases/gwas/summary_statistics/GCST90002001-GCST90003000/GCST90002398 |
| Monocyte count | MONO | continuous | Bothsex | 408112 | - | - | Vuckovic et al | PMID: 32888494 | http://ftp.ebi.ac.uk/pub/databases/gwas/summary_statistics/GCST90002001-GCST90003000/GCST90002393 |
| Lymphocyte counts | LYMPHO | continuous | Bothsex | 408112 | - | - | Vuckovic et al | PMID: 32888494 | http://ftp.ebi.ac.uk/pub/databases/gwas/summary_statistics/GCST90002001-GCST90003000/GCST90002388 |
| Platelet count | PLT | continuous | Bothsex | 408112 | - | - | Vuckovic et al | PMID: 32888494 | http://ftp.ebi.ac.uk/pub/databases/gwas/summary_statistics/GCST90002001-GCST90003000/GCST90002402 |

#### Consortium Authors

#### DISCO Investigators

Nicolas COMBARET[1,] MD and the DISCO Investigators : Pascal MOTREFF[1], MD, PhD ; Géraud SOUTEYRAND[1], MD ; Edouard GERBAUD[2], MD, PhD ; François DERIMAY[3], MD ; Sara BOUAJILA[4], MD; Stéphane MANZO-SILBERMAN[4], MD ; Grégoire RANGE[5], MD ; Nicolas MENEVEAU[6], MD, PhD ; Brahim HARBAOUI[7], MD, PhD ; Benoit LATTUCA[8], MD, PhD ; Didier BRESSON[9], MD ; Lionel MANGIN[10], MD ; Thibault LHERMUSIER[11], MD ; Emmanuel BOIFFARD[12]MD ; Emmanuelle FILIPPI[13], MD ; Vincent ROULE[14], MD, PhD; Jean-Louis GEORGES[15], MD ; Arnaud FLUTTAZ[16], MD ; Stéphanie MARLIERE[17]

1: Department of cardiology, CHU Clermont-Ferrand, CNRS, Université Clermont Auvergne, Clermont-Ferrand, France
2: Cardiology Intensive Care Unit and Interventional Cardiology, Hôpital Cardiologique du Haut Lévêque, CHU de Bordeaux, 5 Avenue de Magellan, 33604 Pessac, France. Bordeaux Cardio-Thoracic Research Centre, U1045, Bordeaux University, Hôpital Xavier Arnozan, Avenue du Haut Lévêque, 33600 Pessac, France
3: Department of Interventional Cardiology, Cardiovascular Hospital and Claude-Bernard University, INSERM Unit 1060 CARMEN, Lyon, France.
4: Department of Cardiology, Hôpital Lariboisière, Assistance Publique des Hôpitaux de Paris, Paris, France.
5: Service de cardiologie, Les Hôpitaux de Chartres, Le Coudray, France
6: Besancon University Hospital, EA3920, University of Burgundy Franche-Comté, Besancon, France.
7: Service de cardiologie, hôpital Croix-Rousse et hôpital Lyon Sud, hospices civils de Lyon, Lyon, France; Université Lyon 1, CREATIS UMR5220, Inserm U1044, INSA-15, Lyon, France.
8: Department of Cardiology, Nimes University Hospital, Montpellier University, Nimes, France
9: Department of Cardiology, Intensive Coronary Care Unit, Emile Muller Hospital, Mulhouse, France.
10: Department of cardiology, centre hospitalier Annecy-Genevois, 74370 Metz-Tessy, France
11: Department of Cardiology, University Hospital of Rangueil, Toulouse, France
12: Department of Cardiology, Centre Hospitalier Départemental (CHD) de Vendée ,La Roche sur Yon, France.
13: Department of Cardiology, General Hospital of Atlantic Brittany, Vannes, France.
14: Department of Cardiology, Caen University Hospital, Caen, France.
15 : Department of cardiology, Centre Hospitalier de Versailles, Le Chesnay-Rocquencourt, France.
16 : Department of Cardiology, Centre Hospitalier Metropole Savoie, Chambery, France.
17 : Department of Cardiology, Grenoble University Hospital, Grenoble, France.

**International Stroke Genetics Consortium (ISGC) Intracranial Aneurysm Working Group:**
Mark K. Bakker
Department of Neurology and Neurosurgery, University Medical Center Utrecht Brain Center, Utrecht University, Utrecht, The Netherlands.

Philippe Bijlenga
Neurosurgery Division, Department of Clinical Neurosciences, Faculty of Medicine, Geneva University Hospitals, Geneva, Switzerland

Romain Bourcier
Université de Nantes, CHU Nantes, INSERM, CNRS, l'institut du thorax, Nantes, France.
CHU Nantes, Department of Neuroradiology, Nantes, France.

Joseph P. Broderick
University of Cincinnati College of Medicine, Cincinnati, OH, USA.

Mikael Fraunberg
Neurosurgery NeuroCenter, Kuopio University Hospital, Kuopio, Finland
Institute of Clinical Medicine, Faculty of Health Sciences, University of Eastern Finland, Kuopio, Finland.

Emilia Gaal–Paavola
Department of Neurosurgery, Helsinki University Hospital, University of Helsinki, Helsinki, Finland and Clinical Neurosciences, University of Helsinki, Helsinki, Finland.

Isabel C. Hostettler
Department of Neurosurgery, Kantonspital St. Gallen, Rorschacher Strasse 95, 9007, St. Gallen, Switzerland.

Juha E. Jaaskelainen
Neurosurgery NeuroCenter, Kuopio University Hospital, Kuopio, Finland. And Institute of Clinical Medicine, Faculty of Health Sciences, University of Eastern Finland, Kuopio, Finland.

Yoichiro Kamatani
Graduate School of Frontier Sciences, The University of Tokyo, Tokyo, Japan.

Antti Lindgren
Neurosurgery NeuroCenter, Kuopio University Hospital, Kuopio, Finland.
Institute of Clinical Medicine, Faculty of Health Sciences, University of Eastern Finland, Kuopio, Finland.

Sandrine Morel
Neurosurgery Division, Department of Clinical Neurosciences, Faculty of Medicine, Geneva University Hospitals, Geneva, Switzerland and Department of Pathology and Immunology, Faculty of Medicine, University of Geneva, Geneva, Switzerland.

Mika Niemela
Department of Neurosurgery, Helsinki University Hospital, University of Helsinki, Helsinki, Finland.

Richard Redon
l'institut du thorax Université de Nantes, CHU Nantes, INSERM, CNRS, Nantes, France.

Gabriel J.E. Rinkel

Department of Neurology and Neurosurgery, University Medical Center Utrecht Brain Center, Utrecht University, Utrecht, The Netherlands.

Guy A Rouleau
Montréal Neurological Institute and Hospital, McGill University, Montréal, QC, Canada.

Robin G. Walters
Clinical Trial Service Unit and Epidemiological Studies Unit, Nuffield Department of Population Health, University of Oxford, Oxford, U.K.
Medical Research Council Population Health Research Unit, University of Oxford, Oxford, U.K.

Bradford B. Worrall
Departments of Neurology and Public Health Sciences, University of Virginia School of Medicine, Charlottesville, VA, USA.

Joanna Pera
Department of Neurology, Faculty of Medicine, Jagiellonian University Medical College, ul. Botaniczna 3, 31-503, Krakow, Poland.

Ynte M. Ruigrok
Department of Neurology and Neurosurgery, University Medical Center Utrecht Brain Center, Utrecht University, Utrecht, The Netherlands.

Agnieszka Slowik
Department of Neurology, Faculty of Medicine, Jagiellonian University Medical College, ul. Botaniczna 3, 31-503, Krakow, Poland.

David J. Werring
Stroke Research Centre, University College London Queen Square Institute of Neurology, London, UK.

Bendik S Winsvold
Department of Research, Innovation and Education, Division of Clinical Neuroscience, Oslo University Hospital, Oslo, Norway.
K. G. Jebsen Center for Genetic Epidemiology, Department of Public Health and Nursing, Faculty of Medicine and Health Sciences, Norwegian University of Science and Technology, Trondheim, Norway.

Daniel Woo
University of Cincinnati College of Medicine, Cincinnati, OH, USA.

### MEGASTROKE CONSORTIUM

Rainer Malik [1], Ganesh Chauhan [2], Matthew Traylor [3], Muralidharan Sargurupremraj [4,5], Yukinori Okada [6,7,8], Aniket Mishra [4,5], Loes Rutten-Jacobs [3], Anne-Katrin Giese [9], Sander W van der Laan [10], Solveig Gretarsdottir [11], Christopher D Anderson [12,13,14,14], Michael Chong [15], Hieab HH Adams [16,17], Tetsuro Ago [18], Peter Almgren [19], Philippe Amouyel [20,21], Hakan Ay [22,13], Traci M Bartz [23], Oscar R Benavente [24], Steve Bevan [25], Giorgio B Boncoraglio [26], Robert D Brown, Jr. [27], Adam S Butterworth [28,29], Caty Carrera [30,31], Cara L Carty [32,33], Daniel I Chasman [34,35], Wei-Min Chen [36], John W Cole [37], Adolfo Correa [38], Ioana Cotlarciuc [39], Carlos Cruchaga [40,41], John Danesh [28,42,43,44], Paul IW de Bakker [45,46], Anita L DeStefano [47,48], Marcel den Hoed [49], Qing Duan [50], Stefan T Engelter [51,52], Guido J Falcone [53,54], Rebecca F Gottesman [55], Raji P Grewal [56], Vilmundur Gudnason [57,58], Stefan Gustafsson [59], Jeffrey Haessler [60], Tamara B Harris [61], Ahamad Hassan [62], Aki S Havulinna [63,64], Susan R Heckbert [65], Elizabeth G Holliday [66,67], George Howard [68], Fang-Chi Hsu [69], Hyacinth I Hyacinth [70], M Arfan Ikram [16], Erik Ingelsson [71,72], Marguerite R Irvin [73], Xueqiu Jian [74], Jordi Jiménez-Conde [75], Julie A Johnson [76,77], J Wouter Jukema [78], Masahiro Kanai [6,7,79], Keith L Keene [80,81], Brett M Kissela [82], Dawn O Kleindorfer [82], Charles Kooperberg [60], Michiaki Kubo [83], Leslie A Lange [84], Carl D Langefeld [85], Claudia Langenberg [86], Lenore J Launer [87], Jin-Moo Lee [88], Robin Lemmens [89,90], Didier Leys [91], Cathryn M Lewis [92,93], Wei-Yu Lin [28,94], Arne G Lindgren [95,96], Erik Lorentzen [97], Patrik K Magnusson [98], Jane Maguire [99], Ani Manichaikul [36], Patrick F McArdle [100], James F Meschia [101], Braxton D Mitchell [100,102], Thomas H Mosley [103,104], Michael A Nalls [105,106], Toshiharu Ninomiya [107], Martin J O'Donnell [15,108], Bruce M Psaty [109,110,111,112], Sara L Pulit [113,45], Kristiina Rannikmäe [114,115], Alexander P Reiner [65,116], Kathryn M Rexrode [117], Kenneth Rice [118], Stephen S Rich [36], Paul M Ridker [34,35], Natalia S Rost [9,13], Peter M Rothwell [119], Jerome I Rotter [120,121], Tatjana Rundek [122], Ralph L Sacco [122], Saori Sakaue [7,123], Michele M Sale [124], Veikko Salomaa [63], Bishwa R Sapkota [125], Reinhold Schmidt [126], Carsten O Schmidt [127], Ulf Schminke [128], Pankaj Sharma [39], Agnieszka Slowik [129], Cathie LM Sudlow [114,115], Christian Tanislav [130], Turgut Tatlisumak [131,132], Kent D Taylor [120,121], Vincent NS Thijs [133,134], Gudmar Thorleifsson [11], Unnur Thorsteinsdottir [11], Steffen Tiedt [1], Stella Trompet [135], Christophe Tzourio [5,136,137], Cornelia M van Duijn [138,139], Matthew Walters [140], Nicholas J Wareham [86], Sylvia Wassertheil-Smoller [141], James G Wilson [142], Kerri L Wiggins [109], Qiong Yang [47], Salim Yusuf [15], Najaf Amin [16], Hugo S Aparicio [185,48], Donna K Arnett [186], John Attia [187], Alexa S Beiser [47,48], Claudine Berr [188], Julie E Buring [34,35], Mariana Bustamante [189], Valeria Caso [190], Yu-Ching Cheng [191], Seung Hoan Choi [192,48], Ayesha Chowhan [185,48], Natalia Cullell [31], Jean-François Dartigues [193,194], Hossein Delavaran [95,96], Pilar Delgado [195], Marcus Dörr [196,197], Gunnar Engström [19], Ian Ford [198], Wander S Gurpreet [199], Anders Hamsten [200,201], Laura Heitsch [202], Atsushi Hozawa [203], Laura Ibanez [204], Andreea Ilinca [95,96], Martin Ingelsson [205], Motoki Iwasaki [206], Rebecca D Jackson [207], Katarina Jood [208], Pekka Jousilahti [63], Sara Kaffashian [4,5], Lalit Kalra [209], Masahiro Kamouchi [210], Takanari Kitazono [211], Olafur Kjartansson [212], Manja Kloss [213], Peter J Koudstaal [214], Jerzy Krupinski [215], Daniel L Labovitz [216], Cathy C Laurie [118],

Christopher R Levi [217], Linxin Li [218], Lars Lind [219], Cecilia M Lindgren [220,221], Vasileios Lioutas [222,48], Yong Mei Liu [223], Oscar L Lopez [224], Hirata Makoto [225], Nicolas Martinez-Majander [172], Koichi Matsuda [225], Naoko Minegishi [203], Joan Montaner [226], Andrew P Morris [227,228], Elena Muiño [31], Martina Müller-Nurasyid [229,230,231], Bo Norrving [95,96], Soichi Ogishima [203], Eugenio A Parati [232], Leema Reddy Peddareddygari [56], Nancy L Pedersen [98,233], Joanna Pera [129], Markus Perola [63,234], Alessandro Pezzini [235], Silvana Pileggi [236], Raquel Rabionet [237], Iolanda Riba-Llena [30], Marta Ribasés [238], Jose R Romero [185,48], Jaume Roquer [239,240], Anthony G Rudd [241,242], Antti-Pekka Sarin [243,244], Ralhan Sarju [199], Chloe Sarnowski [47,48], Makoto Sasaki [245], Claudia L Satizabal [185,48], Mamoru Satoh [245], Naveed Sattar [246], Norie Sawada [206], Gerli Sibolt [172], Ásgeir Sigurdsson [247], Albert Smith [248], Kenji Sobue [245], Carolina Soriano-Tárraga [240], Tara Stanne [249], O Colin Stine [250], David J Stott [251], Konstantin Strauch [229,252], Takako Takai [203], Hideo Tanaka [253,254], Kozo Tanno [245], Alexander Teumer [255], Liisa Tomppo [172], Nuria P Torres-Aguila [31], Emmanuel Touze [256,257], Shoichiro Tsugane [206], Andre G Uitterlinden [258], Einar M Valdimarsson [259], Sven J van der Lee [16], Henry Völzke [255], Kenji Wakai [253], David Weir [260], Stephen R Williams [261], Charles DA Wolfe [241,242], Quenna Wong [118], Huichun Xu [191], Taiki Yamaji [206], Dharambir K Sanghera [125,169,170], Olle Melander [19], Christina Jern [171], Daniel Strbian [172,173], Israel Fernandez-Cadenas [31,30], W T Longstreth, Jr [174,65], Arndt Rolfs [175], Jun Hata [107], Daniel Woo [82], Jonathan Rosand [12,13,14], Guillaume Pare [15], Jemma C Hopewell [176], Danish Saleheen [177], Kari Stefansson [11,178], Bradford B Worrall [179], Steven J Kittner [37], Sudha Seshadri [180,48], Myriam Fornage [74,181], Hugh S Markus [3], Joanna MM Howson [28], Yoichiro Kamatani [6,182], Stephanie Debette [4,5], Martin Dichgans [1,183,184]

1 Institute for Stroke and Dementia Research (ISD), University Hospital, LMU Munich, Munich, Germany

2 Centre for Brain Research, Indian Institute of Science, Bangalore, India

3 Stroke Research Group, Division of Clinical Neurosciences, University of Cambridge, UK

4 INSERM U1219 Bordeaux Population Health Research Center, Bordeaux, France

5 University of Bordeaux, Bordeaux, France

6 Laboratory for Statistical Analysis, RIKEN Center for Integrative Medical Sciences, Yokohama, Japan

7 Department of Statistical Genetics, Osaka University Graduate School of Medicine, Osaka, Japan

8 Laboratory of Statistical Immunology, Immunology Frontier Research Center (WPI-IFReC), Osaka University, Suita, Japan.

9 Department of Neurology, Massachusetts General Hospital, Harvard Medical School, Boston, MA, USA

10 Laboratory of Experimental Cardiology, Division of Heart and Lungs, University Medical Center Utrecht, University of Utrecht, Utrecht,Netherlands

11 deCODE genetics/AMGEN inc, Reykjavik, Iceland

12 Center for Genomic Medicine, Massachusetts General Hospital (MGH), Boston, MA, USA

13 J. Philip Kistler Stroke Research Center, Department of Neurology, MGH, Boston, MA, USA
14 Program in Medical and Population Genetics, Broad Institute, Cambridge, MA, USA
15 Population Health Research Institute, McMaster University, Hamilton, Canada
16 Department of Epidemiology, Erasmus University Medical Center, Rotterdam, Netherlands
17 Department of Radiology and Nuclear Medicine, Erasmus University Medical Center, Rotterdam, Netherlands
18 Department of Medicine and Clinical Science, Graduate School of Medical Sciences, Kyushu University, Fukuoka, Japan
19 Department of Clinical Sciences, Lund University, Malmö, Sweden
20 Univ. Lille, Inserm, Institut Pasteur de Lille, LabEx DISTALZ-UMR1167, Risk factors and molecular determinants of aging-related diseases, F-59000 Lille, France
21 Centre Hosp. Univ Lille, Epidemiology and Public Health Department, F-59000 Lille, France
22 AA Martinos Center for Biomedical Imaging, Department of Radiology, Massachusetts General Hospital, Harvard Medical School, Boston, MA, USA
23 Cardiovascular Health Research Unit, Departments of Biostatistics and Medicine, University of Washington, Seattle, WA, USA
24 Division of Neurology, Faculty of Medicine, Brain Research Center, University of British Columbia, Vancouver, Canada
25 School of Life Science, University of Lincoln, Lincoln, UK
26 Department of Cerebrovascular Diseases, Fondazione IRCCS Istituto Neurologico "Carlo Besta", Milano, Italy
27 Department of Neurology, Mayo Clinic Rochester, Rochester, MN, USA
28 MRC/BHF Cardiovascular Epidemiology Unit, Department of Public Health and Primary Care, University of Cambridge, Cambridge, UK
29 The National Institute for Health Research Blood and Transplant Research Unit in Donor Health and Genomics, University of Cambridge, UK
30 Neurovascular Research Laboratory, Vall d'Hebron Institut of Research, Neurology and Medicine Departments-Universitat Autònoma de Barcelona, Vall d’Hebrón Hospital, Barcelona, Spain
31 Stroke Pharmacogenomics and Genetics, Fundacio Docència i Recerca MutuaTerrassa, Terrassa, Spain
32 Children's Research Institute, Children's National Medical Center, Washington, DC, USA
33 Center for Translational Science, George Washington University, Washington, DC, USA
34 Division of Preventive Medicine, Brigham and Women's Hospital, Boston, MA, USA
35 Harvard Medical School, Boston, MA, USA
36 Center for Public Health Genomics, Department of Public Health Sciences, University of Virginia, Charlottesville, VA, USA
37 Department of Neurology, University of Maryland School of Medicine and Baltimore VAMC, Baltimore, MD, USA

38 Departments of Medicine, Pediatrics and Population Health Science, University of Mississippi Medical Center, Jackson, MS, USA
39 Institute of Cardiovascular Research, Royal Holloway University of London, UK & Ashford and St Peters Hospital, Surrey UK
40 Department of Psychiatry,The Hope Center Program on Protein Aggregation and Neurodegeneration (HPAN),Washington University, School of Medicine, St. Louis, MO, USA
41 Department of Developmental Biology, Washington University School of Medicine, St. Louis, MO, USA
42 NIHR Blood and Transplant Research Unit in Donor Health and Genomics, Department of Public Health and Primary Care, University of Cambridge, Cambridge, UK
43 Wellcome Trust Sanger Institute, Wellcome Trust Genome Campus, Hinxton, Cambridge, UK
44 British Heart Foundation, Cambridge Centre of Excellence, Department of Medicine, University of Cambridge, Cambridge, UK
45 Department of Medical Genetics, University Medical Center Utrecht, Utrecht, Netherlands
46 Department of Epidemiology, Julius Center for Health Sciences and Primary Care, University Medical Center Utrecht, Utrecht, Netherlands
47 Boston University School of Public Health, Boston, MA, USA
48 Framingham Heart Study, Framingham, MA, USA
49 Department of Immunology, Genetics and Pathology and Science for Life Laboratory, Uppsala University, Uppsala, Sweden
50 Department of Genetics, University of North Carolina, Chapel Hill, NC, USA
51 Department of Neurology and Stroke Center, Basel University Hospital, Switzerland
52 Neurorehabilitation Unit, University and University Center for Medicine of Aging and Rehabilitation Basel, Felix Platter Hospital, Basel, Switzerland
53 Department of Neurology, Yale University School of Medicine, New Haven, CT, USA
54 Program in Medical and Population Genetics, The Broad Institute of Harvard and MIT, Cambridge, MA, USA
55 Department of Neurology, Johns Hopkins University School of Medicine, Baltimore, MD, USA
56 Neuroscience Institute, SF Medical Center, Trenton, NJ, USA
57 Icelandic Heart Association Research Institute, Kopavogur, Iceland
58 University of Iceland, Faculty of Medicine, Reykjavik, Iceland
59 Department of Medical Sciences, Molecular Epidemiology and Science for Life Laboratory, Uppsala University, Uppsala, Sweden
60 Division of Public Health Sciences, Fred Hutchinson Cancer Research Center, Seattle, WA, USA
61 Laboratory of Epidemiology and Population Science, National Institute on Aging, National Institutes of Health, Bethesda, MD, USA
62 Department of Neurology, Leeds General Infirmary, Leeds Teaching Hospitals NHS Trust, Leeds, UK

63 National Institute for Health and Welfare, Helsinki, Finland
64 FIMM - Institute for Molecular Medicine Finland, Helsinki, Finland
65 Department of Epidemiology, University of Washington, Seattle, WA, USA
66 Public Health Stream, Hunter Medical Research Institute, New Lambton, Australia
67 Faculty of Health and Medicine, University of Newcastle, Newcastle, Australia
68 School of Public Health, University of Alabama at Birmingham, Birmingham, AL, USA
69 Department of Biostatistical Sciences, Wake Forest School of Medicine, Winston-Salem, NC, USA
70 Aflac Cancer and Blood Disorder Center, Department of Pediatrics, Emory University School of Medicine, Atlanta, GA, USA
71 Department of Medicine, Division of Cardiovascular Medicine, Stanford University School of Medicine, CA, USA
72 Department of Medical Sciences, Molecular Epidemiology and Science for Life Laboratory, Uppsala University, Uppsala, Sweden
73 Epidemiology, School of Public Health, University of Alabama at Birmingham, USA
74 Brown Foundation Institute of Molecular Medicine, University of Texas Health Science Center at Houston, Houston, TX, USA
75 Neurovascular Research Group (NEUVAS), Neurology Department, Institut Hospital del Mar d'Investigació Mèdica, Universitat Autònoma de Barcelona, Barcelona, Spain
76 Department of Pharmacotherapy and Translational Research and Center for Pharmacogenomics, University of Florida, College of Pharmacy, Gainesville, FL, USA
77 Division of Cardiovascular Medicine, College of Medicine, University of Florida, Gainesville, FL, USA
78 Department of Cardiology, Leiden University Medical Center, Leiden, the Netherlands
79 Program in Bioinformatics and Integrative Genomics, Harvard Medical School, Boston, MA, USA
80 Department of Biology, East Carolina University, Greenville, NC, USA
81 Center for Health Disparities, East Carolina University, Greenville, NC, USA
82 University of Cincinnati College of Medicine, Cincinnati, OH, USA
83 RIKEN Center for Integrative Medical Sciences, Yokohama, Japan
84 Department of Medicine, University of Colorado Denver, Anschutz Medical Campus, Aurora, CO, USA
85 Center for Public Health Genomics and Department of Biostatistical Sciences, Wake Forest School of Medicine, Winston-Salem, NC, USA
86 MRC Epidemiology Unit, University of Cambridge School of Clinical Medicine, Institute of Metabolic Science, Cambridge Biomedical Campus, Cambridge, UK
87 Intramural Research Program, National Institute on Aging, National Institutes of Health, Bethesda, MD, USA

88 Department of Neurology, Radiology, and Biomedical Engineering, Washington University School of Medicine, St. Louis, MO, USA
89 KU Leuven – University of Leuven, Department of Neurosciences, Experimental Neurology, Leuven, Belgium
90 VIB Center for Brain & Disease Research, University Hospitals Leuven, Department of Neurology, Leuven, Belgium
91 Univ.-Lille, INSERM U 1171. CHU Lille. Lille, France
92 Department of Medical and Molecular Genetics, King's College London, London, UK
93 SGDP Centre, Institute of Psychiatry, Psychology & Neuroscience, King's College London, London, UK
94 Northern Institute for Cancer Research, Paul O'Gorman Building, Newcastle University, Newcastle, UK
95 Department of Clinical Sciences Lund, Neurology, Lund University, Lund, Sweden
96 Department of Neurology and Rehabilitation Medicine, Skåne University Hospital, Lund, Sweden
97 Bioinformatics Core Facility, University of Gothenburg, Gothenburg, Sweden
98 Department of Medical Epidemiology and Biostatistics, Karolinska Institutet, Stockholm, Sweden
99 University of Technology Sydney, Faculty of Health, Ultimo, Australia
100 Department of Medicine, University of Maryland School of Medicine, MD, USA
101 Department of Neurology, Mayo Clinic, Jacksonville, FL, USA
102 Geriatrics Research and Education Clinical Center, Baltimore Veterans Administration Medical Center, Baltimore, MD, USA
103 Division of Geriatrics, School of Medicine, University of Mississippi Medical Center, Jackson, MS, USA
104 Memory Impairment and Neurodegenerative Dementia Center, University of Mississippi Medical Center, Jackson, MS, USA
105 Laboratory of Neurogenetics, National Institute on Aging, National institutes of Health, Bethesda, MD, USA
106 Data Tecnica International, Glen Echo MD, USA
107 Department of Epidemiology and Public Health, Graduate School of Medical Sciences, Kyushu University, Fukuoka, Japan
108 Clinical Research Facility, Department of Medicine, NUI Galway, Galway, Ireland
109 Cardiovascular Health Research Unit, Department of Medicine, University of Washington, Seattle, WA, USA
110 Department of Epidemiology, University of Washington, Seattle, WA
111 Department of Health Services, University of Washington, Seattle, WA, USA
112 Kaiser Permanente Washington Health Research Institute, Seattle, WA, USA

113 Brain Center Rudolf Magnus, Department of Neurology, University Medical Center Utrecht, Utrecht, The Netherlands
114 Usher Institute of Population Health Sciences and Informatics, University of Edinburgh, Edinburgh, UK
115 Centre for Clinical Brain Sciences, University of Edinburgh, Edinburgh, UK
116 Fred Hutchinson Cancer Research Center, University of Washington, Seattle, WA, USA
117 Department of Medicine, Brigham and Women's Hospital, Boston, MA, USA
118 Department of Biostatistics, University of Washington, Seattle, WA, USA
119 Nuffield Department of Clinical Neurosciences, University of Oxford, UK
120 Institute for Translational Genomics and Population Sciences, Los Angeles Biomedical Research Institute at Harbor-UCLA Medical Center, Torrance, CA, USA
121 Division of Genomic Outcomes, Department of Pediatrics, Harbor-UCLA Medical Center, Torrance, CA, USA
122 Department of Neurology, Miller School of Medicine, University of Miami, Miami, FL, USA
123 Department of Allergy and Rheumatology, Graduate School of Medicine, the University of Tokyo, Tokyo, Japan
124 Center for Public Health Genomics, University of Virginia, Charlottesville, VA, USA
125 Department of Pediatrics, College of Medicine, University of Oklahoma Health Sciences Center, Oklahoma City, OK, USA
126 Department of Neurology, Medical University of Graz, Graz, Austria
127 University Medicine Greifswald, Institute for Community Medicine, SHIP-KEF, Greifswald, Germany
128 University Medicine Greifswald, Department of Neurology, Greifswald, Germany
129 Department of Neurology, Jagiellonian University, Krakow, Poland
130 Department of Neurology, Justus Liebig University, Giessen, Germany
131 Department of Clinical Neurosciences/Neurology, Institute of Neuroscience and Physiology, Sahlgrenska Academy at University of Gothenburg, Gothenburg, Sweden
132 Sahlgrenska University Hospital, Gothenburg, Sweden
133 Stroke Division, Florey Institute of Neuroscience and Mental Health, University of Melbourne, Heidelberg, Australia
134 Austin Health, Department of Neurology, Heidelberg, Australia
135 Department of Internal Medicine, Section Gerontology and Geriatrics, Leiden University Medical Center, Leiden, the Netherlands
136 INSERM U1219, Bordeaux, France
137 Department of Public Health, Bordeaux University Hospital, Bordeaux, France
138 Genetic Epidemiology Unit, Department of Epidemiology, Erasmus University Medical Center Rotterdam, Netherlands
139 Center for Medical Systems Biology, Leiden, Netherlands

140 School of Medicine, Dentistry and Nursing at the University of Glasgow, Glasgow, UK
141 Department of Epidemiology and Population Health, Albert Einstein College of Medicine, NY, USA
142 Department of Physiology and Biophysics, University of Mississippi Medical Center, Jackson, MS, USA
143 A full list of members and affiliations appears in the Supplementary Note
144 Department of Human Genetics, McGill University, Montreal, Canada
145 Department of Pathophysiology, Institute of Biomedicine and Translation Medicine, University of Tartu, Tartu, Estonia
146 Department of Cardiac Surgery, Tartu University Hospital, Tartu, Estonia
147 Clinical Gene Networks AB,Stockholm, Sweden
148 Department of Genetics and Genomic Sciences, The Icahn Institute for Genomics and Multiscale Biology Icahn School of Medicine at Mount Sinai, New York, NY , USA
149 Department of Pathophysiology, Institute of Biomedicine and Translation Medicine, University of Tartu, Biomeedikum, Tartu, Estonia
150 Integrated Cardio Metabolic Centre, Department of Medicine, Karolinska Institutet, Karolinska Universitetssjukhuset, Huddinge, Sweden.
151 Clinical Gene Networks AB, Stockholm, Sweden
152 Sorbonne Universités, UPMC Univ. Paris 06, INSERM, UMR_S 1166, Team Genomics & Pathophysiology of Cardiovascular Diseases, Paris, France
153 ICAN Institute for Cardiometabolism and Nutrition, Paris, France
154 Department of Biomedical Engineering, University of Virginia, Charlottesville, VA, USA
155 Group Health Research Institute, Group Health Cooperative, Seattle, WA, USA
156 Seattle Epidemiologic Research and Information Center, VA Office of Research and Development, Seattle, WA, USA
157 Cardiovascular Research Center, Massachusetts General Hospital, Boston, MA, USA
158 Department of Medical Research, Bærum Hospital, Vestre Viken Hospital Trust, Gjettum, Norway
159 Saw Swee Hock School of Public Health, National University of Singapore and National University Health System, Singapore
160 National Heart and Lung Institute, Imperial College London, London, UK
161 Department of Gene Diagnostics and Therapeutics, Research Institute, National Center for Global Health and Medicine, Tokyo, Japan
162 Department of Epidemiology, Tulane University School of Public Health and Tropical Medicine, New Orleans, LA, USA
163 Department of Cardiology,University Medical Center Groningen, University of Groningen, Netherlands

164 MRC-PHE Centre for Environment and Health, School of Public Health, Department of Epidemiology and Biostatistics, Imperial College London, London, UK
165 Department of Epidemiology and Biostatistics, Imperial College London, London, UK
166 Department of Cardiology, Ealing Hospital NHS Trust, Southall, UK
167 National Heart, Lung and Blood Research Institute, Division of Intramural Research, Population Sciences Branch, Framingham, MA, USA
168 A full list of members and affiliations appears at the end of the manuscript
169 Department of Phamaceutical Sciences, Collge of Pharmacy, University of Oklahoma Health Sciences Center, Oklahoma City, OK, USA
170 Oklahoma Center for Neuroscience, Oklahoma City, OK, USA
171 Department of Pathology and Genetics, Institute of Biomedicine, The Sahlgrenska Academy at University of Gothenburg, Gothenburg, Sweden
172 Department of Neurology, Helsinki University Hospital, Helsinki, Finland
173 Clinical Neurosciences, Neurology, University of Helsinki, Helsinki, Finland
174 Department of Neurology, University of Washington, Seattle, WA, USA
175 Albrecht Kossel Institute, University Clinic of Rostock, Rostock, Germany
176 Clinical Trial Service Unit and Epidemiological Studies Unit, Nuffield Department of Population Health, University of Oxford, Oxford, UK
177 Department of Genetics, Perelman School of Medicine, University of Pennsylvania, PA, USA
178 Faculty of Medicine, University of Iceland, Reykjavik, Iceland
179 Departments of Neurology and Public Health Sciences, University of Virginia School of Medicine, Charlottesville, VA, USA
180 Department of Neurology, Boston University School of Medicine, Boston, MA, USA
181 Human Genetics Center, University of Texas Health Science Center at Houston, Houston, TX, USA
182 Center for Genomic Medicine, Kyoto University Graduate School of Medicine, Kyoto, Japan
183 Munich Cluster for Systems Neurology (SyNergy), Munich, Germany
184 German Center for Neurodegenerative Diseases (DZNE), Munich, Germany
185 Boston University School of Medicine, Boston, MA, USA
186 University of Kentucky College of Public Health, Lexington, KY, USA
187 University of Newcastle and Hunter Medical Research Institute, New Lambton, Australia
188 Univ. Montpellier, Inserm, U1061, Montpellier, France
189 Centre for Research in Environmental Epidemiology, Barcelona, Spain
190 Department of Neurology, Università degli Studi di Perugia, Umbria, Italy
191 Department of Medicine, University of Maryland School of Medicine, Baltimore, MD, USA
192 Broad Institute, Cambridge, MA, USA
193 Univ. Bordeaux, Inserm, Bordeaux Population Health Research Center, UMR 1219, Bordeaux, France

194 Bordeaux University Hospital, Department of Neurology, Memory Clinic, Bordeaux, France
195 Neurovascular Research Laboratory. Vall d'Hebron Institut of Research, Neurology and Medicine Departments-Universitat Autònoma de Barcelona. Vall d'Hebrón Hospital, Barcelona, Spain
196 University Medicine Greifswald, Department of Internal Medicine B, Greifswald, Germany
197 DZHK, Greifswald, Germany
198 Robertson Center for Biostatistics, University of Glasgow, Glasgow, UK
199 Hero DMC Heart Institute, Dayanand Medical College & Hospital, Ludhiana, India
200 Atherosclerosis Research Unit, Department of Medicine Solna, Karolinska Institutet, Stockholm, Sweden
201 Karolinska Institutet, Stockholm, Sweden
202 Division of Emergency Medicine, and Department of Neurology, Washington University School of Medicine, St. Louis, MO, USA
203 Tohoku Medical Megabank Organization, Sendai, Japan
204 Department of Psychiatry, Washington University School of Medicine, St. Louis, MO, USA
205 Department of Public Health and Caring Sciences / Geriatrics, Uppsala University, Uppsala, Sweden
206 Epidemiology and Prevention Group, Center for Public Health Sciences, National Cancer Center, Tokyo, Japan
207 Department of Internal Medicine and the Center for Clinical and Translational Science, The Ohio State University, Columbus, OH, USA
208 Institute of Neuroscience and Physiology, the Sahlgrenska Academy at University of Gothenburg, Goteborg, Sweden
209 Department of Basic and Clinical Neurosciences, King's College London, London, UK
210 Department of Health Care Administration and Management, Graduate School of Medical Sciences, Kyushu University, Japan
211 Department of Medicine and Clinical Science, Graduate School of Medical Sciences, Kyushu University, Japan
212 Landspitali National University Hospital, Departments of Neurology & Radiology, Reykjavik, Iceland
213 Department of Neurology, Heidelberg University Hospital, Germany
214 Department of Neurology, Erasmus University Medical Center
215 Hospital Universitari Mutua Terrassa, Terrassa (Barcelona), Spain
216 Albert Einstein College of Medicine, Montefiore Medical Center, New York, NY, USA
217 John Hunter Hospital, Hunter Medical Research Institute and University of Newcastle, Newcastle, NSW, Australia
218 Centre for Prevention of Stroke and Dementia, Nuffield Department of Clinical Neurosciences, University of Oxford, UK

219 Department of Medical Sciences, Uppsala University, Uppsala, Sweden
220 Genetic and Genomic Epidemiology Unit, Wellcome Trust Centre for Human Genetics, University of Oxford, Oxford, UK
221 The Wellcome Trust Centre for Human Genetics, Oxford, UK
222 Beth Israel Deaconess Medical Center, Boston, MA, USA
223 Wake Forest School of Medicine, Wake Forest, NC, USA
224 Department of Neurology, University of Pittsburgh, Pittsburgh, PA, USA
225 BioBank Japan, Laboratory of Clinical Sequencing, Department of Computational biology and medical Sciences, Graduate school of Frontier Sciences, The University of Tokyo, Tokyo, Japan
226 Neurovascular Research Laboratory, Vall d'Hebron Institut of Research, Neurology and Medicine Departments-Universitat Autònoma de Barcelona. Vall d'Hebrón Hospital, Barcelona, Spain
227 Department of Biostatistics, University of Liverpool, Liverpool, UK
228 Wellcome Trust Centre for Human Genetics, University of Oxford, Oxford, UK
229 Institute of Genetic Epidemiology, Helmholtz Zentrum München - German Research Center for Environmental Health, Neuherberg, Germany
230 Department of Medicine I, Ludwig-Maximilians-Universität, Munich, Germany
231 DZHK (German Centre for Cardiovascular Research), partner site Munich Heart Alliance, Munich, Germany
232 Department of Cerebrovascular Diseases, Fondazione IRCCS Istituto Neurologico "Carlo Besta", Milano, Italy
233 Karolinska Institutet, MEB, Stockholm, Sweden
234 University of Tartu, Estonian Genome Center, Tartu, Estonia, Tartu, Estonia
235 Department of Clinical and Experimental Sciences, Neurology Clinic, University of Brescia, Italy
236 Translational Genomics Unit, Department of Oncology, IRCCS Istituto di Ricerche Farmacologiche Mario Negri, Milano, Italy
237 Department of Genetics, Microbiology and Statistics, University of Barcelona, Barcelona, Spain
238 Psychiatric Genetics Unit, Group of Psychiatry, Mental Health and Addictions, Vall d'Hebron Research Institute (VHIR), Universitat Autònoma de Barcelona, Biomedical Network Research Centre on Mental Health (CIBERSAM), Barcelona, Spain
239 Department of Neurology, IMIM-Hospital del Mar, and Universitat Autònoma de Barcelona, Spain
240 IMIM (Hospital del Mar Medical Research Institute), Barcelona, Spain
241 National Institute for Health Research Comprehensive Biomedical Research Centre, Guy's & St. Thomas' NHS Foundation Trust and King's College London, London, UK
242 Division of Health and Social Care Research, King's College London, London, UK
243 FIMM-Institute for Molecular Medicine Finland, Helsinki, Finland
244 THL-National Institute for Health and Welfare, Helsinki, Finland

245 Iwate Tohoku Medical Megabank Organization, Iwate Medical University, Iwate, Japan
246 BHF Glasgow Cardiovascular Research Centre, Faculty of Medicine, Glasgow, UK
247 deCODE Genetics/Amgen, Inc., Reykjavik, Iceland
248 Icelandic Heart Association, Reykjavik, Iceland
249 Institute of Biomedicine, the Sahlgrenska Academy at University of Gothenburg, Goteborg, Sweden
250 Department of Epidemiology, University of Maryland School of Medicine, Baltimore, MD, USA
251 Institute of Cardiovascular and Medical Sciences, Faculty of Medicine, University of Glasgow, Glasgow, UK
252 Chair of Genetic Epidemiology, IBE, Faculty of Medicine, LMU Munich, Germany
253 Division of Epidemiology and Prevention, Aichi Cancer Center Research Institute, Nagoya, Japan
254 Department of Epidemiology, Nagoya University Graduate School of Medicine, Nagoya, Japan
255 University Medicine Greifswald, Institute for Community Medicine, SHIP-KEF, Greifswald, Germany
256 Department of Neurology, Caen University Hospital, Caen, France
257 University of Caen Normandy, Caen, France
258 Department of Internal Medicine, Erasmus University Medical Center, Rotterdam, Netherlands
259 Landspitali University Hospital, Reykjavik, Iceland
260 Survey Research Center, University of Michigan, Ann Arbor, MI, USA
261 University of Virginia Department of Neurology, Charlottesville, VA, USA

### CARDIoGRAMplusC4D Consortium

Krishna G Aragam[1,2,3,4]*, Tao Jiang[5]*, Anuj Goel[6,7]*, Stavroula Kanoni[8]*, Brooke N Wolford[9]*, Elle M Weeks[4], Minxian Wang[3,4], George Hindy[10], Wei Zhou[4,11,12,9], Christopher Grace[6,7], Carolina Roselli[3], Nicholas A Marston[13], Frederick K Kamanu[13], Ida Surakka[14], Loreto Muñoz Venegas[15,16], Paul Sherliker[17], Satoshi Koyama[18], Kazuyoshi Ishigaki[19], Bjørn O Åsvold[20,21,22], Michael R Brown[23], Ben Brumpton[20,21], Paul S de Vries[23], Olga Giannakopoulou[8], Panagiota Giardoglou[24], Daniel F Gudbjartsson[25,26], Ulrich Güldener[27], Syed M. Ijlal Haider[15], Anna Helgadottir[25], Maysson Ibrahim[28], Adnan Kastrati[27,29], Thorsten Kessler[27,29], Ling Li[27], Lijiang Ma[30,31], Thomas Meitinger[32,33,29], Sören Mucha[15], Matthias Munz[15], Federico Murgia[28], Jonas B Nielsen[34,20], Markus M Nöthen[35], Shichao Pang[27], Tobias Reinberger[15], Gudmar Thorleifsson[25], Moritz von Scheidt[27,29], Jacob K Ulirsch[4,11,36], EPIC-CVD Consortium, Biobank Japan, David O Arnar[25,37,38], Deepak S Atri[39,3], Noël P Burtt[4], Maria C Costanzo[4], Jason Flannick[40], Rajat M Gupta[39,3,4], Kaoru Ito[18], Dong-Keun Jang[4], Yoichiro Kamatani[41], Amit V Khera[2,3,4], Issei Komuro[42], Iftikhar J Kullo[43], Luca A Lotta[44], Christopher P Nelson[45], Robert Roberts[46], Gudmundur Thorgeirsson[25,37,38], Unnur Thorsteinsdottir[25,37], Thomas R Webb[45], Aris Baras[44], Johan LM Björkegren[47,48,49], Eric Boerwinkle[23,50], George Dedoussis[24], Hilma Holm[25], Kristian Hveem[20,21], Olle Melander[51], Alanna C Morrison[23], Marju Orho-Melander[51], Loukianos S Rallidis[52], Arno Ruusalepp[53], Marc S Sabatine[13], Kari Stefansson[25,37], Pierre Zalloua[54,55], Patrick T Ellinor[1,3], Martin Farrall[6,7], John Danesh[5,56,57,58,59,60], Christian T Ruff[13], Hilary K Finucane[4,11,12], Jemma C Hopewell[28], Robert Clarke[28], Jeanette Erdmann[15,61]†, Nilesh J Samani[45]†, Heribert Schunkert[27,29]†, Hugh Watkins[6,7]†, Cristen J Willer[14,9,62]†, Panos Deloukas[8,63]†, Sekar Kathiresan[64]†, Adam S Butterworth[5,56,60,59,57]† on behalf of.

[1]Cardiovascular Research Center, Massachusetts General Hospital, 185 Cambridge St., Boston, MA, 02114, USA

[2]Center for Genomic Medicine, Massachusetts General Hospital, 185 Cambridge St., Boston, MA, 02114, USA

[3]Cardiovascular Disease Initiative, Broad Institute of MIT and Harvard, 75 Ames St., Cambridge, MA, 02142, USA

[4]Program in Medical and Population Genetics, Broad Institute of MIT and Harvard, 75 Ames St., Cambridge, MA, 02142, USA

[5]BHF Cardiovascular Epidemiology Unit, Department of Public Health and Primary Care, University of Cambridge, Worts Causeway, Cambridge, CB1 8RN, UK

[6]Radcliffe Department of Medicine, Division of Cardiovascular Medicine, University of Oxford, Headley Way, Oxford, OX3 9DU, UK

[7]Wellcome Centre for Human Genetics, University of Oxford, Roosevelt Drive, Oxford, OX3 7BN, UK

[8]William Harvey Research Institute, Barts and the London School of Medicine and Dentistry, Queen Mary University of London, Charterhouse square, London, EC1M 6BQ, UK

[9]Department of Computational Medicine & Bioinformatics, University of Michigan, Palmer Ave., Ann Arbor, Michigan, 48109, USA

[10]Department of Population Medicine, Qatar University College of Medicine, Doha, Qatar

[11]Analytic and Translational Genetics Unit, Massachusetts General Hospital, Boston, Massachusetts, 02114, USA

[12]Stanley Center for Psychiatric Research, Broad Institute of MIT and Harvard, Cambridge, Massachusetts, USA

[13]TIMI Study Group, Division of Cardiovascular Medicine, Brigham and Women's Hospital, Harvard Medical School, 60 Fenwood Rd., Boston, Massachusetts, 02115, USA

[14]Department of Internal Medicine, Cardiology, University of Michigan, E. Catherine St., Ann Arbor, Michigan, 48109, USA

[15]Institute for Cardiogenetics, University of Lübeck, Lübeck, 23562, Germany [16]DZHK (German Research Center for Cardiovascular Research), partner site Hamburg-Lübeck-Kiel, Lübeck, Germany

[17]Medical Research Council Population Health Research Unit, CTSU - Nuffield Department of Population Health, Medical Sciences Division, University of Oxford, Roosevelt Drive, Oxford, OX3 7LF, UK

[18]Laboratory for Cardiovascular Genomics and Informatics, RIKEN Center for Integrative Medical Sciences, 1-7-22 Suehiro-cho, Tsurumi-ku, Yokohama, Kanagawa, 230-0045, Japan

[19]Laboratory for Statistical and Translational Genetics, RIKEN Center for Integrative Medical Sciences, 1- 7-22 Suehiro-cho, Tsurumi-ku, Yokohama, Kanagawa, 230-0045, Japan

[20]K.G. Jebsen Center for Genetic Epidemiology, Department of Public Health and Nursing, Norwegian University of Science and Technology, NTNU, Trondheim, Norway

[21]HUNT Research Centre, Norwegian University of Science and Technology, Levanger, Norway

[22]Department of Endocrinology, Clinic of Medicine, St. Olavs Hospital, Trondheim, Norway

[23]Human Genetics Center, Department of Epidemiology, Human Genetics, and Environmental Sciences, School of Public Health, The University of Texas Health Science Center at Houston, 1200 Pressler St., Houston, TX, 77030, USA

[24]School of Health Science & Education, Department of Nutrition-Dietetics, Harokopio University, Eleftheriou Venizelou 70, Athens, 176 71, Greece

[25]deCODE genetics/Amgen, Inc., Sturlugata 8, Reykjavik, 102, Iceland

[26]School of Engineering and Natural Sciences, University of Iceland, Sæmundargötu 2, Reykjavik, 102, Iceland

[27]German Heart Centre Munich, Department of Cardiology, Technical University of Munich, Lazarettstr. 36, Munich, 80636, Germany

[28]CTSU - Nuffield Department of Population Health, Medical Sciences Division, University of Oxford, Roosevelt Drive, Oxford, OX3 7LF, UK

[29]German Research Center for Cardiovascular Research (DZHK e.V.), partner site Munich Heart Alliance, Lazarettstr. 36, Munich, 80636, Germany

[30]Department of Genetics and Genomic Science, Icahn Institute for Genomics and Multiscale Biology, Icahn School of Medicine at Mount Sinai, New York, NY, 10029, USA

[31]The Zena and Michael A. Wiener Cardiovascular Institute, Icahn School of Medicine at Mount Sinai, New York, NY, 10029, USA

[32]Institute of Human Genetics, Helmholtz Zentrum München, German Research Center for Environmental Health, Neuherberg, Germany

[33]Klinikum rechts der Isar, Institute of Human Genetics, Technical University of Munich, Munich, Germany

[34]Department of Internal Medicine, Cardiology, University of Michigan, E. Catherine St., Ann Arbor, Michigan, 48109, US

[35]School of Medicine & University Hospital Bonn, Institute of Human Genetics, University of Bonn, Bonn, Germany

[36]Program in Biological and Biomedical Sciences, Harvard Medical School, Boston, Massachusetts, 02115, USA

[37]Faculty of Medicine, University of Iceland, Sæmundargötu 2, Reykjavik, 102, Iceland

[38]Department of Internal Medicine, Division of Cardiology, Landspitali – National University Hospital of Iceland, Hringbraut, Reykjavik, 101, Iceland

[39]Divisions of Cardiovascular Medicine and Genetics, Brigham and Women's Hospital, Harvard Medical School, 75 Francis St., Boston, MA, 02115, USA

[40]Division of Genetics and Genomics, Boston Children's Hospital, 300 Longwood Ave., Boston, MA, 02115, USA

[41]Department of Computational Biology and Medical Sciences, Graduate School of Frontier Sciences, The University of Tokyo, 4-6-1 Shirokanedai, Minato-ku, Tokyo, Tokyo, 108-8639, Japan

[42]Department of Cardiovascular Medicine, The University of Tokyo, 7-3-1 Hongo, Bunyko-ku, Tokyo, Tokyo, 113-8655, Japan
[43]Department of Cardiovascular Medicine, Mayo Clinic, MN, USA
[44]Regeneron Genetics Center, Regeneron Pharmaceuticals, 777 Old Saw Mill River Rd., Tarrytown, NY, 10591, USA
[45]Department of Cardiovascular Sciences and NIHR Leicester Biomedical Research Centre, University of Leicester, Glenfield Hospital, Groby Road, Leicester, LE3 9QP, UK
[46]Cardiovascular Genomics & Genetics, University of Arizona, College of Medicine, Phoenix, Arizona, USA
[47]Clinical Gene Networks AB, Stockholm, Sweden
[48]Department of Genetics & Genomic Sciences, Institute of Genomics and Multiscale Biology, Icahn School of Medicine at Mount Sinai, New York, USA
[49]Integrated Cardio Metabolic Centre, Karolinska Institutet, Karolinska Universitetssjukhuset, Huddinge, Sweden
[50]Human Genome Sequencing Center, Baylor College of Medicine, 1 Baylor Plaza, Houston, TX, 77030, USA
[51]Department of Clinical Sciences in Malmö, Lund University, Malmo, Sweden
[52]Second Department of Cardiology, Medical School, National and Kapodistrian University of Athens, University General Hospital Attikon, Athens, Greece
[53]Department of Cardiac Surgery and The Heart Clinic, Tartu University Hospital, Tartu, Estonia
[54]University of Balamand, East Med Res Institute, School of Medicine, P.O. Box 33, Amioun, Lebanon
[55]Harvard T.H.Chan School of Public Health, Boston, MA, USA
[56]Health Data Research UK Cambridge, Wellcome Genome Campus and University of Cambridge, Cambridge, UK
[57]The National Institute for Health Research Blood and Transplant Unit (NIHR BTRU) in Donor Health and Genomics at the University of Cambridge, Cambridge, UK
[58]Human Genetics, Wellcome Sanger Institute, Saffron Walden, UK
[59]National Institute for Health Research Cambridge Biomedical Research Centre, Cambridge University Hospitals, Cambridge, UK
[60]British Heart Foundation Centre of Excellence, Division of Cardiovascular Medicine, Addenbrooke's Hospital, Hills Road, Cambridge, UK
[61]DZHK (German Research Center for Cardiovascular Research), partner site Hamburg-Lübeck-Kiel, 23562, Germany
[62]Department of Human Genetics, University of Michigan, E. Catherine St., Ann Arbor, Michigan, 48109, USA
[63]Princess Al-Jawhara Al-Brahim Centre of Excellence in Research of Hereditary Disorders (PACER-HD), King Abdulaziz University, Jeddah, Saudi Arabia
[64]Verve Therapeutics, Cambridge, MA, 02139, USA
